# Comorbidity Exposure-Window Definitions and Multidimensional Disparities in Long COVID Risk: Evidence from a U.S. National Cohort (2020–2024)

**DOI:** 10.64898/2026.07.11.26357794

**Authors:** Yewen Chen, Zhetao Chen, Ge Yang, Bingnan Li, Kehinde Olawale Ogunyemi, Jialing Liu, Fangzhi Luo, Yuan Ke, Leonardo Martinez, Xianyan Chen, Janani Rajbhandari, Ye Shen, the National Clinical Cohort Collaborative

## Abstract

Long COVID (LC) affects millions of individuals worldwide, particularly those with preexisting comorbidities. However, when assessing comorbidity-related risk factors for LC, it remains unclear whether preexisting comorbidities should be defined prior to SARS-CoV-2 infection or LC diagnosis. This methodological choice may substantially bias estimates of LC risk. In this study, leveraging Electronic Health Records (EHR) from 6,130,413 adults with documented COVID-19 across 49 U.S. states in the National Clinical Cohort Collaborative (N3C) from 2020 to 2024, we evaluated the impact of different comorbidity exposure-window definitions on LC risk estimates. We developed ensemble cross-fitted double/debiased machine learning to adjust for individual- and county-level confounders. We found that defining comorbidities before infection rather than before LC diagnosis yielded 8%–314% higher adjusted attributable risks and 3%–34% higher adjusted relative risks across 16 major comorbidities. Comorbidity-associated risks generally declined from 2020 to 2024, while substantial demographic, socioeconomic, geographic, and multimorbidity-related disparities persisted. Exposure-window specification can be a major source of bias in estimates of LC risk, particularly for comorbidities prone to delayed clinical recognition or underascertainment. More broadly, our results highlight how temporal misclassification of exposures in longitudinal EHR studies can distort risk attribution and population-level inference for risk stratification for LC and prioritization of prevention strategies in clinical medicine and public health, respectively.

## 1 Introduction

Despite reductions in acute morbidity and mortality associated with Coronavirus Disease 2019 (COVID–19), concerns about its long–term health trajectory remain substantial.^1^ Chronic conditions occurring after Severe Acute Respiratory Syndrome Coronavirus 2 (SARS-CoV-2) infection, commonly referred to as post-COVID-19 conditions or long COVID (LC), are not uncommon.^1–3^ For example, as of early 2023, approximately 18.4 million adults in the United States (U.S.) had experienced LC.^4^

Accurately identifying populations at elevated risk remains important for prevention, surveillance, and clinical planning, because LC can impair physical functioning and mental health, potentially leading to increased healthcare utilization and associated costs.^5,6^ Prior studies have associated LC with demographic and clinical factors, including sex, age, and chronic conditions including type 2 diabetes, cardiovascular disease, chronic kidney disease, depression, and others.^7,8^ However, much of this evidence has been derived from early-pandemic data, hospitalized or geographically restricted populations, or analyses in which comorbidities were defined only before SARS-CoV-2 infection.^7,9,10^ These limitations leave two unresolved questions: whether comorbidity-associated LC risk estimates change substantially when the comorbidity exposure-window is defined differently, and whether these estimates vary across later pandemic phases and population subgroups.

The comorbidity exposure-window definition has important epidemiologic implications. Comorbidities documented before COVID-19 infection reflected baseline vulnerability to subsequent LC, whereas comorbidities documented before LC diagnosis captured a broader prediagnostic vulnerability that includes both pre-infection comorbidities and conditions emerging after infection. In clinical practice, delayed diagnosis and underascertainment of chronic diseases are common, particularly among individuals who are uninsured or have limited access to healthcare.^11–13^ Following acute COVID-19 infection, many individuals can have increased healthcare utilization and greater opportunities for clinical evaluation than they had before the pandemic.^14,15^ As a result, an underlying comorbidity that was already present before COVID-19 infection but remained undiagnosed or underascertained may subsequently be diagnosed after infection. Given evidence that several health problems can persist or worsen for up to 3 years after infection^8^ and that a higher proportion of individuals experienced multiple COVID-19 infections between 2020 and 2024,^16^ the interval between the initial COVID-19 infection and the initial LC diagnosis may have been relatively long for some individuals. Post-infection comorbidities identified in these individuals may also provide meaningful information and should be considered in LC risk assessment. Comparing the pre-infection and pre-LC comorbidity exposure windows may help distinguish baseline vulnerability from broader clinical complexity before LC diagnosis and determine whether pre-infection comorbidities alone are sufficient to characterize the risk of developing LC.

In this study, we used electronic health record (EHR) data between 2020 and 2024 in the National Clinical Cohort Collaborative (N3C).^17^ This multiyear national observation window is essential for estimating temporal trends in LC burden rather than relying solely on early-pandemic risk-factor estimates. It also provides an opportunity to examine comorbidity exposure windows in the context of changing virus variants, vaccination patterns, reinfections, clinical recognition, and health care utilization.^3,18^ By further linking individual-level N3C data with county-level measures of health resources, social vulnerability, and urbanicity,^19–21^ it enables assessment of LC risk heterogeneity across demographic, geographic, and temporal contexts, rather than estimating a single average comorbidity effect. To robustly estimate and compare LC risk in the setting of high-dimensional data and binary exposure and outcome variables, we developed an ensemble Double/Debiased Machine Learning (DML).^22–24^

## 2 Results

### Descriptive results

The number of diagnosed LC cases from 2020 to 2024 was 88,586 (Supplementary Table S2). Among the study population, 2,720,949 (44.4%) individuals had at least one preexisting comorbidity (Supplementary Table S9). Compared with individuals without LC, those with LC were older (mean age 58.3 vs. 51.9 years), more likely to be female (64.2% vs. 57.0%), and had higher BMI (34.4 vs. 32.4). Individuals with LC also had substantially higher prevalences of preexisting comorbidities and their multimorbidity (Table S2). The three most common preexisting comorbidities in this population were hypertension (1,562,008 cases; 25.5%), neuropsychiatric disorders (969,103 cases; 15.81%), and cancer (943,600 cases; 15.39%). In contrast, dementia was the least common, with 83,137 cases (1.36%). Higher percentages of individuals with medical conditions, including LC, were observed in the older adult group (65 years and above) than in the younger group (under 65 years, Table S9). For each comorbidity, absolute yearly case counts were generally higher in urban areas compared to rural areas; conversely, the prevalence of some comorbidities from rural areas were higher in certain years, such as hypertension (Supplementary Table S10).

### Risks of preexisting comorbidities on developing LC

Under the four analytic cohorts (Table 1; Supplementary Section S1.1), all preexisting comorbidities were significantly associated with an increased risk of developing LC, as indicated by positive aARs with 95% CIs that did not include zero (Table 2 and Supplementary Table S14). Overall, Cohort I showed higher aAR and aRR estimates than Cohort II across all comorbidities. Among individual conditions, chronic lung disease exhibited the strongest association with LC risk, with the highest aARs in both Cohort I (1.494%; 95% CI: 1.452%–1.535%) and Cohort II (1.091%; 1.054%–1.128%). It also showed the largest aRRs, at 2.156 (95% CI: 2.121–2.192) and 1.813 (1.783–1.844), respectively. Other highly ranked comorbidities included peripheral vascular disease, cerebrovascular disease, heart failure, cardiomyopathy, coronary artery disease, and depression, all of which demonstrated elevated aARs exceeding 1.3% and aRRs ranging from 1.966 to 2.046 in Cohort I. In contrast, dementia, diabetes without complications, and cancer-related conditions showed comparatively smaller associations with LC risk. Dementia had the lowest aARs in both cohorts, with 0.728% (95% CI: 0.645%–0.811%) in Cohort I and 0.176% (0.100%–0.253%) in Cohort II, alongside the lowest aRRs of 1.494 (1.437–1.552) and 1.118 (1.067–1.170), respectively. Diabetes without complications also demonstrated relatively modest associations compared with other comorbidities.

**Table 1.** Definitions of target-comorbidity status (i.e., *X_i_*_,*c*_) across Cohorts I–IV. Cohorts I, III, and IV were designed to investigate the association between pre-infection comorbidities and LC risk, whereas Cohort II was designed to evaluate the association between pre-LC comorbidities and LC risk. Cohort I used the COVID-19 diagnosis date as the sole cutoff for determining exposure status, which may introduce two limitations: (a) the absence of a minimum time interval between target-comorbidity documentation and LC diagnosis; and (b) potential exposure misclassification among individuals with the target-comorbidity documented after infection but before LC diagnosis. Cohorts II--IV were designed to address limitation (a) by imposing a minimum interval of 90 days between target-comorbidity documentation and LC diagnosis. Cohort IV was designed to mitigate this limitation by excluding certain individuals who met the removal criteria. Cohort II addressed limitation (b) by using the date 90 days before LC diagnosis, rather than the COVID-19 diagnosis date, as the cutoff for determining exposure status. Under these definitions, Cohorts I--III shared the same analytic population, whereas Cohort IV included a smaller analytic population due to its additional exclusion criteria. Note: *E* denotes the target-comorbidity diagnosis date; *LC* denotes the Long COVID diagnosis date; *C* denotes the COVID-19 diagnosis date; and *E* = ∅ indicates no recorded target-comorbidity diagnosis.

| Cohort | $X_{i,c} = 0$ (with $E = \emptyset$ included) | $X_{i,c} = 1$ | Removed |
| --- | --- | --- | --- |
| I | With and without LC: $E \geq C$ | With and without LC: $E < C$ | — |
| II | With LC: $E \geq \min(LC - 90, C + 180)$<br>Without LC: $E \geq C + 180$ | With LC: $E < \min(LC - 90, C + 180)$<br>Without LC: $E < C + 180$ | — |
| III | With LC: $E \geq \min(C, LC - 90)$<br>Without LC: $E \geq C$ | With LC: $E < \min(C, LC - 90)$<br>Without LC: $E < C$ | — |
| IV | With LC: $E \geq LC$<br>Without LC: $E > C + 180$ | With LC: $E < \min(C, LC - 90)$<br>Without LC: $E < C$ | With LC: $\min(C, LC - 90) \leq E < LC$<br>Without LC: $C \leq E \leq C + 180$ |

**Table 2.**
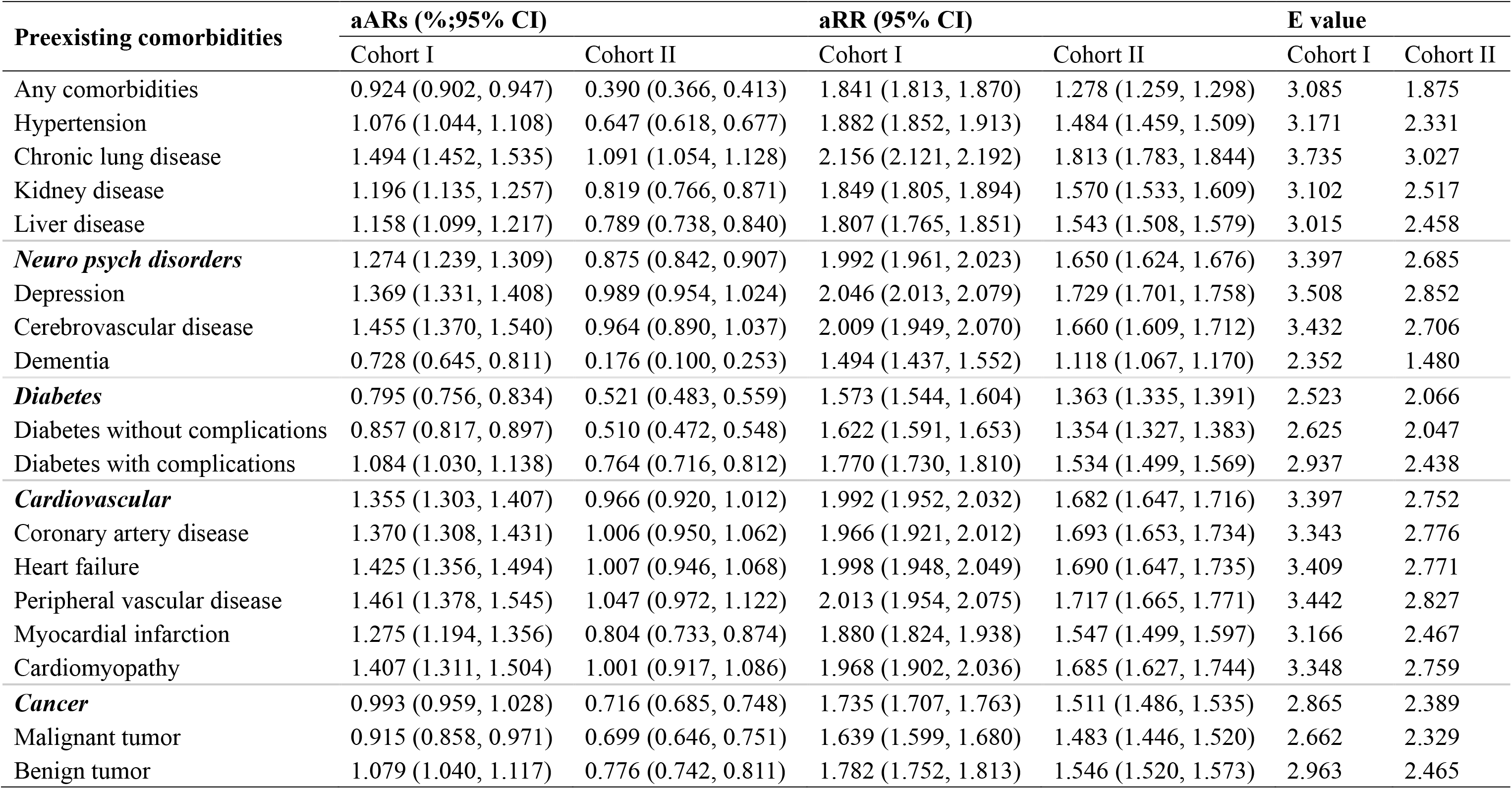
Adjusted attributable risks and relative risks (95% CIs) and sensitivity analysis using E values for pre-infection comorbidities in Cohort I and pre-LC comorbidities in Cohort II

| Preexisting comorbidities | aARs (%;95% CI) |  | aRR (95% CI) |  | E value |  |
| --- | --- | --- | --- | --- | --- | --- |
|  | Cohort I | Cohort II | Cohort I | Cohort II | Cohort I | Cohort II |
| Any comorbidities | 0.924 (0.902, 0.947) | 0.390 (0.366, 0.413) | 1.841 (1.813, 1.870) | 1.278 (1.259, 1.298) | 3.085 | 1.875 |
| Hypertension | 1.076 (1.044, 1.108) | 0.647 (0.618, 0.677) | 1.882 (1.852, 1.913) | 1.484 (1.459, 1.509) | 3.171 | 2.331 |
| Chronic lung disease | 1.494 (1.452, 1.535) | 1.091 (1.054, 1.128) | 2.156 (2.121, 2.192) | 1.813 (1.783, 1.844) | 3.735 | 3.027 |
| Kidney disease | 1.196 (1.135, 1.257) | 0.819 (0.766, 0.871) | 1.849 (1.805, 1.894) | 1.570 (1.533, 1.609) | 3.102 | 2.517 |
| Liver disease | 1.158 (1.099, 1.217) | 0.789 (0.738, 0.840) | 1.807 (1.765, 1.851) | 1.543 (1.508, 1.579) | 3.015 | 2.458 |
| ***Neuro psych disorders*** | 1.274 (1.239, 1.309) | 0.875 (0.842, 0.907) | 1.992 (1.961, 2.023) | 1.650 (1.624, 1.676) | 3.397 | 2.685 |
| Depression | 1.369 (1.331, 1.408) | 0.989 (0.954, 1.024) | 2.046 (2.013, 2.079) | 1.729 (1.701, 1.758) | 3.508 | 2.852 |
| Cerebrovascular disease | 1.455 (1.370, 1.540) | 0.964 (0.890, 1.037) | 2.009 (1.949, 2.070) | 1.660 (1.609, 1.712) | 3.432 | 2.706 |
| Dementia | 0.728 (0.645, 0.811) | 0.176 (0.100, 0.253) | 1.494 (1.437, 1.552) | 1.118 (1.067, 1.170) | 2.352 | 1.480 |
| ***Diabetes*** | 0.795 (0.756, 0.834) | 0.521 (0.483, 0.559) | 1.573 (1.544, 1.604) | 1.363 (1.335, 1.391) | 2.523 | 2.066 |
| Diabetes without complications | 0.857 (0.817, 0.897) | 0.510 (0.472, 0.548) | 1.622 (1.591, 1.653) | 1.354 (1.327, 1.383) | 2.625 | 2.047 |
| Diabetes with complications | 1.084 (1.030, 1.138) | 0.764 (0.716, 0.812) | 1.770 (1.730, 1.810) | 1.534 (1.499, 1.569) | 2.937 | 2.438 |
| ***Cardiovascular*** | 1.355 (1.303, 1.407) | 0.966 (0.920, 1.012) | 1.992 (1.952, 2.032) | 1.682 (1.647, 1.716) | 3.397 | 2.752 |
| Coronary artery disease | 1.370 (1.308, 1.431) | 1.006 (0.950, 1.062) | 1.966 (1.921, 2.012) | 1.693 (1.653, 1.734) | 3.343 | 2.776 |
| Heart failure | 1.425 (1.356, 1.494) | 1.007 (0.946, 1.068) | 1.998 (1.948, 2.049) | 1.690 (1.647, 1.735) | 3.409 | 2.771 |
| Peripheral vascular disease | 1.461 (1.378, 1.545) | 1.047 (0.972, 1.122) | 2.013 (1.954, 2.075) | 1.717 (1.665, 1.771) | 3.442 | 2.827 |
| Myocardial infarction | 1.275 (1.194, 1.356) | 0.804 (0.733, 0.874) | 1.880 (1.824, 1.938) | 1.547 (1.499, 1.597) | 3.166 | 2.467 |
| Cardiomyopathy | 1.407 (1.311, 1.504) | 1.001 (0.917, 1.086) | 1.968 (1.902, 2.036) | 1.685 (1.627, 1.744) | 3.348 | 2.759 |
| ***Cancer*** | 0.993 (0.959, 1.028) | 0.716 (0.685, 0.748) | 1.735 (1.707, 1.763) | 1.511 (1.486, 1.535) | 2.865 | 2.389 |
| Malignant tumor | 0.915 (0.858, 0.971) | 0.699 (0.646, 0.751) | 1.639 (1.599, 1.680) | 1.483 (1.446, 1.520) | 2.662 | 2.329 |
| Benign tumor | 1.079 (1.040, 1.117) | 0.776 (0.742, 0.811) | 1.782 (1.752, 1.813) | 1.546 (1.520, 1.573) | 2.963 | 2.465 |

For individuals with any preexisting comorbidity, aARs were 0.924% (95% CI: 0.902%–0.947%) in Cohort I and 0.390% (0.366%–0.413%) in Cohort II, corresponding to aRRs of 1.841 (1.813–1.870) and 1.278 (1.259–1.298), respectively. Across broader disease categories, cardiovascular conditions and neuropsychological disorders showed consistently strong associations with LC. Cardiovascular diseases exhibited aARs of 1.355% (1.303%–1.407%) in Cohort I and 0.966% (0.920%–1.012%) in Cohort II, with corresponding aRRs of 1.992 (1.952–2.032) and 1.682 (1.647–1.716). Neuropsychological disorders similarly demonstrated elevated risks, with aARs of 1.274% (1.239%–1.309%) and 0.875% (0.842%–0.907%) and aRRs of 1.992 (1.961–2.023) and 1.650 (1.624–1.676) across the two cohorts, respectively.

### Risks of multimorbidity on developing LC

For each target preexisting comorbidity, our results showed increasing trends in both aR_0_ and aR_1_, but declining trends in both aARs and aRRs, as the number of additional comorbidities increased from none to two or more (Supplementary Figures S2–S3). Moreover, the changes in aARs and aRRs were generally larger when moving from no additional comorbidity to one additional comorbidity than when moving from one to two or more additional comorbidities.

### Disparities in risk estimates of LC across cohorts

Among the four analytic cohorts, Cohort II, which defined comorbidities using the pre-LC exposure-window, generally yielded the smallest aAR and aRR estimates, whereas Cohort I, which used the pre-infection exposure-window without imposing a minimum interval between comorbidity documentation and LC diagnosis, generally yielded the largest estimates (Tables 2 and S14). Interestingly, although Cohort IV excluded individuals with comorbidities first documented after COVID-19 infection, its estimates of aR_0_, aR_1_, aAR, and aRR were generally similar to those from Cohort III, which did not apply this exclusion (Supplementary Tables S11-S12 and S14, Supplementary Figures S13-S16). Overall, these cohorts showed only small differences in aR_0_ values across comorbidties, whereas substantial differences were observed in aR_1_, aAR, and aRR.

Despite substantial differences in LC risk estimates across the four cohorts for each comorbidity, a discernible pattern was observed in the differences of such estimates across individual comorbidities associated with the pre-infection and pre-LC exposure-window definitions. We quantified these differences by comparing aRR between Cohorts I and II (Table 2). Comorbidities characterized by prolonged clinical courses or greater susceptibility to delayed clinical recognition or underascertainment usually tended to exhibit larger differences in aRR estimates. For example, dementia showed the largest relative reduction in aRR, decreasing from 1.494 in Cohort I to 1.118 in Cohort II, corresponding to a relative reduction of 25.17%. Hypertension exhibited the second-largest relative reduction (21.15%), followed by cerebrovascular disease (17.37%). Other comorbidities with relatively large reductions included myocardial infarction (17.71%), cerebrovascular disease (17.37%), chronic lung disease (15.91%), and depression (15.49%). In contrast, smaller relative reductions were observed for certain comorbidities that may be more likely to receive clinical attention and be documented in EHRs, such as malignant tumors (9.52%), which showed a smaller reduction than benign tumors (13.24%). Similarly, diabetes with complications showed a smaller relative reduction (13.33%) than diabetes without complications (16.52%), potentially reflecting differences in clinical recognition and documentation.

### Disparities across subpopulations

Compared with White individuals, Asian individuals generally exhibited lower aARs, whereas Black individuals tended to have higher aARs across most comorbidities (Figures 2a and 2b). Similar patterns were observed for aRRs (Supplementary Figures S6a and S6b). Rural residents generally exhibited higher aARs than urban residents (Figures 2c and 2d), although differences in aRRs were typically modest across comorbidities (Figures S6c and S6d). Individuals aged 65 years and older showed higher adjusted baseline and exposed risks (Supplementary Figures S4e and S4f; Supplementary Figures S5e and S5f), whereas individuals younger than 65 tended to have larger aARs and aRRs, suggesting stronger absolute and relative risk contrasts despite lower overall risks (Figures 2e and 2f; Figures S6e and S6f). Females generally exhibited higher LC risks than males, with higher aR_0_ and aR_1_ values across nearly all investigated comorbidities (Figures S4g and S4h; Figures S5g and S5h) and slightly higher aARs for most conditions (Figures 2g and 2h), whereas aRRs were not consistently higher among females (Figures S6g and S6h).

**Figure 1:**
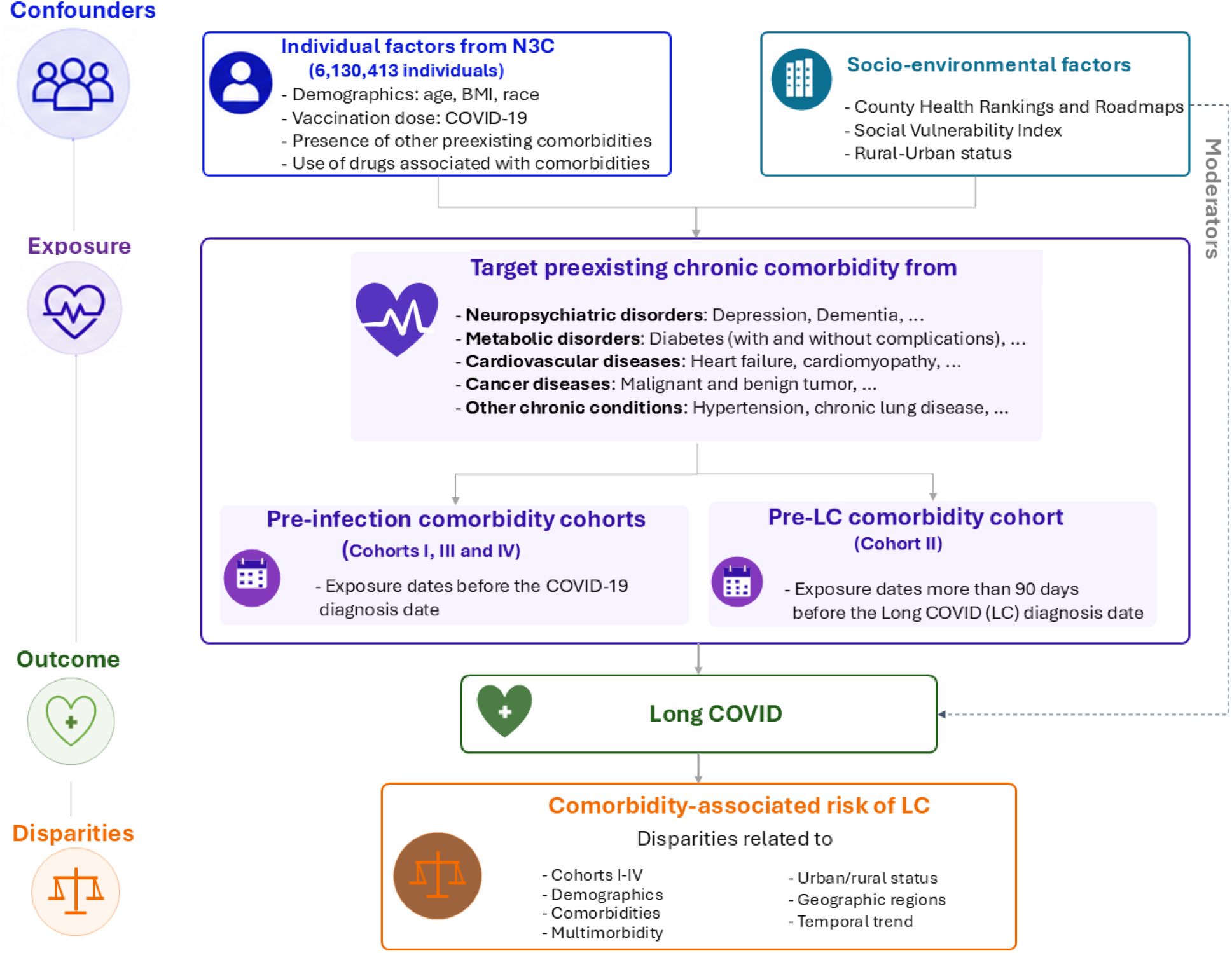
Study framework for investigating the impact of comorbidity exposure windows on the risk of developing LC and associated multidimensional disparities, using individual-level variables from N3C and county-level socio-environmental variables from three additional data sources. Three pre-infection chronic comorbidity cohorts (the primary Cohort I and supplementary Cohorts III–IV) and one pre-LC chronic comorbidity cohort (the primary Cohort II) were constructed based on 16 comorbidities representing leading health burdens in the U.S., as well as five combinations of these comorbidities.

**Figure 2:**
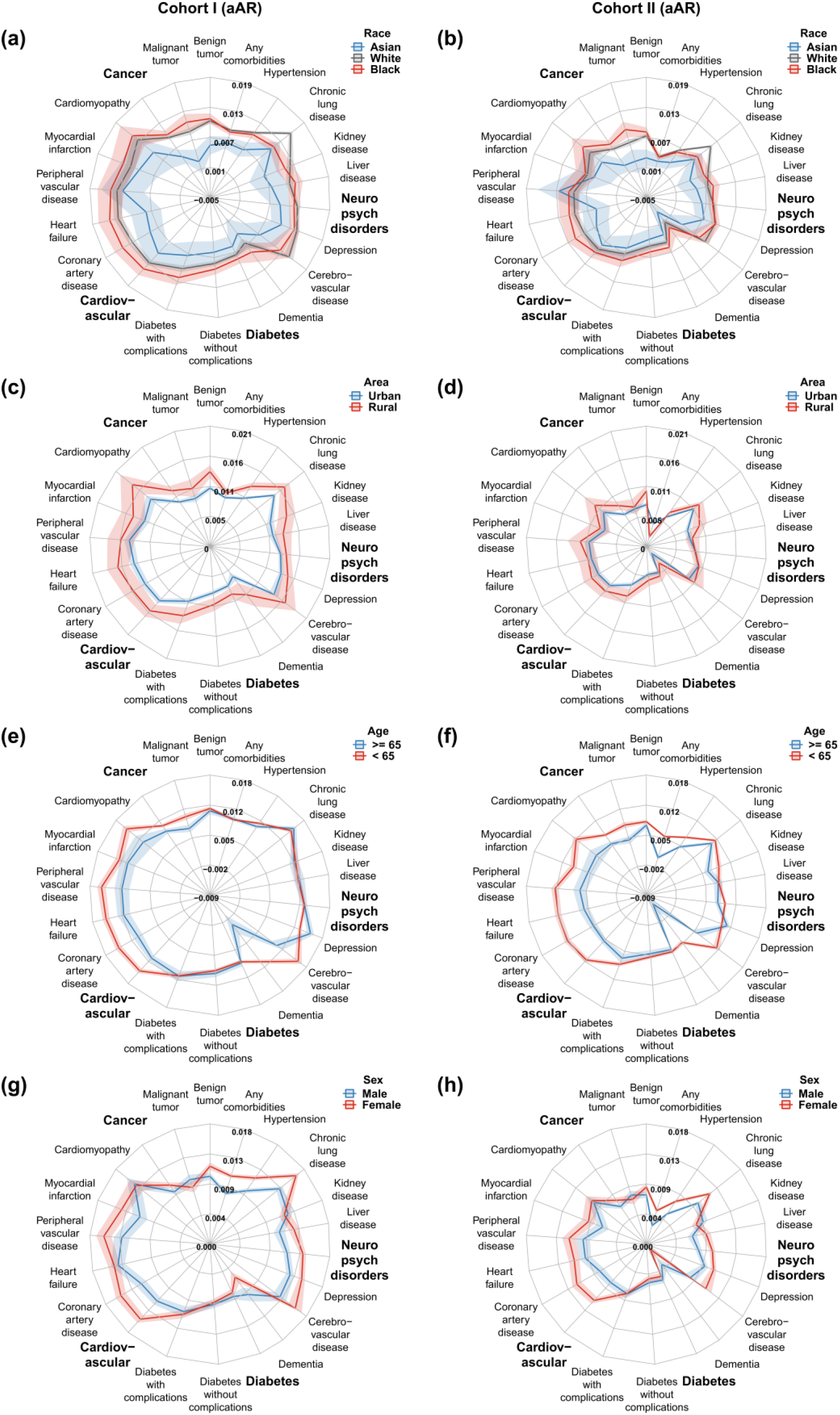
Adjusted attributable risks (aARs; %) of LC associated with comorbidities across subpopulations, with 95% confidence intervals.

### Disparities across regions

Regional analyses showed geographic heterogeneity in comorbidity-associated LC risk (Supplementary Table S7; Figure 3; and Supplementary Figures S7-S12). Adjusted absolute risk differences (aAR) were generally highest in the Midwest and West and lowest in the Northeast. Higher baseline risks (aR_0_) were identified in the South compared to the other regions (Figures S7 and S9). The South and West usually showed higher aR_1_ values (Figure S8). Due to the moderately high aR_0_ and relatively lower aR_1_ in the Northeast, the aAR and aRR were generally lower there than in the other regions, whereas the highest aRR was observed in the Midwest across comorbidities (Figure 3 and Supplementary Figures S11-S12).

**Figure 3:**
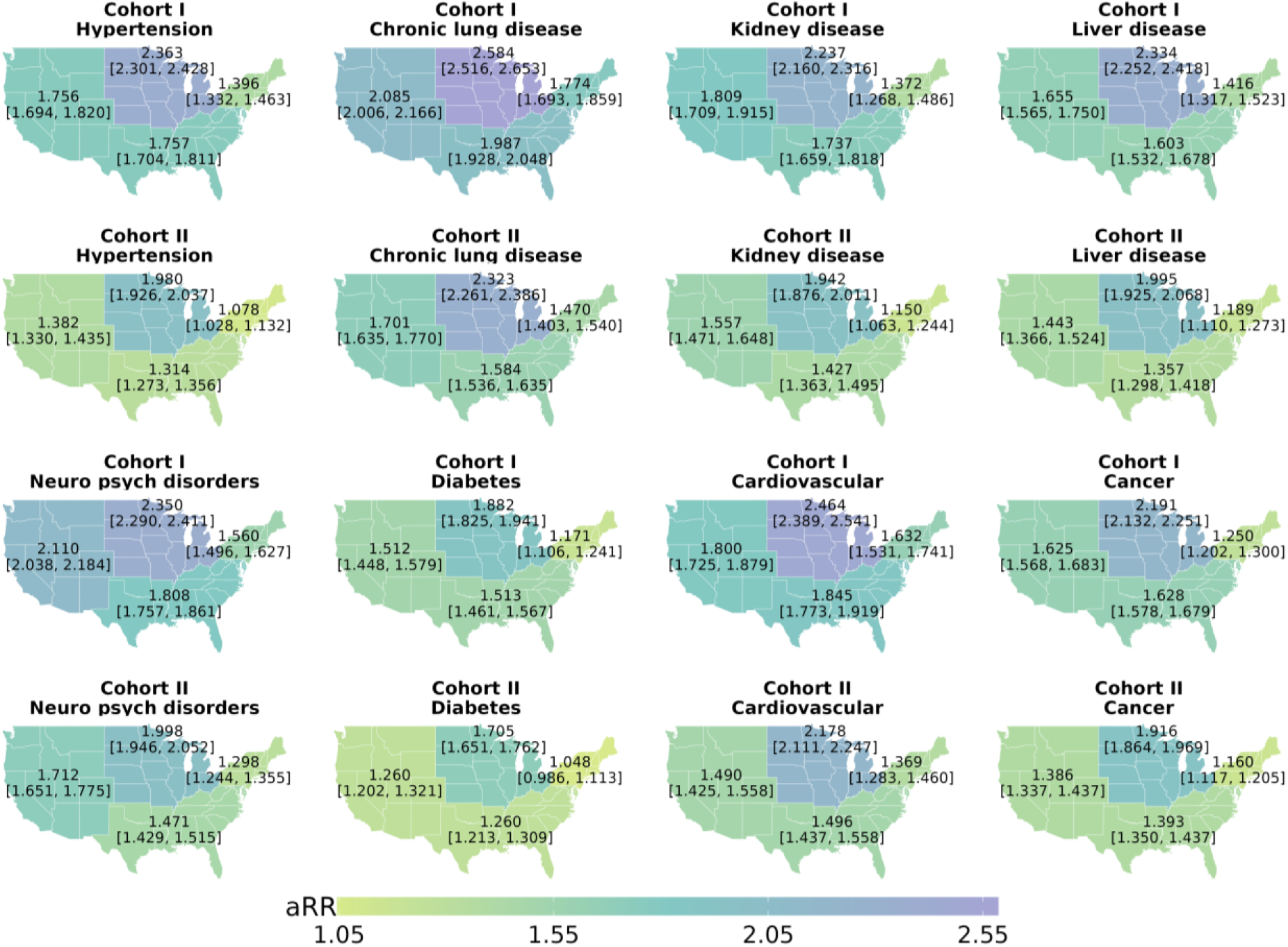
The adjusted relative risks (aRRs; %) of LC associated with comorbidities across census regions, with 95% confidence intervals.

### Disparities in temporal trends of comorbidity–associated LC risk

The 2020–2024 observation period enabled estimation of risk trends across multiple pandemic phases (Figures 4-5; Supplementary Figures S13-S16). Most comorbidities in all cohorts exhibited a generally decreasing trend in aARs and aRRs from 2020 to 2024, suggesting temporal changes in the epidemiology of LC. Several conditions showed modest increases in aARs and aR_1_ from 2020 to 2021 while aR_0_ of LC generally increased before stabilizing over the period from 2020 to 2024 for all comorbidities. Dementia was a notable exception, showing an overall stationary trend for both aARs and aRRs. Cohort I generally exhibited the highest aARs and aRRs over time, whereas Cohort II showed the lowest, with Cohorts III and IV yielding similar estimates.

**Figure 4:**
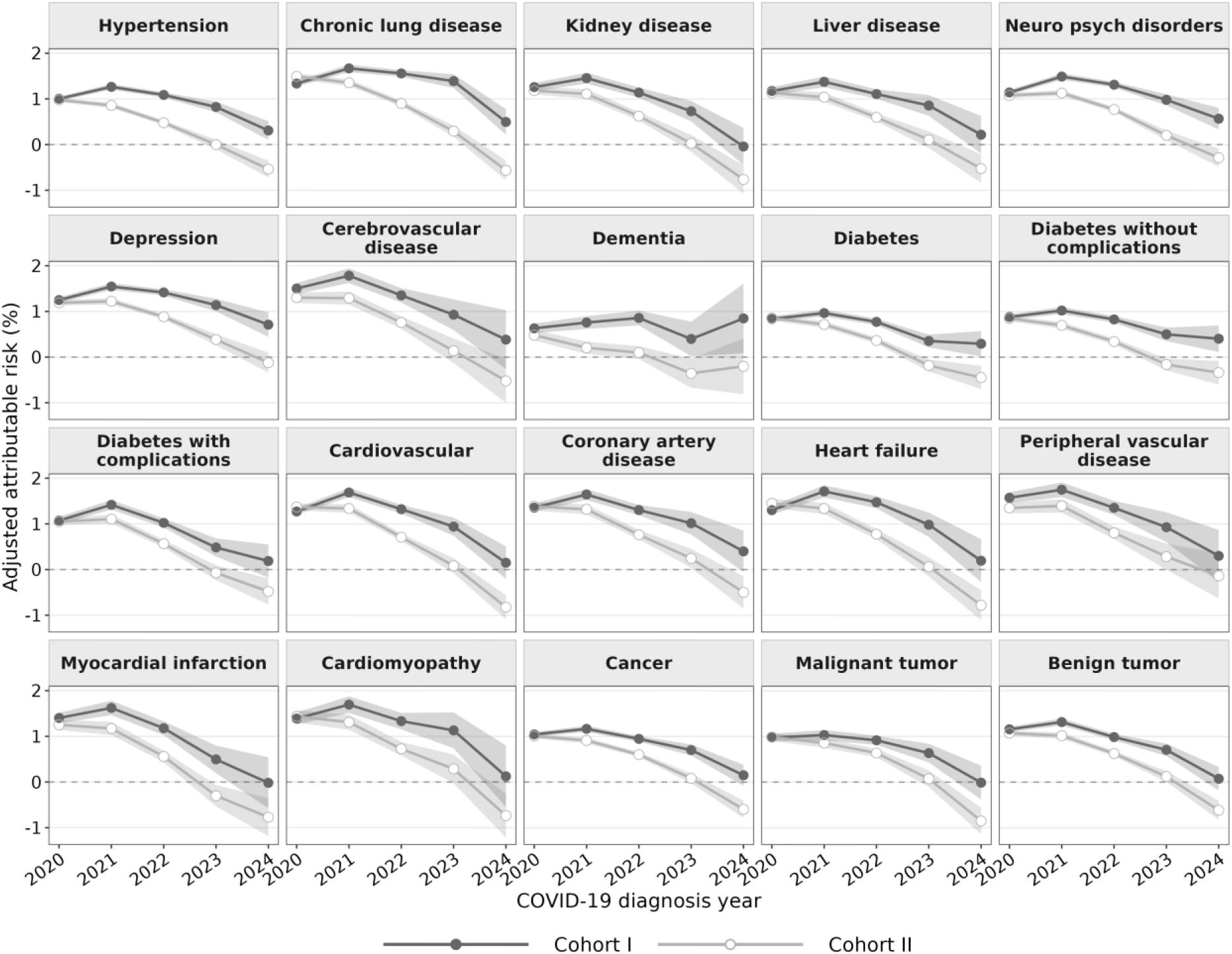
Temporal trends in adjusted attributable risks of LC associated with comorbidities, with 95% confidence intervals.

**Figure 5:**
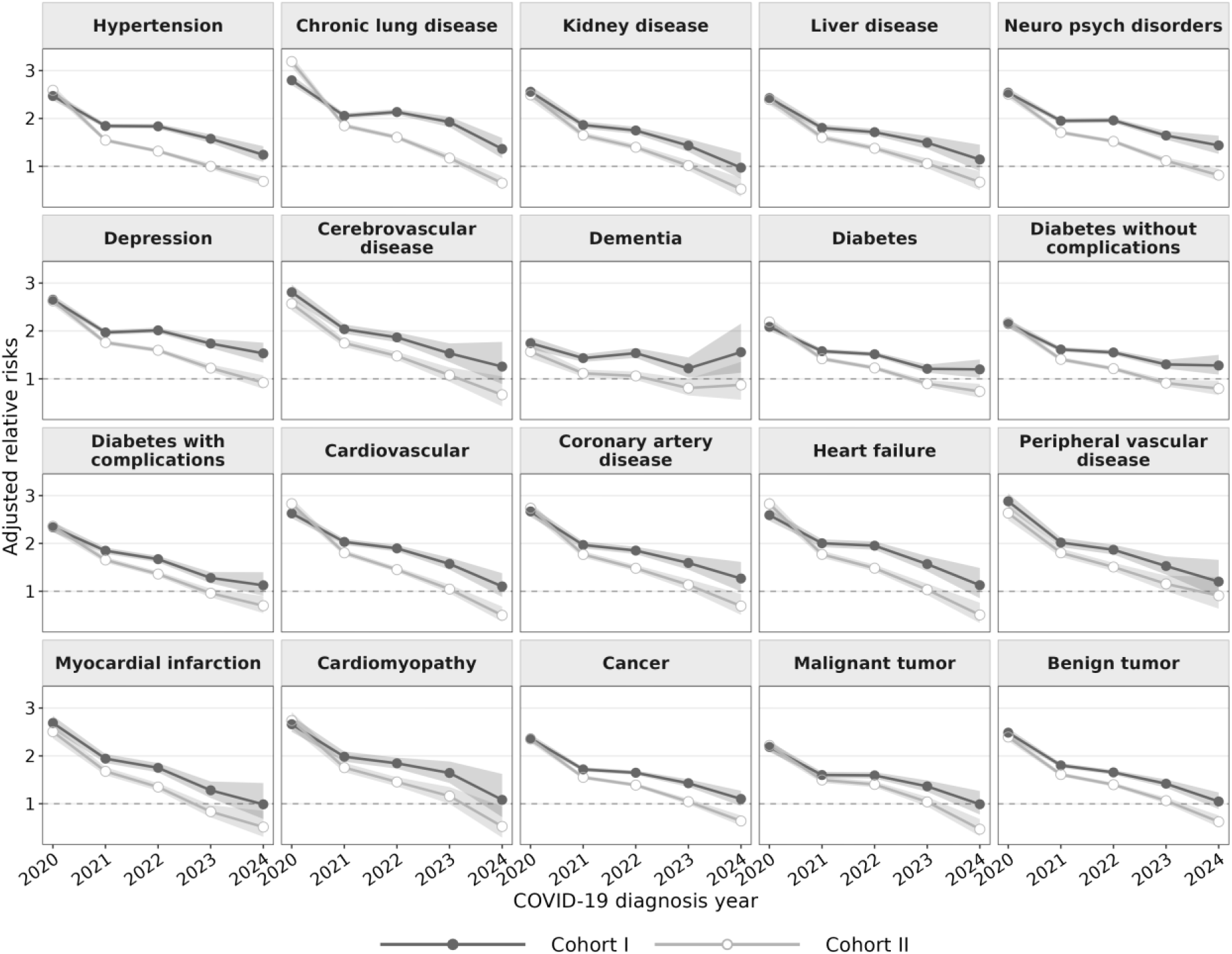
Temporal trends in adjusted relative risks of LC associated with comorbidities, with 95% confidence intervals.

### Sensitivity analysis

Most covariates achieved acceptable balance for the majority of comorbidities, as indicated by standardized mean differences (SMDs) before and after inverse probability weighting (Supplementary Figures S17–S20). Residual imbalance was observed for a limited number of covariates in some cases, potentially reflecting their strong associations with both exposure and outcome or their possible roles as mediators rather than confounders. Additionally, the propensity score distributions for many comorbidities showed substantial overlap (Supplementary Figures S21–S22). For comorbidities with limited propensity score overlap and residual covariate imbalance, the corresponding estimates should be interpreted cautiously because they may be more sensitive to violations of the positivity assumption and residual confounding.

E-values were generally large across comorbidities, ranging from 1.480 to 3.735, suggesting that substantial unmeasured confounding would be required to fully explain away the observed associations (Table 2). Chronic lung disease yielded the largest E-values in both cohorts (3.735 in Cohort I and 3.027 in Cohort II), followed by depression (3.508 and 2.852), peripheral vascular disease (3.442 and 2.827), cerebrovascular disease (3.432 and 2.706), and heart failure (3.409 and 2.771), respectively, suggesting that these associations were relatively robust to potential unmeasured confounding.

To investigate the robustness of the primary findings, which excluded individuals who died within 180 days of COVID-19 infection and employed imputation for missing data, two sensitivity analyses were conducted. Scenario 1 used a complete-case approach, excluding all participants with missing data, whereas Scenario 2 included individuals who died within 180 days of infection. Results from both sensitivity analyses were basically consistent with the primary findings (Supplementary Table S13). Additionally, the two supplementary pre-infection cohorts (Cohorts III and IV) exhibited overall patterns across comorbidities like those observed in Cohorts I and II (Supplementary Table S13 and Table 2, respectively).

## 3 Discussion

In this national EHR-based dataset of more than 6 million adults with documented COVID-19 infection over a five-year period, we developed flexible machine learning methods for assessing comorbidity–associated LC risks under different comorbidity exposure-window designs. We constructed multiple analytic cohorts (Cohorts I–IV) to systematically investigate the impact of two types of comorbidity exposure windows, pre-infection and pre-LC, on the estimated risk of developing LC. This window design extended prior risk-factor studies by distinguishing baseline pre-infection susceptibility from broader pre-diagnostic clinical vulnerability.^7–10^ We examined the importance of imposing a minimum interval between comorbidity documentation and LC diagnosis when evaluating LC risk using the HER data. We further characterized the risk disparities of LC attributable to a wide range of leading comorbidities.

We found that adjusted baseline risks (aR_0_) did not differ significantly across Cohorts I–IV, likely reflecting the robustness of aR_0_ estimation in the large populations without the target comorbidity, whereas significant differences were more often observed in aR_1_, which may reflect greater sensitivity of aR_1_ estimation in the relatively small populations with the target comorbidity (Supplementary Section S1.3). Thus, comorbidity exposure-window definitions appeared to have a greater impact on aR_1_ estimates than on aR_0_ estimates (Supplementary Tables S12-S13, and Supplementary Figures S13-S14). When similar criteria were applied, Cohort IV yielded aAR and aRR estimates comparable to those of Cohort III, despite its smaller study population, supporting the robustness of our estimation method. Among Cohorts I–III, which were based on the same study population, the substantial differences in risk estimates between Cohort II and the other cohorts highlight the importance of the choice between pre-infection and pre-LC exposure windows. When no restriction was imposed on the minimum temporal separation between comorbidity documentation and LC diagnosis, Cohort I yielded the largest aAR and aRR estimates among all cohorts. This further underscores the sensitivity of the resulting risk estimates to the definition of the exposure-window.

Distinguishing between exposure windows may be particularly important for certain comorbidities when assessing the risk of developing LC. Notably, the discrepancy between pre-infection and pre-LC risk estimates tended to be greater for comorbidities with more prolonged clinical courses than for those with shorter courses (Table 2 and Supplementary Table S14). On one hand, one plausible explanation is that chronic comorbidities with less acute clinical manifestations, such as hypertension, may be more susceptible to delayed recognition or underascertainment before COVID-19 infection due to various factors, such as limited access to or utilization of healthcare services.^11–13^ Therefore, potential misclassification or exclusion usually resulted in smaller denominators (i.e., the exposed population at risk) and higher aR_1_ relative to aR_0_ in Cohorts I, III and IV than in Cohort II, potentially leading to overestimated aARs and aRRs (Section S1.3). On the other hand, the physiological stress associated with the COVID-19 pandemic, together with increased healthcare contact, may subsequently reveal previously subclinical or undocumented conditions, leading to their first documentation during the post-infection period.^14,15^ This interpretation was also consistent with the observations in Supplementary Table S1, where many cases of comorbidities with prolonged clinical courses and less acute manifestations were first identified during the specified post-infection period. Further, the differences between Cohort II and the other cohorts may be more pronounced for comorbidities that COVID-19 is known to unmask, precipitate, or exacerbate, particularly in reinfection settings, where LC may be diagnosed following the latest infection, and comorbidities may have been identified during the relatively long interval between the first infection and LC diagnosis. Thus, including individuals with comorbidities diagnosed between infection and LC diagnosis from Cohort II may help reduce bias in the estimated aARs and aRRs. This inclusion resulted in a larger denominator (i.e., the exposed population at risk) and a lower aR_1_ relative to aR_0_ in Cohort II compared with the other cohorts. These mechanisms, together with those discussed earlier, may help explain why the estimated aARs and aRRs were generally larger under the pre-infection comorbidity definitions used in Cohorts I, III, and IV than under the pre-LC comorbidity definition used in Cohort II (Table 2 and Supplementary Table S14). Overall, Cohort II may help mitigate bias in LC risk estimation arising from underascertainment of preexisting comorbidities in EHR data and provide a relatively complete assessment of comorbidity-associated risk factors for LC.

Our results across cohorts consistently showed that cardiovascular–related comorbidities, chronic lung disease, and several neuropsychiatric disorders were generally associated with substantially higher risks of developing LC. These results suggest that individuals with these conditions are particularly vulnerable, underscoring the importance of considering cardiovascular, pulmonary, and neuropsychiatric health in strategies aimed at mitigating LC risk. Our findings should be interpreted in relation to prior studies that evaluated earlier COVID-19 pandemic periods or smaller regions. For instance, a rough comparison with adjusted hazard ratios reported in the United Kingdom (UK) shows that, for many comorbidities, including heart failure, peripheral vascular disease, cerebrovascular disease, cardiomyopathy, dementia, kidney disease, and myocardial infarction, there was no significant evidence of increased LC risk.^7^ In contrast, our estimated aRRs generally exceeded 1. Several factors may explain these differences. Differences in COVID–19 vaccination policies between the U.S. and the UK may be important contributors. Unlike vaccination policies in many European countries that prioritize targeted populations, the U.S. has generally maintained a broader strategy recommending seasonal COVID-19 vaccination for all individuals aged six months and older.^25^ Additionally, if high–dimensional confounders involve complex nonlinear effects, the hazard ratio models used in previous studies may fail to capture these effects, since these models often rely on linear regression assumptions. In contrast, our analysis using DML can better account for complex confounding. Other important factors may relate to population characteristics associated with health disparities and other methodological differences. Compared with previous studies, our study provided more precise risk estimates, as evidenced by substantially narrower confidence intervals. For example, a regional analysis in the U.S. reported an adjusted risk ratio of 1.72 (1.17–2.52) for depression.^9^ In contrast, the corresponding estimates in our study were 2.046 (2.013–2.079) in Cohort I and 1.729 (1.701–1.758) in Cohort II. Precise risk estimates may also be important for other LC studies, such as population attributable fraction (PAF) analyses.^26^

Previous studies provided important insights into comorbidities as risk factors for LC, but did not establish whether risk estimates differ according to when comorbidities are measured relative to LC diagnosis or how adjusted risk estimates evolved from 2020 to 2024. The early increase revealed from this study may be partly attributable to underdiagnosis of LC during the first year of the COVID–19 pandemic.^27^ The overall decreasing trend may reflect multiple concurrent changes, including vaccination coverage, healthcare utilization, coding practices, population immunity, treatment availability, and the evolving nature of SARS–CoV–2 variants. For example, variants such as Delta or Omicron, which emerged since 2021, may be associated with a lower risk of developing LC, compared to the original SARS–CoV–2 strain.^28^ Additionally, this trend may also be partly attributable to the protective effects of COVID–19 vaccination and underreporting of COVID-19 cases.^29,30^

This study revealed multidimensional disparities in comorbidity-associated risk of LC, ranging from individual-level features to regional geographic characteristics, thereby providing a more comprehensive understanding of the heterogeneity of these risks. The heterogeneity by age, sex, race, and multimorbidity burden may reflect both biological susceptibility and differences in EHR ascertainment. This may inform related health strategies. For example, contrary to prior U.S. studies reporting similar risks of LC among Black and White individuals, our findings indicate that the adjusted attributable risk of developing LC is generally highest among Black individuals, followed by White individuals and then Asian individuals, across most preexisting comorbidities.^31^

Based on the extensive geographic visualizations, we also observed substantial regional heterogeneity in comorbidity-associated LC risk across U.S. Census regions. This heterogeneity may reflect regional differences in population health profiles, healthcare access, vaccination coverage, socioeconomic conditions, and healthcare-seeking behaviors. Although rural residents generally exhibited slightly higher aARs than urban residents, the relatively modest urban–rural differences in aRRs suggest that broader geographic factors contribute more substantially to LC risk heterogeneity than urbanicity alone.

This study has several strengths. The N3C cohort provided over 6 million adults with documented COVID-19 across 49 states, supporting precise estimates across multiple comorbidities and strata. The 2020-2024 study period allowed estimation of risk trends across multiple pandemic phases, unlike studies limited to early-pandemic data.^7,9,27^ The four-cohort design enabled direct comparisons between pre-infection and pre-LC exposure windows, deepening our understanding of comorbidity-associated LC risks. Linkage with county-level data supported analysis of geographic and social-context heterogeneity, while the ensemble DML framework facilitated flexible adjustment for high-dimensional confounders and accommodated heterogeneous effects.

This study also has limitations. LC was identified from EHR diagnosis codes and may be under ascertained, particularly among individuals with mild symptoms, limited access to care, or care outside N3C-contributing sites.^3,32^ The N3C dataset is large but not a probability sample of the U.S. population, and geographic site distribution may influence regional estimates. Comparisons across cohorts should therefore be interpreted with caution given potential biases inherent in EHR data. Cohorts II-IV incorporated minimum intervals between comorbidity documentation and LC diagnosis; however, their cohort definitions were dependent on the study outcome. These observations suggest the need for further investigation of cutoff dates to ensure comparable opportunities for comorbidity ascertainment between LC and non-LC populations. Differences in ascertainment opportunities, as well as other aspects of cohort construction, may also contribute to variations in LC risk estimates beyond those attributable to exposure-window definitions. Cohort II captured prediagnostic clinical vulnerability rather than baseline susceptibility and should not be interpreted as estimating the causal effect of comorbidities that arise after COVID-19. In addition, marginal causal risk estimates should be interpreted cautiously because the corresponding causal intervention may not be well defined and modifiable when comorbidity status is treated as exposure, and the required identification assumptions for valid inference may be difficult to fully justify using observational data (Supplementary Section S4.3). Residual confounding, differential follow-up, exposure and outcome misclassification, and limited positivity for several comorbidities may remain despite DML adjustment and sensitivity analyses. Future studies could extend our framework by explicitly modeling spatiotemporal dependence in comorbidity-associated LC risks across counties and calendar periods to improve understanding of transmission dynamics.^3,33,34^

In this national EHR-based cohort spanning 2020 through 2024, estimates of comorbidity-associated LC risks were sensitive to exposure-window definitions and varied across demographic, geographic, urbanicity, multimorbidity, and temporal strata. Across 16 major comorbidities, defining comorbidities before infection (Cohorts I and III-IV) rather than before LC diagnosis (Cohort II) yielded 8%–314% higher aARs and 3%–34% higher aRRs (Table 2 and Supplementary Table S14). Our findings highlight the importance of carefully specifying comorbidity exposure windows in LC research. In particular, LC risk estimates for comorbidities more susceptible to delayed clinical recognition or underascertainment appeared to be more sensitive to exposure-window specifications than those with shorter or more acute courses. Beyond what was described, other broader clinical implications that can be discussed are the potential of our results to inform: 1) precise individual LC risk stratification with clinical risk chart for people with preexisting comorbidities, and 2) healthcare LC screening strategies prioritization among people with preexisting comorbidities.^35,36^ These findings could improve the accuracy of PAF estimates by providing exposure-window-specific risk estimates,^26^ thereby informing appropriate comorbidity risk reduction strategies for LC prevention.

## 4 Methods

This study was based on individual-level EHR data from the N3C Data Enclave, version 185. The analytic population included 6,130,413 adults aged 18 years or older with documented COVID-19 from 2020 through 2024, representing 2,864 U.S. counties across 49 states. Figure 1 presents the overall study design for this analysis, including the collection and use of confounders, the definitions of exposures and outcomes, and the assessment of multidimensional disparities in comorbidity-associated LC risk.

### National Clinical Cohort Collaborative (N3C)

N3C represents a centralized data resource established through collaboration among many organizations. The platform operates under the overall stewardship of the National Institutes of Health. Detailed descriptions of the N3C infrastructure and data harmonization processes have been published previously.^17^ To support this study, we further processed the data using a series of criteria, including matching and screening. More details of the N3C data processing workflow can be found in Supplementary Figure S1.

### Study outcome

LC after COVID–19 infection is the study outcome. The diagnosis date of COVID–19 infection was based on the ICD–10-CM U07.1 code or antibody/PCR test results. LC was defined as a binary indicator using diagnosis codes for post-COVID-19 condition or sequelae of infectious disease, following prior N3C-based definitions. Specifically, the B94.8 code was used for diagnoses prior to October 1, 2021, and both U09.9 and/or B94.8 codes were used thereafter.^18^ In practice, LC symptoms that cannot be explained by an alternative medical condition may emerge, persist, resolve, and recur over weeks, months, or even years. The duration criteria also vary across definitions, ranging from at least 28 days in earlier studies to 3 months in the WHO definition.^2,3,37^ For individuals with multiple records, the earliest qualifying dates for COVID-19 infection and LC diagnosis were used, respectively. To reduce misclassification, diagnoses before March 1, 2020, were excluded.^18^ Under these settings, we have LC as the study outcome.

### Comorbidities as exposure variables

We systematically extracted records for comorbidities representing leading health burdens in the U.S.^38^ The analysis included 16 comorbidities (Supplementary Table S1): five cardiovascular diseases (heart failure, coronary artery disease, peripheral vascular disease, cardiomyopathy, and myocardial infarction); three neurological and psychiatric disorders (cerebrovascular disease, dementia, and depression); two metabolic conditions (diabetes with and without complications); two cancer diseases (Malignant tumor and Benign tumor); hypertension; and conditions for the respiratory, renal, and hepatic systems (chronic lung disease, kidney disease, and liver disease, respectively).

For each comorbidity or combination of comorbidities, we investigated two primary scenarios (Figure 1): pre-infection comorbidities documented before COVID-19 infection (Cohort I) and pre-LC comorbidities defined based on occurrence more than 90 days before the LC diagnosis date (Cohort II). Details of the construction of these two cohorts were provided in Table 1 and Supplementary Section S1.1, where we also discussed the rationale for the criteria used in cohort construction. Cohort I represented baseline pre-infection susceptibility, whereas Cohort II captured broader prediagnostic clinical vulnerability. In addition, for the pre-infection scenario, we constructed two supplementary cohorts for more refined comparative analyses: Cohort III, which imposed a minimum temporal interval between comorbidity documentation and LC diagnosis, and Cohort IV, which excluded individuals potentially subject to exposure misclassification. In particular, a substantial proportion of individuals were identified with comorbidities between COVID-19 infection and the date 90 days prior to LC diagnosis (Supplementary Tables S1-S2). Excluding or misclassifying these cases may bias estimates of LC risk. The multiple parallel cohort definitions allowed us to evaluate how comorbidity-associated LC risks varied across exposure-window definitions and to assess whether relying solely on pre-infection comorbidities is sufficient for accurately estimating the risk of developing LC.

### Other covariates

Candidate covariates included in this study were at individual- and county-level. Individual factors included age, race, sex (male, female, no matching concept, and missing, all of which were considered in our statistical modeling analysis), medication use for comorbidities, and number of COVID-19 vaccine doses from N3C. County-level factors included social disparity data from County Health Rankings and Roadmaps (CHRR; 2020–2025),^20^ the Social Vulnerability Index (SVI),^21^ and Rural–Urban Continuum Codes (RUCC).^19^ County-level domains covers education, income, health insurance, community safety, environmental factors, socioeconomic status, household composition, racial and ethnic minority composition, housing type, vehicle access, and urbanicity (Supplementary Tables S3-S6).

### Collinearity and missing data

Because some continuous candidate variables were highly correlated, we performed an initial selection using Pearson correlation coefficients to refine the candidate covariate set (Supplementary Section S3). We finally included 60 adjustment covariates ***Z*** in the DML development, including 31 individual- and county-level variables, 14 medication indicators, and 15 comorbidity indicators (excluding target comorbidity) (Supplementary Table S8). For covariates with missing, we employed multiple imputation using the *mice* package in R.^39^

### Data analysis framework

#### Double/debiased Machine Learning (DML)

Estimating complex associations between LC and other factors requires methods that can accommodate high-dimensional and nonlinear confounding. Traditional regression approaches may be vulnerable to model misspecification, whereas prediction-oriented machine learning methods are not designed to directly estimate adjusted risk contrasts with valid statistical inference.^36,40^ DML combines flexible nuisance-function estimation with orthogonal scores and cross-fitting, reducing sensitivity to regularization bias in nuisance models while enabling flexible adjustment for high-dimensional covariates.^22–24^

#### Propensity score and potential outcome models

For each exposure definition, our DML estimated one propensity score model and two outcome models. Let *X_i_*_,*c*_ denote the binary indicator that individual *i* had the *c* th pre-infection or pre-LC comorbidity under the applicable exposure definition (Table 1). Let ***Z****_i_*_,*c*_ denote the adjustment covariates used for the *c*th comorbidity (Supplementary Table S8). The propensity score was defined as

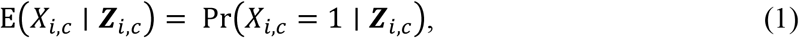

where E(·∣·) denotes the conditional expectation, which equals the conditional probability Pr(·∣·) for the binary exposure variable *X_i_*_,*c*_.

Under comorbidity status e= {0, 1}, separate potential outcome models were defined as

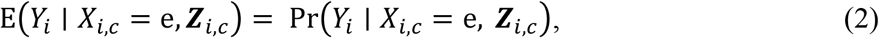

where *Y_i_* = 1 indicates that individual *i* developed LC and *Y_i_* = 0 otherwise; e = 0 and e = 1 distinguish individual *i* without and with the *c*th comorbidity, respectively.

#### Ensemble-based prediction of propensity scores and outcomes

DML estimators developed based on (1)–(2) are doubly robust when at least one of the two models is correctly specified.^41^ In our setting, however, correct specification can be challenging because the nuisance functions, involving ***Z****_i_*_,*c*_, may be high-dimensional and nonlinear. To flexibly capture these complex structures and thereby reduce bias related to model misspecification, we estimated (1)–(2) with an ensemble of logistic regression, gradient boosting machines, and random forests (Supplementary Section S4.2).^40,42,43^ In this implementation, we employed cross-fitting procedures to develop these ensemble models and then generated predictions for the potential outcomes among individuals with and without each comorbidity (Steps 1-4 of Algorithm 1).

#### Risk estimation using orthogonal score for binary outcome and exposure variables

For each comorbidity, we reported the observed LC risks among populations without and with the *c*th preexisting comorbidity, denoted as R_0_ andR_1_, based on the raw data. Correspondingly, we calculated the adjusted risks, aR_0_ and aR_1_, by combining the estimated propensity scores and predicted potential outcomes from the held-out using orthogonal score methods (Step 6 of Algorithm 1). In DML, aAR = aR_1_ − aR_0_. We also calculated AR = R_1_ − R_0_, RR = R_1_/R_0_, and aRR = aR_1_/aR_0_.^44^ All these risk estimates for each comorbidity were separately assessed across Cohorts I-IV.

#### Subpopulation analysis for heterogeneous LC risk

Algorithm 1 allows 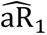 and 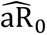 to vary with the adjustment covariates since the estimated propensity scores and predicted potential outcomes depend on ***Z****_j_*_,*c*_ (Steps 2 and 4 of Algorithm 1). For each prespecified subgroup, we calculated subgroup-specific aR_0_ and aR_1_ using these scores and outcomes based on results of Step 5 of Algorithm 1 using formulas in Step 6 (Supplementary Section S4.1).^23,45,46^ This approach substantially reduces the computational burden associated with separately refitting the DML models within each subgroup in the large N3C dataset. It also allows the complex nuisance functions in Equations (1)-(2) to be learned from the full analytical cohort, improving the robustness and stability of the estimated aR_0_and aR_1_.

##### Algorithm 1.

Ensemble DML for binary exposure and outcome variables to assess heterogeneous Long COVID risk associated with the *c*th preexisting comorbidity.

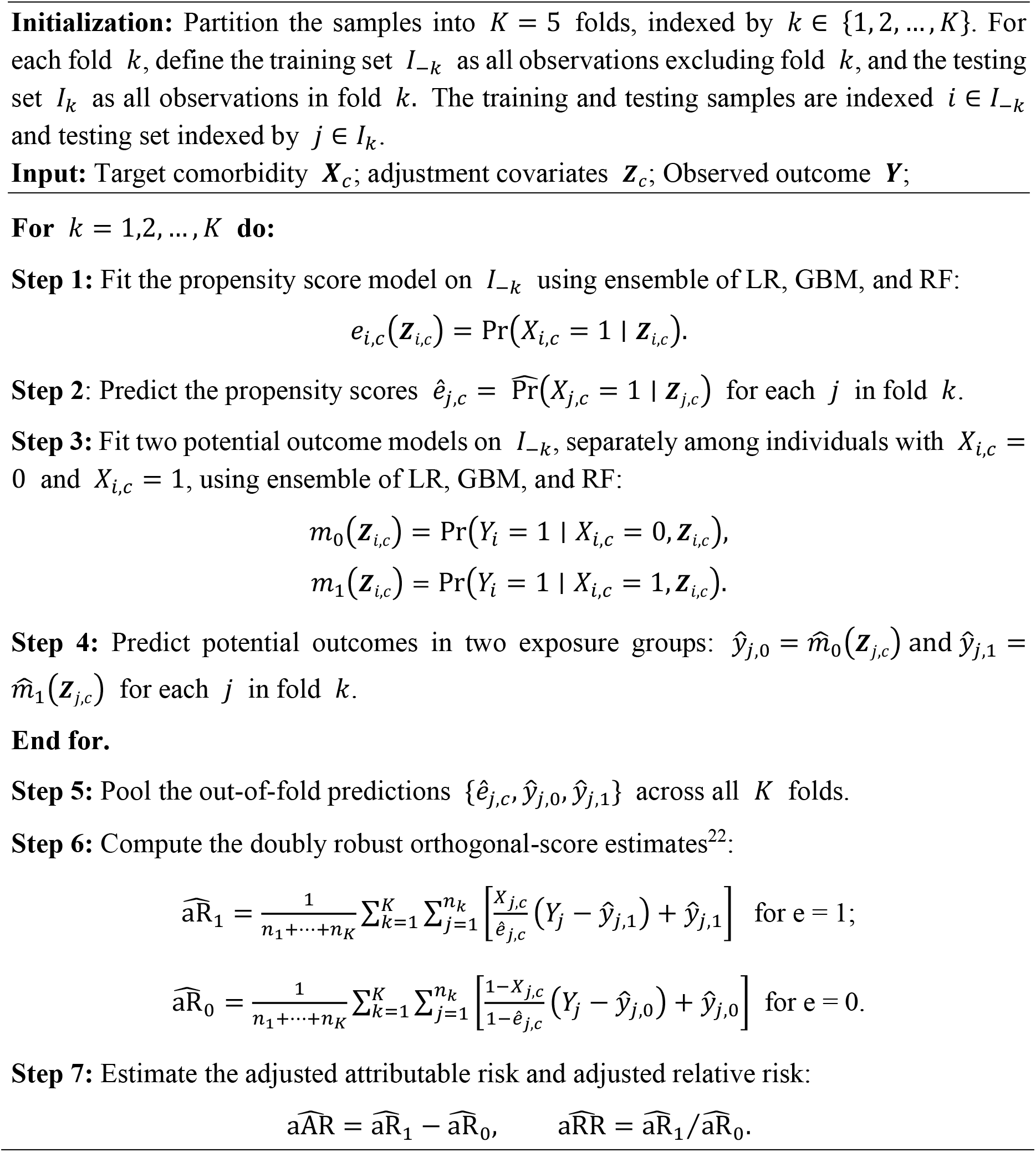

### Sensitivity analysis designs

We examined the robustness of our results through three perspectives. First, we assessed the potential impact of unmeasured confounding by calculating the E–value using the formula, 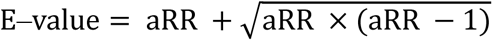.^47,48^ This metric quantifies the minimum magnitude of unmeasured confounding required to explain away the estimated association. Second, we modeled the N3C data under two distinct scenarios for two primary Cohorts I-II. In the first scenario, we incorporated individuals who died within 180 days of COVID–19 infection (without a recorded LC diagnosis) into the comparison group. In the second scenario, we restricted the analysis to complete cases with no missing data. Third, we conducted two supplementary analyses using Cohorts III–IV to assess the robustness of our findings under alternative cohort constructions.

All analyses were conducted using the open-source statistical software R (version 4.4.0),^49^ and the ensemble DML models were implemented with the *h2o* package.^50^

## Data availability

All data used in this study is available through the N3C Enclave to approved users. See https://ncats.nih.gov/research/research-activities/n3c/overview for instructions on how to access the data.

## Code availability

All R code used in the analyses is publicly available on Code Ocean (\url{https://codeocean.com/capsule/1739410/tree}) or on GitHub (https://github.com/ChenYW68/EnDML).

## Acknowledgments

Y. Shen would like to acknowledge partial support from NIH grants/contracts R35GM146612. The analyses described in this publication were conducted with data or tools accessed through the NCATS N3C Data Enclave https://covid.cd2h.org and N3C Attribution & Publication Policy v 1.2-2020-08-25b supported by NCATS Contract No. 75N95023D00001, Axle Informatics Subcontract: NCATS-P00438-B. Y. Shen would like to acknowledge support from the Ernest Corn Endowed Professorship at the University of Georgia.

## Author Contributions

**Access to data:** Y. Chen, Z. Chen, B. Li, and J. Liu had full access to all the data in the study and took responsibility for the integrity of the data and the accuracy of the data analysis; **Concept and design:** Y. Chen, K. Ogunyemi, J. Rajbhandari; **Acquisition, analysis, or interpretation of data:** Y. Chen and Z. Chen; **Statistical analysis:** Y. Chen; **Coding:** Y. Chen; **Drafting of the manuscript:** Y. Chen and Y. Ge; **Critical revision of the manuscript for important intellectual content:** Y. Chen, Z. Chen, Y. Ge, B. Li, K. Ogunyemi, J. Liu, F. Luo, Y. Ke, L. Martinez, X. Chen, J. Rajbhandari, and Y. Shen; **Obtained funding:** Y. Shen; **Administrative, technical, or material support:** Y. Shen; **Supervision:** Y. Shen

### Role of the Funder/Sponsor

The funders had no role in the study design, data collection and analysis, decision to publish, or preparation of the manuscript.

## Conflict of Interest

All authors report no potential conflicts of interest.

## Additional information

Supplementary Material provided more details, including descriptions of the datasets and methods, data processing, and additional results.

## Disclaimer

The N3C Publication committee confirmed that this manuscript MSID: 2729.862 is in accordance with N3C data use and attribution policies; however, this content is solely the responsibility of the authors and does not necessarily represent the official views of the National Institutes of Health or the N3C program.

## IRB

The IRB ID of this project is PROJECT00007079, and the Data Use Request (DUR) ID is **DRR-ARHG05H**. The N3C Data Enclave is managed under the authority of the NIH; information can be found at https://ncats.nih.gov/n3c/resources.

## Individual Acknowledgements for Core Contributors

We gratefully acknowledge the following core contributors to N3C: Adam B. Wilcox, Adam M. Lee, Alexis Graves, Alfred (Jerrod) Anzalone, Amin Manna, Amit Saha, Amy Olex, Andrea Zhou, Andrew E. Williams, Andrew M. Southerland, Andrew T. Girvin, Anita Walden, Anjali Sharathkumar, Benjamin Amor, Benjamin Bates, Brian Hendricks, Brijesh Patel, G. Caleb Alexander, Carolyn T. Bramante, Cavin Ward-Caviness, Charisse Madlock-Brown, Christine Suver, Christopher G. Chute, Christopher Dillon, Chunlei Wu, Clare Schmitt, Cliff Takemoto, Dan Housman, Davera Gabriel, David A. Eichmann, Diego Mazzotti, Donald E. Brown, Eilis Boudreau, Elaine L. Hill, Emily Carlson Marti, Emily R. Pfaff, Evan French, Farrukh M Koraishy, Federico Mariona, Fred Prior, George Sokos, Greg Martin, Harold P. Lehmann, Heidi Spratt, Hemalkumar B. Mehta, J.W. Awori Hayanga, Jami Pincavitch, Jaylyn Clark, Jeremy Richard Harper, Jessica Yasmine Islam, Jin Ge, Joel Gagnier, Johanna J. Loomba, John B. Buse, Jomol Mathew, Joni L. Rutter, Julie A. McMurry, Justin Guinney, Justin Starren, Karen Crowley, Katie Rebecca Bradwell, Kellie M. Walters, Ken Wilkins, Kenneth R. Gersing, Kenrick Cato, Kimberly Murray, Kristin Kostka, Lavance Northington, Lee Pyles, Lesley Cottrell, Lili M. Portilla, Mariam Deacy, Mark M. Bissell, Marshall Clark, Mary Emmett, Matvey B. Palchuk, Melissa A. Haendel, Meredith Adams, Meredith Temple-O’Connor, Michael G. Kurilla, Michele Morris, Nasia Safdar, Nicole Garbarini, Noha Sharafeldin, Ofer Sadan, Patricia A. Francis, Penny Wung Burgoon, Philip R.O. Payne, Randeep Jawa, Rebecca Erwin-Cohen, Rena C. Patel, Richard A. Moffitt, Richard L. Zhu, Rishikesan Kamaleswaran, Robert Hurley, Robert T. Miller, Saiju Pyarajan, Sam G. Michael, Samuel Bozzette, Sandeep K. Mallipattu, Satyanarayana Vedula, Scott Chapman, Shawn T. O’Neil, Soko Setoguchi, Stephanie S. Hong, Steven G. Johnson, Tellen D. Bennett, Tiffany J. Callahan, Umit Topaloglu, Valery Gordon, Vignesh Subbian, Warren A. Kibbe, Wenndy Hernandez, Will Beasley, Will Cooper, William Hillegass, Xiaohan Tanner Zhang. Details of contributions available at covid.cd2h.org/core-contributors

The authors would like to thank JiaLi Zhu (St John’s College Cardiff, UK) for constructive discussions.

## Data Partners with Released Data

The following institutions whose data is released or pending:

Available: Advocate Health Care Network — UL1TR002389: The Institute for Translational Medicine (ITM) • Aurora Health Care Inc — UL1TR002373: Wisconsin Network For Health Research • Boston University Medical Campus — UL1TR001430: Boston University Clinical and Translational Science Institute • Brown University — U54GM115677: Advance Clinical Translational Research (Advance-CTR) • Carilion Clinic — UL1TR003015: iTHRIV Integrated Translational health Research Institute of Virginia • Case Western Reserve University — UL1TR002548: The Clinical& Translational Science Collaborative of Cleveland (CTSC) • Charleston Area Medical Center — U54GM104942: West Virginia Clinical and Translational Science Institute (WVCTSI) • Children’s Hospital Colorado — UL1TR002535: Colorado Clinical and Translational Sciences Institute • Columbia University Irving Medical Center — UL1TR001873: Irving Institute for Clinical and Translational Research • Dartmouth College — None (Voluntary) Duke University — UL1TR002553: Duke Clinical and Translational Science Institute • George Washington Children’s Research Institute — UL1TR001876: Clinical and Translational Science Institute at Children’s National (CTSA-CN) • George Washington University — UL1TR001876: Clinical and Translational Science Institute at Children’s National (CTSA-CN) • Harvard Medical School — UL1TR002541: Harvard Catalyst • Indiana University School of Medicine — UL1TR002529: Indiana Clinical and Translational Science Institute • Johns Hopkins University — UL1TR003098: Johns Hopkins Institute for Clinical and Translational Research • Louisiana Public Health Institute — None (Voluntary) • Loyola Medicine — Loyola University Medical Center • Loyola University Medical Center — UL1TR002389: The Institute for Translational Medicine (ITM) • Maine Medical Center — U54GM115516: Northern New England Clinical & Translational Research (NNE-CTR) Network • Mary Hitchcock Memorial Hospital & Dartmouth Hitchcock Clinic — None (Voluntary) • Massachusetts General Brigham — UL1TR002541: Harvard Catalyst • Mayo Clinic Rochester — UL1TR002377: Mayo Clinic Center for Clinical and Translational Science (CCaTS) • Medical University of South Carolina

— UL1TR001450: South Carolina Clinical & Translational Research Institute (SCTR) • MITRE Corporation — None (Voluntary) • Montefiore Medical Center — UL1TR002556: Institute for Clinical and Translational Research at Einstein and Montefiore • Nemours — U54GM104941: Delaware CTR ACCEL Program • NorthShore University HealthSystem — UL1TR002389: The Institute for Translational Medicine (ITM) • Northwestern University at Chicago — UL1TR001422: Northwestern University Clinical and Translational Science Institute (NUCATS) • OCHIN — INV-018455: Bill and Melinda Gates Foundation grant to Sage Bionetworks • Oregon Health & Science University — UL1TR002369: Oregon Clinical and Translational Research Institute • Penn State Health Milton S. Hershey Medical Center — UL1TR002014: Penn State Clinical and Translational Science Institute • Rush University Medical Center — UL1TR002389: The Institute for Translational Medicine (ITM) • Rutgers, The State University of New Jersey — UL1TR003017: New Jersey Alliance for Clinical and Translational Science • Stony Brook University — U24TR002306 • The Alliance at the University of Puerto Rico, Medical Sciences Campus — U54GM133807: Hispanic Alliance for Clinical and Translational Research (The Alliance) • The Ohio State University — UL1TR002733: Center for Clinical and Translational Science • The State University of New York at Buffalo — UL1TR001412: Clinical and Translational Science Institute • The University of Chicago — UL1TR002389: The Institute for Translational Medicine (ITM) • The University of Iowa — UL1TR002537: Institute for Clinical and Translational Science • The University of Miami Leonard M. Miller School of Medicine — UL1TR002736: University of Miami Clinical and Translational Science Institute • The University of Michigan at Ann Arbor — UL1TR002240: Michigan Institute for Clinical and Health Research • The University of Texas Health Science Center at Houston — UL1TR003167: Center for Clinical and Translational Sciences (CCTS) • The University of Texas Medical Branch at Galveston — UL1TR001439: The Institute for Translational Sciences • The University of Utah — UL1TR002538: Uhealth Center for Clinical and Translational Science • Tufts Medical Center — UL1TR002544: Tufts Clinical and Translational Science Institute • Tulane University — UL1TR003096: Center for Clinical and Translational Science • The Queens Medical Center — None (Voluntary) • University Medical Center New Orleans — U54GM104940: Louisiana Clinical and Translational Science (LA CaTS) Center • University of Alabama at Birmingham — UL1TR003096: Center for Clinical and Translational Science • University of Arkansas for Medical Sciences — UL1TR003107: UAMS Translational Research Institute • University of Cincinnati — UL1TR001425: Center for Clinical and Translational Science and Training • University of Colorado Denver, Anschutz Medical Campus — UL1TR002535: Colorado Clinical and Translational Sciences Institute • University of Illinois at Chicago — UL1TR002003:

UIC Center for Clinical and Translational Science • University of Kansas Medical Center — UL1TR002366: Frontiers: University of Kansas Clinical and Translational Science Institute • University of Kentucky — UL1TR001998: UK Center for Clinical and Translational Science • University of Massachusetts Medical School Worcester — UL1TR001453: The UMass Center for Clinical and Translational Science (UMCCTS) • University Medical Center of Southern Nevada — None (voluntary) • University of Minnesota — UL1TR002494: Clinical and Translational Science Institute • University of Mississippi Medical Center — U54GM115428: Mississippi Center for Clinical and Translational Research (CCTR) • University of Nebraska

Medical Center — U54GM115458: Great Plains IDeA-Clinical & Translational Research • University of North Carolina at Chapel Hill — UL1TR002489: North Carolina Translational and Clinical Science Institute • University of Oklahoma Health Sciences Center — U54GM104938: Oklahoma Clinical and Translational Science Institute (OCTSI) • University of Pittsburgh — UL1TR001857: The Clinical and Translational Science Institute (CTSI) • University of Pennsylvania — UL1TR001878: Institute for Translational Medicine and Therapeutics • University of Rochester — UL1TR002001: UR Clinical & Translational Science Institute • University of Southern California — UL1TR001855: The Southern California Clinical and Translational Science Institute (SC CTSI) • University of Vermont — U54GM115516: Northern New England Clinical & Translational Research (NNE-CTR) Network • University of Virginia — UL1TR003015: iTHRIV Integrated Translational health Research Institute of Virginia • University of Washington — UL1TR002319: Institute of Translational Health Sciences • University of Wisconsin-Madison — UL1TR002373: UW Institute for Clinical and Translational Research • Vanderbilt University Medical Center — UL1TR002243: Vanderbilt Institute for Clinical and Translational Research • Virginia Commonwealth University — UL1TR002649: C. Kenneth and Dianne Wright Center for Clinical and Translational Research • Wake Forest University Health Sciences — UL1TR001420: Wake Forest Clinical and Translational Science Institute • Washington University in St. Louis — UL1TR002345: Institute of Clinical and Translational Sciences • Weill Medical College of Cornell University — UL1TR002384: Weill Cornell Medicine Clinical and Translational Science Center • West Virginia University — U54GM104942: West Virginia Clinical and Translational Science Institute (WVCTSI) • Submitted: Icahn School of Medicine at Mount Sinai — UL1TR001433: ConduITS Institute for Translational Sciences • The University of Texas Health Science Center at Tyler — UL1TR003167: Center for Clinical and Translational Sciences (CCTS) • University of California, Davis — UL1TR001860: UCDavis Health Clinical and Translational Science Center • University of California, Irvine — UL1TR001414: The UC Irvine Institute for Clinical and Translational Science (ICTS) • University of California, Los Angeles — UL1TR001881: UCLA Clinical Translational Science Institute • University of California, San Diego — UL1TR001442: Altman Clinical and Translational Research Institute • University of California, San Francisco — UL1TR001872: UCSF Clinical and Translational Science Institute NYU Langone Health Clinical Science Core, Data Resource Core, and PASC Biorepository Core — OTA-21-015A: Post-Acute Sequelae of SARS-CoV-2 Infection Initiative (RECOVER) Pending: Arkansas Children’s Hospital — UL1TR003107: UAMS Translational Research Institute • Baylor College of Medicine — None (Voluntary) • Children’s Hospital of Philadelphia — UL1TR001878: Institute for Translational Medicine and Therapeutics • Cincinnati Children’s Hospital Medical Center — UL1TR001425: Center for Clinical and Translational Science and Training • Emory University — UL1TR002378: Georgia Clinical and Translational Science Alliance • HonorHealth — None (Voluntary) • Loyola University Chicago — UL1TR002389: The Institute for Translational Medicine (ITM) • Medical College of Wisconsin — UL1TR001436: Clinical and Translational Science Institute of Southeast Wisconsin • MedStar Health Research Institute — None (Voluntary) • Georgetown University — UL1TR001409: The Georgetown-Howard Universities Center for Clinical and Translational Science (GHUCCTS) • MetroHealth — None (Voluntary) • Montana State University — U54GM115371: American Indian/Alaska Native CTR • NYU Langone Medical Center — UL1TR001445: Langone Health’s Clinical and Translational Science Institute • Ochsner Medical Center — U54GM104940: Louisiana Clinical and Translational Science (LA CaTS) Center • Regenstrief Institute — UL1TR002529: Indiana Clinical and Translational Science Institute • Sanford Research — None (Voluntary) • Stanford University — UL1TR003142: Spectrum: The Stanford Center for Clinical and Translational Research and Education • The Rockefeller University — UL1TR001866: Center for Clinical and Translational Science • The Scripps Research Institute — UL1TR002550: Scripps Research Translational Institute • University of Florida — UL1TR001427: UF Clinical and Translational Science Institute • University of New Mexico Health Sciences Center — UL1TR001449: University of New Mexico Clinical and Translational Science Center • University of Texas Health Science Center at San Antonio — UL1TR002645: Institute for Integration of Medicine and Science • Yale New Haven Hospital — UL1TR001863: Yale Center for Clinical Investigation.

## Notes

### Competing Interest Statement

The authors have declared no competing interest.

### Author Declarations

Ethics approval Access to the N3C Data Enclave analytics platform for research purposes was granted to the University of Georgia, with institutional review board (IRB) identification number: PROJECT00007079 and data use request (DUR) identification number: DRR-ARHG05H

### Summary of Updates

Conducted extensive additional numerical analyses by introducing two new cohorts to facilitate more comprehensive comparisons across different comorbidity exposure-window definitions.

## Reference

1. Al-Aly Z, Davis H, McCorkell L, et al. Long COVID science, research and policy. Nat Med. 2024;30(8):2148–2164.

2. Centers for Disease Control and Prevention. Long COVID Basics. 2025. https://www.cdc.gov/long-covid/about/index.html

3. Chen Z, Li B, Chen Y, et al. Mapping spatial and social inequities of long COVID across the United States: a retrospective cohort study. Lancet Reg Heal. 2026;56:101401.

4. Fang Z, Ahrnsbrak R, Rekito A. Evidence mounts that about 7% of US adults have had long COVID. JAMA. 2024;332(1):5–6.

5. Nehme M, Braillard O, Chappuis F, Guessous I. The chronification of post-COVID condition associated with neurocognitive symptoms, functional impairment and increased healthcare utilization. Sci Rep. 2022;12(1):14505.

6. Gottlieb M, Spatz ES, Yu H, et al. Effects of long COVID on healthcare utilization. Plos One. 2025;20(7):e0327218.

7. Subramanian A, Nirantharakumar K, Hughes S, et al. Symptoms and risk factors for long COVID in non-hospitalized adults. Nat Med. 2022;28(8):1706–1714.

8. Berentschot JC, Bek LM, Drost M, et al. Health outcomes up to 3 years and post-exertional malaise in patients after hospitalization for COVID-19: a multicentre prospective cohort study (CO-FLOW). Lancet Reg Heal. 2025;53.

9. Jacobs ET, Catalfamo CJ, Colombo PM, et al. Pre-existing conditions associated with post-acute sequelae of COVID-19. J Autoimmun. 2023;135:102991. doi:10.1016/j.jaut.2022.102991

10. Greißel A, Schneider A, Donnachie E, Gerlach R, Tauscher M, Hapfelmeier A. Impact of pre-existing mental health diagnoses on development of post-COVID and related symptoms: a claims data-based cohort study. Sci Rep. 2024;14(1):2408.

11. Samuels TA, Cohen D, Brancati FL, Coresh J, Kao WHL. Delayed diagnosis of incident type 2 diabetes mellitus in the ARIC study. Am J Manag Care. 2006;12(12):717–724.

12. Whitfield E, White B, Denaxas S, Lyratzopoulos G. Diagnostic windows in non-neoplastic diseases: a systematic review. Br J Gen Pract. Published online 2023.

13. Institute of Medicine (US) Committee on Health Insurance Status and Its Consequences. Coverage Matters. In: America’s Uninsured Crisis: Consequences for Health and Health Care. National Academies Press; 2009. https://www.ncbi.nlm.nih.gov/books/NBK214963/

14. Tartof SY, Malden DE, Liu ILA, et al. Health Care Utilization in the 6 Months Following SARS-CoV-2 Infection. JAMA Netw Open. 2022;5(8):e2225657. doi:10.1001/jamanetworkopen.2022.25657

15. Hernandez-Romieu AC, Leung S, Mbanya A, et al. Health Care Utilization and Clinical Characteristics of Nonhospitalized Adults in an Integrated Health Care System 28–180 Days After COVID-19 Diagnosis—Georgia, May 2020–March 2021. MMWR Morb Mortal Wkly Rep. 2021;70(17):644–650. doi:10.15585/mmwr.mm7017e3

16. Respiratory Therapy. Repeat COVID Infections Behind Most Long COVID Cases — Four-Year Study Finds. Published online 2025. https://respiratory-therapy.com/disorders-diseases/infectious-diseases/coronavirus/repeat-covid-infections-most-long-covid-cases-four-year-study-finds/

17. Haendel MA, Chute CG, Bennett TD, et al. The National COVID Cohort Collaborative (N3C): rationale, design, infrastructure, and deployment. J Am Med Inform Assoc. 2021;28(3):427–443.

18. Hadley E, Yoo YJ, Patel S, et al. Insights from an N3C RECOVER EHR-based cohort study characterizing SARS-CoV-2 reinfections and Long COVID. Commun Med. 2024;4(1):129.

19. USDA. Rural-Urban Continuum Codes (2023). U.S. Department of Agriculture. https://www.ers.usda.gov/data-products/rural-urban-continuum-codes

20. CHR Roadmaps. County Health Rankings (2025). University of Wisconsin Population Health Institute. https://www.countyhealthrankings.org/

21. Flanagan BE, Hallisey EJ, Adams E, Lavery A. Measuring community vulnerability to natural and anthropogenic hazards: the Centers for Disease Control and Prevention’s Social Vulnerability Index. J Environ Health. 2018;80(10):34.

22. Chernozhukov V, Chetverikov D, Demirer M, Duflo E, Hansen C, Newey W. Double/debiased/neyman machine learning of treatment effects. Am Econ Rev. 2017;107(5):261–265.

23. Chernozhukov V, Chetverikov D, Demirer M, et al. Double/debiased machine learning for treatment and structural parameters. Econom J. 2018;21(1):C1–C68. doi:10.1111/ectj.12097

24. Moccia C, Moirano G, Popovic M, et al. Machine learning in causal inference for epidemiology. Eur J Epidemiol. 2024;39(10):1097–1108. doi:10.1007/s10654-024-01173-x

25. Prasad V, Makary MA. An evidence-based approach to Covid-19 vaccination. N Engl J Med. 2025;392(24):2484–2486.

26. Ogunyemi K, Shen Y, Li B, et al. Long COVID Burden Attributable to Preexisting Comorbidity: A Retrospective Cohort Study. SSRN Electron J. Published online 2026. doi:10.2139/ssrn.6763417

27. Wu Q, Ailshire JA, Crimmins EM. Long COVID and symptom trajectory in a representative sample of Americans in the first year of the pandemic. Sci Rep. 2022;12(1):11647.

28. Wee LE, Lim JT, Tay AT, et al. Autoimmune sequelae after delta or omicron variant SARS-CoV-2 infection in a highly vaccinated cohort. JAMA Netw Open. 2024;7(8):e2430983–e2430983.

29. Byambasuren O, Stehlik P, Clark J, Alcorn K, Glasziou P. Effect of covid-19 vaccination on long covid: systematic review. BMJ Med. 2023;2(1):e000385.

30. Wells CR, Galvani AP. Tackling the politicisation of COVID-19 data reporting through open access data sharing. Lancet Infect Dis. 2022;22(12):1660.

31. Louie P, Wu C. Race, socioeconomic status, and long COVID. Soc Curr. 2024;11(3):203–215.

32. Li B, Ke Y, Chen X, Martinez L, Shen Y. Robust COVID-19 Mortality Risk Assessment: Validation of a Two-Step Algorithm From the National COVID Cohort Collaborative. J Infect Dis. 2026;233(2):351–362.

33. Chen Y, Chang X, Luo F, Huang H. Additive dynamic models for correcting numerical model outputs. Comput Stat Data Anal. 2023;187:107799. doi:10.1016/j.csda.2023.107799

34. Chen Y, Chang X, Zhang B, Huang H. Efficient and effective calibration of numerical model outputs using hierarchical dynamic models. Ann Appl Stat. 2024;18(2). doi:10.1214/23-AOAS1823

35. Li Y, Kong Y, Ebell MH, et al. Development and validation of a two-step predictive risk stratification model for coronavirus disease 2019 in-hospital mortality: a multicenter retrospective cohort study. Front Med. 2022;9:827261.

36. Bergquist T, Loomba J, Pfaff E, et al. Crowd-sourced machine learning prediction of long COVID using data from the National COVID Cohort Collaborative. EBioMedicine. 2024;108.

37. National Academies of Sciences, Engineering, and Medicine. A Long COVID Definition: A Chronic, Systemic Disease State with Profound Consequences. National Academies Press; 2024. doi:10.17226/27696

38. World Health Organization. The top 10 causes of death. Published online 2024. https://www.who.int/news-room/fact-sheets/detail/the-top-10-causes-of-death

39. Van Buuren S, Groothuis-Oudshoorn K. mice: Multivariate imputation by chained equations in R. J Stat Softw. 2011;45:1–67.

40. Yu W, Li S, Ye T, Xu R, Song J, Guo Y. Deep ensemble machine learning framework for the estimation of PM 2.5 concentrations. Environ Health Perspect. 2022;130(3):037004.

41. Imbens GW. Causal inference in the social sciences. Annu Rev Stat Its Appl. 2024;11.

42. Rane N, Choudhary SP, Rane J. Ensemble deep learning and machine learning: applications, opportunities, challenges, and future directions. Stud Med Health Sci. 2024;1(2):18–41.

43. Chen Y, Luo F, Martinez L, Jiang S, Shen Y. Identifying key aspects to enhance predictive modeling for early identification of schistosomiasis hotspots to guide mass drug administration. PLoS Negl Trop Dis. 2025;19(7):e0013315.

44. Hailpern SM, Visintainer PF. Odds ratios and logistic regression: further examples of their use and interpretation. Stata J. 2003;3(3):213–225.

45. Snowden JM, Rose S, Mortimer KM. Implementation of G-computation on a simulated data set: demonstration of a causal inference technique. Am J Epidemiol. 2011;173(7):731–738.

46. Semenova V, Chernozhukov V. Debiased machine learning of conditional average treatment effects and other causal functions. Econom J. 2021;24(2):264–289.

47. VanderWeele TJ, Ding P. Sensitivity analysis in observational research: introducing the E-value. Ann Intern Med. 2017;167(4):268–274.

48. Chung WT, Chung KC. The use of the E-value for sensitivity analysis. J Clin Epidemiol. 2023;163:92–94.

49. R Core Team. R: A Language and Environment for Statistical Computing (2025). R Foundation for Statistical Computing. https://www.R-project.org/

50. Candel A, Parmar V, LeDell E, Arora A. Deep learning with H2O. H2O Ai Inc. Published online 2016:1-21.

