## Supplementary Materials for "Comorbidity Exposure-Window Definitions and Multidimensional Disparities in Long COVID Risk: Evidence from a U.S. National Cohort (2020–2024)"

This document provides detailed processing procedures of N3C data, construction of two cohorts, descriptions of the datasets and methods, data exploration analysis, and additional results.

Data were extracted from version 185 of both the Logic Liaison COVID-19 Cases and Controls Fact Day Table and the Condition Occurrence table to establish acute COVID-19 and LC cohorts. Acute COVID-19 was defined by ICD-10 code U07.1, a positive PCR, or antibody test, prioritizing the diagnosis code. The LC cohort was identified via codes U09.9, or B94.8, excluding records before January 1, 2020, and removing daily duplicates. For both groups, only the primary infection or diagnosis date was retained for analysis. The cohorts were then combined using an outer join to capture all individuals with acute infection, LC, or both.

Following the merge, records with missing or incomplete ZIP codes were excluded, and valid five-digit ZIP codes were mapped to counties using geographic center points. Mortality records and clinical covariates, including hospitalization, urban or rural status, and comorbidity diagnosis dates, were then integrated. Finally, we excluded records without a recorded infection date, as well as any records where the primary infection or LC diagnosis occurred before March 1, 2020. This process yielded a final analytic sample of 6,130,413 unique individuals, comprising 6,041,827 non-LC and 88,586 LC patients (**Figure** S1).

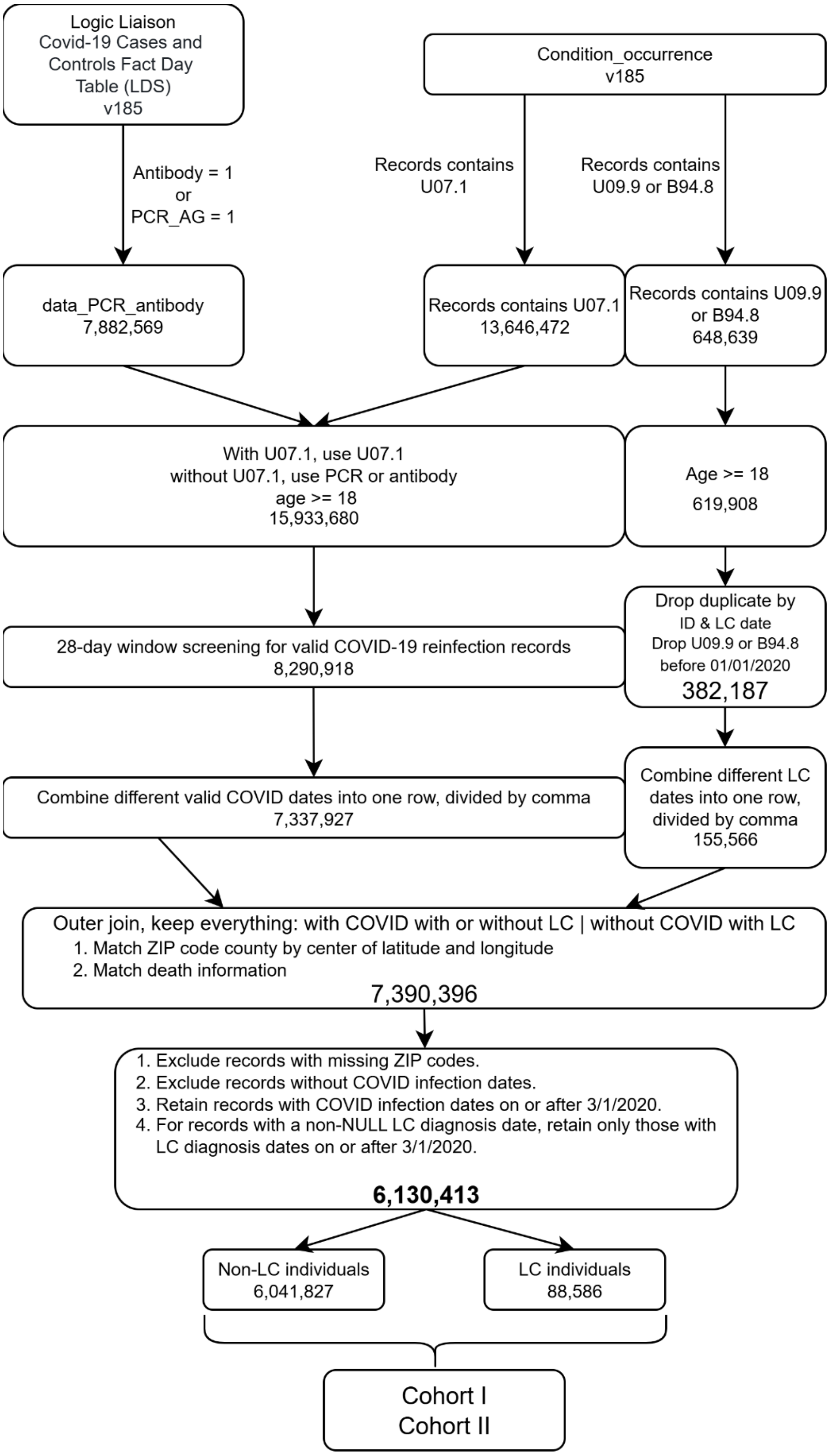


Figure S1: Data processing workflow.

### S1 Cohort developments

As illustrated in the main text, for individuals with multiple records, the earliest qualifying dates for COVID-19 infection and LC diagnosis were used. The longest observed interval between COVID-19 infection and LC diagnosis exceeded 4 years. This extended interval is likely due to multiple COVID-19 infections or delayed clinical recognition of LC (Respiratory Therapy, 2025). Moreover, the proportion of such cases is often substantial rather than negligible (**Table** S1). These practical considerations underscore the importance of incorporating cases with comorbidities occurring between COVID-19 infection and LC diagnosis when assessing comorbidity-associated risks of LC. Excluding these cases could lead to biased LC risk estimates.

Based on the processed data shown in **Figure** S1, we restricted the study population to individuals with available COVID-19 diagnosis dates. Within this population, we constructed four cohorts (I–IV), with Cohorts I, III, and IV representing scenarios involving pre-infection comorbidities and Cohort II representing a scenario involving pre-LC comorbidities.

#### S1.1 Construction of cohorts

In the construction of cohort, we used the $c$th comorbidity as an illustration to define exposure status.

**Overview**: Cohort I used the COVID-19 diagnosis date as a common cutoff to define exposure status for individuals with and without LC, an approach widely employed in previous studies (Subramanian et al., 2022; Jacobs et al., 2023; Berentschot et al., 2025). However, this approach may have two key limitations: (1) exposure misclassification for the $c$th comorbidity documented between the COVID-19 diagnosis date and the LC diagnosis date; and (2) failure to account for the minimum temporal interval between documentation of the $c$th comorbidity and LC diagnosis. In particular, some intervals recorded in N3C may be even shorter than 28 days, a threshold previously used by the CDC to define post-COVID conditions (Reese et al., 2023). Such short intervals may be insufficient to establish the temporal precedence necessary to consider the $c$th comorbidity a potential risk factor for developing LC. To address these limitations, Cohorts II–IV incorporated a minimum interval of 90 days between comorbidity documentation and LC diagnosis. However, Cohort III did not address exposure misclassification, whereas Cohort IV introduced additional exclusion criteria to mitigate this issue, at the cost of reducing the study population relative to Cohorts I–III.

**Cohort I: Pre-Infection comorbidities defined by the COVID-19 diagnosis date without the minimum temporal interval.** Cohort I was constructed to investigate the associations between comorbidities documented before COVID-19 infection and the subsequent development of LC. The exposure status for the $c$th comorbidity was defined as follows:

(a) Individuals with or without LC were considered to be exposed to the $c$th comorbidity ($X_{i,c} = 1$) if the initial diagnosis date of that comorbidity occurred before the COVID-19 diagnosis date (denoted by COVID). Otherwise, they were classified as unexposed to the $c$th comorbidity ($X_{i,c} = 0$).

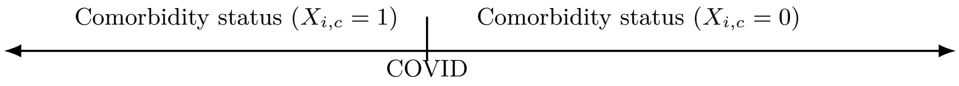

**Cohort II: Pre-LC comorbidities before LC diagnosis with the minimum temporal interval.** Cohort II was constructed to investigate the associations between comorbidities documented prior to LC diagnosis and the development of LC. The exposure status for the $c$th comorbidity was defined as follows:

(a) Individuals with LC were considered to be exposed to the $c$th comorbidity ($X_{i,c} = 1$) if the initial diagnosis date of that comorbidity occurred before 90 days prior to the LC diagnosis date (LC − 90d) when 180 days after the COVID-19 diagnosis date (COVID + 180d) occurred later than (LC − 90d). Otherwise, they were classified as unexposed to the $c$th comorbidity ($X_{i,c} = 0$).

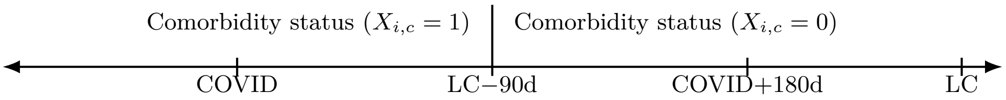


(b) Individuals with LC were considered to be exposed to the $c$th comorbidity ($X_{i,c} = 1$) if the initial diagnosis date of that comorbidity occurred before (COVID + 180d) when (LC − 90d) occurred later than (COVID + 180d). Otherwise, they were classified as unexposed to the $c$th comorbidity ($X_{i,c} = 0$).

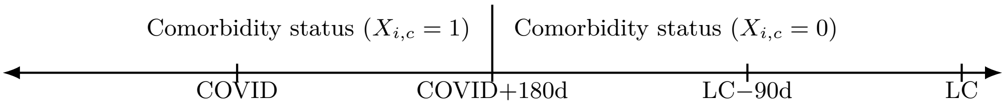


(c) Individuals without LC were considered to be exposed to the $c$th comorbidity ($X_{i,c} = 1$) if the initial diagnosis date of that comorbidity occurred before (COVID + 180d). Otherwise, they were classified as unexposed to the $c$th comorbidity ($X_{i,c} = 0$).

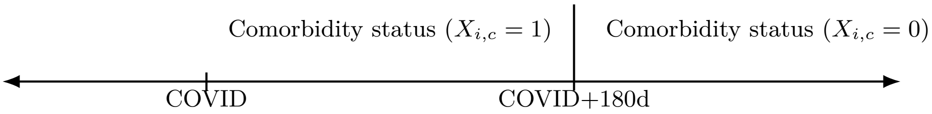

**Cohort III: Pre-Infection comorbidities with the minimum temporal interval.** Cohort III was constructed to investigate the associations between preexisting comorbidities prior to COVID and subsequent LC development, using the same minimum temporal interval as Cohort II. The exposure status for the $c$th comorbidity was defined as follows:

(a) Individuals with LC were considered to be exposed to the $c$th comorbidity ($X_{i,c} = 1$) if the initial diagnosis date of that comorbidity occurred before the earlier of COVID and (LC − 90d), including the following Cases 1-2. Otherwise, they were classified as unexposed to the $c$th comorbidity ($X_{i,c} = 0$).

**Case 1**

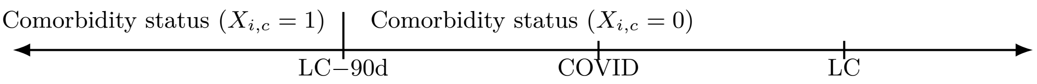


**Case 2**

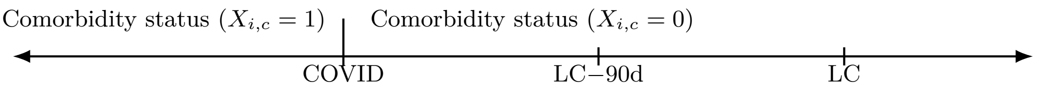


(b) Individuals without LC were considered to be exposed to the $c$th comorbidity ($X_{i,c} = 1$) if the initial diagnosis date of that comorbidity occurred before COVID. Otherwise, they were classified as unexposed to the $c$th comorbidity ($X_{i,c} = 0$).

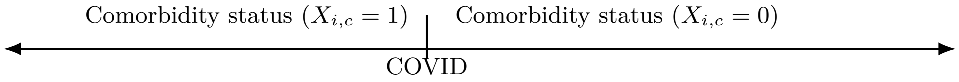


(c) Among individuals either with or without LC, those with no documented occurrence of the $c$th comorbidity were classified as unexposed to the $c$th comorbidity $X_{i,c} = 0$.

**Cohort IV: Pre-Infection comorbidities before COVID-19 with the minimum temporal interval while excluding some individuals.** Cohort IV was also constructed to investigate the associations between comorbidities documented before COVID and subsequent LC development. It adopted the same minimum temporal interval as Cohort II-III and applied additional exclusion criteria to mitigate exposure misclassification. The exposure status for the $c$th comorbidity was defined as follows:

(a) Individuals with LC were considered to be exposed to the $c$th comorbidity ($X_{i,c} = 1$) if the initial diagnosis date of that comorbidity occurred before the earlier of COVID and (LC − 90d), including the following Cases 1-2.

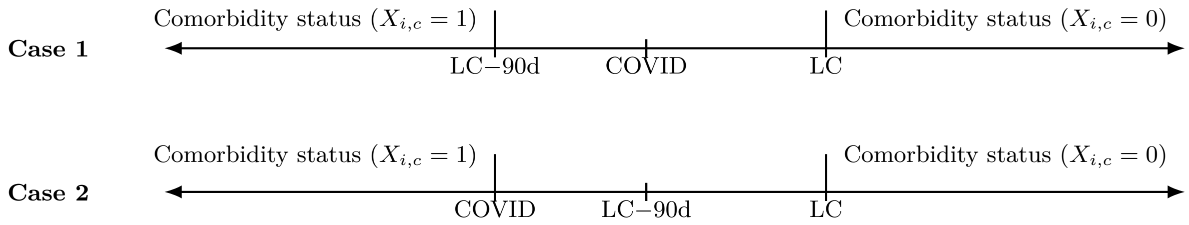


(b) Individuals without LC were considered to be exposed to the $c$th comorbidity ($X_{i,c} = 1$) if the initial diagnosis date of that comorbidity occurred before COVID.

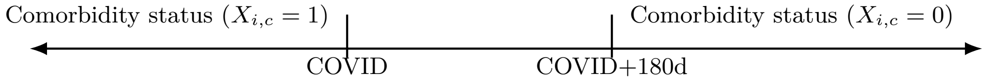


(c) Individuals with LC were excluded from the analysis of the $c$th comorbidity if the initial diagnosis date of that comorbidity occurred between the earlier of COVID and (LC − 90d), and the LC diagnosis date, including the following Cases 1-2:

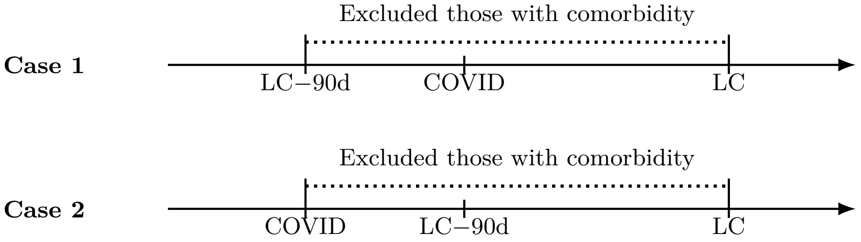


(d) Individuals without LC were excluded from the analysis of the $c$th comorbidity if the initial diagnosis date of that comorbidity occurred between COVID and (COVID + 180d).

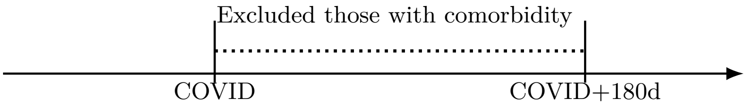


(e) Individuals without LC whose first documentation of the $c$th comorbidity occurred more than (COVID + 180d) were retained and classified as unexposed to the $c$th comorbidity ($X_{i,c} = 0$).

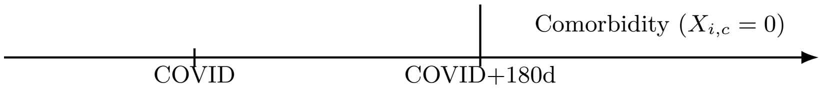


(f) Among individuals either with or without LC, those with no documented occurrence of the $c$th comorbidity were retained and classified as unexposed to the $c$th comorbidity ($X_{i,c} = 0$).

We provided explanations for the rationale underlying criteria (a)–(e) to clarify the relatively complex scenarios in Cohort IV. Some of these explanations also apply to Cohorts II and III. **Criterion (a)** ensured sufficient temporal separation between the $c$th comorbidity and the subsequent LC diagnosis, because a comorbidity documented shortly before the LC diagnosis may not have preceded LC by a sufficient period to be meaningfully evaluated as a potential risk factor for developing LC. **Criterion (b)** defined (COVID + 180d) as the LC risk window, as our previous study showed that most individuals with LC were diagnosed within 180 days after the initial infection (Chen et al., 2026). **Criterion (c)** was introduced to mitigate impacts of potential exposure misclassification in Cohorts I and III on LC risk estimates, given that delayed diagnosis is common across chronic diseases (Samuels et al., 2006; Whitfield et al., 2023), particularly among individuals who are uninsured or have limited access to healthcare (Institute of Medicine (US) Committee on Health Insurance Status and Its Consequences, 2009). Similarly, **Criterion (d)** was also set due to potential exposure misclassification. Therefore, these individuals were removed from Cohort IV. **Criterion (e)** was set to maximize the use of available data. Collectively, these criteria establish temporal precedence, clearly distinguish preexisting from subsequently documented comorbidities, reduce potential exposure misclassification, and make full use of the available data.

Under the definition of Cohort IV, we retained individuals whose $c$th comorbidity was diagnosed after the LC diagnosis date under Criterion (a) or more than (COVID + 180d) under Criterion (e), rather than excluding them like Criteria (c) or (d). This was established based on two considerations. First, for individuals whose $c$th comorbidity was first documented after the LC diagnosis date, the documentation date was less likely to reflect a condition that had already been recognized before LC diagnosis, as LC diagnosis generally requires that the symptoms cannot be explained by other existing conditions (Soriano et al., 2022). Therefore, the $c$th comorbidity occurred after the LC diagnosis date could not have influenced LC onset. Second, the risk of developing LC attributable to the initial infection may be relatively low beyond (COVID + 180d), as most individuals with LC were diagnosed within the 180-day LC assessment period (Chen et al., 2026).

**Comparisons between Cohort II and IV**: Individuals **with** LC whose $c$th comorbidity was first documented more than (COVID + 180d), these individuals were retained in Cohort II under Criteria (a)–(b). However, they may or may not have been retained in Cohort IV under criterion (c), depending on whether (COVID + 180d) was later than the LC diagnosis date. If so, these individuals were retained under Criterion (a); otherwise, they were excluded under Criterion (c).

In Cohort II, a joint cutoff date, defined as the earlier of (LC − 90d) and (COVID + 180d), was applied under Criteria (a)–(b). However, this cutoff date was not applicable to Cohort I, which was designed to assess pre-infection comorbidities, with the COVID-19 diagnosis date serving as the key temporal reference point. Therefore, the other joint cutoff date for Cohort I was defined as the earlier of (LC − 90d) and COVID; see Criterion (a).

#### S1.2 Comorbidity-specific relationships across cohorts

From the perspective of cohort construction, cohorts defined based on pre-infection comorbidities are intended to capture baseline susceptibility before infection, whereas Cohort II captures a distinct clinical construct: prediagnostic vulnerability. Therefore, if the primary research question is to assess the effect of baseline susceptibility on the risk of LC, Cohorts I, III, and IV are directly aligned with that objective. However, if the primary research question is to assess the effect of broader prediagnostic vulnerability on the risk of LC, these cohorts may not fully capture the relevant comorbidity burden, and Cohort II may be more appropriate.

In this study, **we focus on accurately assessing the broader prediagnostic vulnerability present before LC diagnosis, with the aim of determining whether pre-infection comorbidities alone are sufficient to characterize the risk of developing LC**. In previous studies, pre-infection comorbidities have commonly been used to represent preexisting comorbidities, while conditions first documented between COVID-19 infection and LC diagnosis are often not captured, as in Cohorts I, III, and IV (Subramanian et al., 2022; Jacobs et al., 2023; Berentschot et al., 2025). In this context, the definition of preexisting comorbidities may be somewhat ambiguous, as it may refer either to comorbidities documented before COVID-19 infection or to those documented before LC diagnosis. To clarify this distinction, we explicitly differentiate between pre-infection comorbidities and pre-LC comorbidities in this work.

As discussed in the Discussion section of the main text, Cohorts I and III–IV may artificially amplify the contrast between the exposed and unexposed groups for preexisting comorbidities with prolonged disease courses, thereby leading to overestimation of the relative effect sizes; also see Supplementary Section S1.3. In particular, this overestimation may be more pronounced for comorbidities characterized by prolonged disease courses that are more susceptible to delayed clinical recognition or underascertainment. Following acute COVID-19 infection, many individuals can have increased contact with the healthcare system and greater opportunities for clinical evaluation than they had before the pandemic (Tartof et al., 2022; Hernandez-Romieu et al., 2021). As a result, an underlying condition that was already present before infection but had been underascertained is likely to be diagnosed after COVID-19 infection and before LC diagnosis. Under this scenario, Cohort II, designed to capture prediagnostic vulnerability, may identify individuals with preexisting comorbidities that were present but remained undiagnosed prior to infection.

#### S1.3 Effect size analysis

Let the $c$th chronic condition be the target comorbidity, which serves as the exposure of interest. The exposed group consists of individuals with the target comorbidity prior to LC, whereas the unexposed group consists of individuals without it. Define $\mathrm{R}_0 = \frac{a_{01}}{a_{00}+a_{01}}$, $\mathrm{R}_1 = \frac{a_{11}}{a_{10}+a_{11}}$, where $a_{00}$ and $a_{01}$ denote the numbers of individuals without and with LC, respectively, in the unexposed group, and $a_{10}$ and $a_{11}$ denote the corresponding numbers in the exposed group.

The following mechanisms may jointly contribute to the larger effect estimates observed in Cohorts I and III–IV than in Cohort II.

(i) Compared with Cohort II, Cohort IV excluded some individuals from the exposed group, whereas Cohorts I and III may have misclassified some truly exposed individuals as unexposed (see the cohort construction procedures above). These exclusions and misclassifications reduced the denominator $a_{10} + a_{11}$ of $\mathrm{R}_1$. In practice, LC incidence in the study population was generally below 15% (Chen et al., 2026). Accordingly, the reduction in $a_{10}$ associated with these exclusions or misclassifications was generally larger than that in $a_{11}$, as shown in Table S1. The removal of these individuals from the exposed group would lead to an overestimation of $\mathrm{R}_1$ in Cohorts I and III–IV. In contrast, estimates of $\mathrm{R}_0$ may be relatively robust to these exclusions or misclassifications, because the unexposed group was substantially larger than the exposed group, typically comprising more than four million individuals (Table S2). The number of excluded or misclassified individuals was generally small relative to the size of the unexposed group. The subsequent analyses also suggested that estimates of $\mathrm{R}_0$ remained relatively stable across cohorts (Table S11). Consequently, an overestimation of $\mathrm{R}_1$, combined with a relatively stable $\mathrm{R}_0$, could lead to overestimation of aARs and aRRs.

(ii) Cohort II included individuals whose comorbidities were diagnosed between COVID-19 infection and LC diagnosis. Compared with Cohorts I, III, and IV, this broader exposure definition generally increased the denominator of $R_1$ (i.e., the exposed population at risk). Similarly, this inclusion would reduce $R_1$ relative to $R_0$, potentially resulting in smaller aAR and aRR estimates in Cohort II.

Table S1: Numbers of individuals with the target comorbidity who met the removal criteria specified in Table 1 of the main text for Cohort IV.

| **Comorbidity** | **Removed by LC exclusion window** | **Removed by non-LC 180-day exclusion window** | **Removed total** |
|---|---|---|---|
| Any comorbidities | 9,074 | 354,269 | 363,343 |
| Hypertension | 6,510 | 227,587 | 234,097 |
| Chronic lung disease | 7,976 | 150,301 | 158,277 |
| Kidney disease | 2,779 | 74,713 | 77,492 |
| Liver disease | 2,393 | 55,649 | 58,042 |
| ***Neuro psych disorders*** | 5,495 | 153,146 | 158,641 |
| Depression | 4,584 | 116,397 | 120,981 |
| Cerebrovascular disease | 1,548 | 41,309 | 42,857 |
| Dementia | 638 | 27,824 | 28,462 |
| ***Diabetes*** | 3,852 | 118,805 | 122,657 |
| Diabetes without complications | 3,842 | 118,214 | 122,056 |
| Diabetes with complications | 3,306 | 84,009 | 87,315 |
| ***Cardiovascular*** | 5,227 | 115,890 | 121,117 |
| Coronary artery disease | 2,612 | 67,327 | 69,939 |
| Heart failure | 3,479 | 67,244 | 70,723 |
| Peripheral vascular disease | 1,237 | 30,707 | 31,944 |
| Myocardial infarction | 1,943 | 43,320 | 45,263 |
| Cardiomyopathy | 1,398 | 27,417 | 28,815 |
| ***Cancer*** | 3,455 | 117,965 | 121,420 |
| Malignant tumor | 1,340 | 46,013 | 47,353 |
| Benign tumor | 2,826 | 95,485 | 98,311 |

### S2 Data description analysis

Table S2: Absolute numbers and proportions of patients with pre-infection comorbidities (illustrated using Cohort IV) or pre-LC comorbidities (Cohort II).

| **Exposure** | **Without LC (= 0)** | | | | **With LC (=1)** | | | |
|---|---|---|---|---|---|---|---|---|
| | **Exposure = 1** | | **Total** | | **Exposure = 1** | | **Total** | |
| | **Pre-infection** | **Pre-LC** | **Pre-infection** | **Pre-LC** | **Pre-infection** | **Pre-LC** | **Pre-infection** | **Pre-LC** |
| Any comorbidities | 2,592,020 (46.4%) | 2,947,163 (49.6%) | 5,591,562 | 5,947,329 | 59,998 (74.1%) | 63,400 (71.6%) | 81,003 | 88,586 |
| Hypertension | 1,470,630 (25.7%) | 1,699,604 (28.6%) | 5,717,956 | 5,947,329 | 37,309 (44.9%) | 39,687 (44.8%) | 83,037 | 88,586 |
| Chronic lung disease | 717,025 (12.4%) | 869,105 (14.6%) | 5,795,023 | 5,947,329 | 22,454 (27.6%) | 25,432 (28.7%) | 81,492 | 88,586 |
| Kidney disease | 359,928 (6.1%) | 436,286 (7.3%) | 5,870,815 | 5,947,329 | 10,386 (12.0%) | 11,324 (12.8%) | 86,212 | 88,586 |
| Liver disease | 264,395 (4.5%) | 321,138 (5.4%) | 5,890,439 | 5,947,329 | 7,609 (8.8%) | 8,497 (9.6%) | 86,466 | 88,586 |
| ***Neuro psych disorders*** | 911,083 (15.7%) | 1,066,011 (17.9%) | 5,792,006 | 5,947,329 | 26,319 (31.4%) | 28,232 (31.9%) | 83,897 | 88,586 |
| Depression | 753,446 (12.9%) | 870,849 (14.6%) | 5,829,617 | 5,947,329 | 22,565 (26.7%) | 24,217 (27.3%) | 84,659 | 88,586 |
| Cerebrovascular disease | 189,675 (3.2%) | 231,951 (3.9%) | 5,904,929 | 5,947,329 | 5,798 (6.6%) | 6,239 (7.0%) | 87,287 | 88,586 |
| Dementia | 70,716 (1.2%) | 99,693 (1.7%) | 5,918,304 | 5,947,329 | 1,312 (1.5%) | 1,465 (1.7%) | 88,038 | 88,586 |
| ***Diabetes*** | 687,719 (11.8%) | 807,461 (13.6%) | 5,827,421 | 5,947,329 | 17,479 (20.5%) | 18,933 (21.4%) | 85,251 | 88,586 |

Table S2: Absolute numbers and proportions of patients with pre-infection comorbidities (illustrated using Cohort IV) or pre-LC comorbidities (Cohort II).

| **Exposure** | **Without LC (= 0)** | | | | **With LC (=1)** | | | |
|---|---|---|---|---|---|---|---|---|
| | **Exposure = 1** | | **Total** | | **Exposure = 1** | | **Total** | |
| | **Pre-infection** | **Pre-LC** | **Pre-infection** | **Pre-LC** | **Pre-infection** | **Pre-LC** | **Pre-infection** | **Pre-LC** |
| Diabetes with complications | 423,250 (7.2%) | 508,363 (8.5%) | 5,862,059 | 5,947,329 | 11,755 (13.7%) | 13,044 (14.7%) | 85,614 | 88,586 |
| Diabetes without complications | 670,276 (11.5%) | 789,417 (13.3%) | 5,828,015 | 5,947,329 | 17,110 (20.1%) | 18,572 (21.0%) | 85,250 | 88,586 |
| ***Cardiovascular*** | 579,218 (9.9%) | 697,078 (11.7%) | 5,829,241 | 5,947,329 | 16,441 (19.6%) | 18,278 (20.6%) | 84,019 | 88,586 |
| Coronary artery disease | 353,176 (6.0%) | 421,667 (7.1%) | 5,878,695 | 5,947,329 | 10,052 (11.6%) | 10,893 (12.3%) | 86,350 | 88,586 |
| Heart failure | 260,441 (4.4%) | 329,580 (5.5%) | 5,878,058 | 5,947,329 | 7,942 (9.3%) | 9,160 (10.3%) | 85,495 | 88,586 |
| Peripheral vascular disease | 165,386 (2.8%) | 196,754 (3.3%) | 5,915,858 | 5,947,329 | 5,196 (5.9%) | 5,576 (6.3%) | 87,543 | 88,586 |
| Myocardial infarction | 143,506 (2.4%) | 188,179 (3.2%) | 5,902,573 | 5,947,329 | 4,269 (4.9%) | 4,921 (5.6%) | 86,871 | 88,586 |
| Cardiomyopathy | 112,027 (1.9%) | 140,316 (2.4%) | 5,918,968 | 5,947,329 | 3,260 (3.7%) | 3,757 (4.2%) | 87,332 | 88,586 |
| ***Cancer*** | 894,958 (15.4%) | 1,013,579 (17.0%) | 5,828,258 | 5,947,329 | 22,831 (26.6%) | 23,904 (27.0%) | 85,770 | 88,586 |
| Malignant tumor | 353,261 (6.0%) | 400,186 (6.7%) | 5,900,257 | 5,947,329 | 8,752 (10.0%) | 9,186 (10.4%) | 87,515 | 88,586 |
| Benign tumor | 687,307 (11.7%) | 782,842 (13.2%) | 5,851,388 | 5,947,329 | 18,295 (21.2%) | 19,147 (21.6%) | 86,248 | 88,586 |

Table S3: Demographic characteristics of COVID-19 cases by LC status.

| **Characteristics** | **No. (%)** | |
|---|---|---|
| | **Without LC (= 0) N = 6,041,827** | **With LC (=1) N = 88,586** |
| Sex | | |
| Male | 2,593,293 (42.93) | 31,741 (35.83) |
| Female | 3,444,477 (57.02) | 56,827 (64.15) |
| No matching concept | 3,488 (<0.1) | <20 (<0.1) |
| Missing | 569 (<0.1) | <20 (<0.1) |
| Race | | |
| White | 4,173,918 (76.69) | 66,515 (81.09) |
| Black or African American | 782,412 (14.38) | 10,990 (13.40) |
| Asian | 196,114 (3.60) | 2,557 (3.12) |
| American Indian or Alaska Native | 29,132 (0.54) | 620 (0.76) |
| Native Hawaiian or Other Pacific Islander | 22,864 (0.42) | 404 (0.49) |
| Other | 238,266 (4.38) | 941 (1.15) |
| Missing | 599,121 (9.9) | 6,559 (7.4) |
| Age, mean (SD) | 51.91 (18.90) | 58.30 (16.68) |
| BMI, mean (SD) | 32.35 (8.76) | 34.39 (9.43) |
| Poverty Status, mean (SD) | 13.19 (8.36) | 13.11 (8.31) |
| Health Insurance, mean (SD) | 92.38 (4.68) | 92.64 (4.39) |
| COVID vaccine doses | | |
| 0 | 3,667,494 (60.70) | 46,986 (53.04) |
| 1 | 315,831 (5.23) | 6,113 (6.90) |
| 2 | 891,435 (14.75) | 13,563 (15.31) |
| 3 | 647,506 (10.72) | 10,927 (12.33) |
| 4 | 519,561 (8.60) | 10,997 (12.41) |
| **Pre-COVID Comorbidities** | | |
| Any comorbidities | 2,658,175 (44.00) | 62,774 (70.86) |

Table S3: Demographic characteristics of COVID-19 cases by LC status.

| Characteristics | No. (%) | |
|---|---|---|
| Hypertension | 1,522,940 (25.21) | 39,068 (44.10) |
| Chronic lung disease | 744,722 (12.33) | 24,330 (27.46) |
| Kidney disease | 386,847 (6.40) | 11,083 (12.51) |
| Liver disease | 274,094 (4.54) | 8,172 (9.22) |
| ***Neuro psych disorders*** | 941,229 (15.58) | 27,874 (31.47) |
| Depression | 770,327 (12.75) | 23,885 (26.96) |
| Cerebrovascular disease | 202,548 (3.35) | 6,223 (7.02) |
| Dementia | 81,680 (1.35) | 1,457 (1.64) |
| ***Diabetes*** | 716,687 (11.86) | 18,421 (20.79) |
| Diabetes without complications | 698,383 (11.56) | 18,035 (20.36) |
| Diabetes with complications | 445,422 (7.37) | 12,414 (14.01) |
| ***Cardiovascular*** | 616,666 (10.21) | 17,693 (19.97) |
| Coronary artery disease | 376,513 (6.23) | 10,722 (12.10) |
| Heart failure | 284,365 (4.71) | 8,731 (9.86) |
| Peripheral vascular disease | 177,923 (2.94) | 5,501 (6.21) |
| Myocardial infarction | 155,323 (2.57) | 4,665 (5.27) |
| Cardiomyopathy | 120,923 (2.00) | 3,572 (4.03) |
| ***Cancer*** | 919,686 (15.22) | 23,914 (27.00) |
| Malignant tumor | 372,146 (6.16) | 9,200 (10.39) |
| Benign tumor | 698,791 (11.57) | 19,123 (21.59) |
| **Combine Comorbidities** | | |
| Any comorbidities | 3,031,912 (50.18) | 63,400 (71.57) |
| Hypertension | 1,768,371 (29.27) | 39,687 (44.80) |
| Chronic lung disease | 909,588 (15.05) | 25,432 (28.71) |
| Kidney disease | 473,520 (7.84) | 11,324 (12.78) |
| Liver disease | 338,082 (5.60) | 8,497 (9.59) |
| ***Neuro psych disorders*** | 1,106,606 (18.32) | 28,232 (31.87) |
| Depression | 892,559 (14.77) | 24,217 (27.34) |
| Cerebrovascular disease | 248,993 (4.12) | 6,239 (7.04) |
| Dementia | 116,428 (1.93) | 1,465 (1.65) |
| ***Diabetes*** | 846,993 (14.02) | 18,933 (21.37) |
| Diabetes without complications | 828,004 (13.70) | 18,572 (20.96) |
| Diabetes with complications | 540,504 (8.95) | 13,044 (14.72) |
| ***Cardiovascular*** | 750,494 (12.42) | 18,278 (20.63) |
| Coronary artery disease | 452,078 (7.48) | 10,893 (12.30) |
| Heart failure | 364,862 (6.04) | 9,160 (10.34) |
| Peripheral vascular disease | 212,511 (3.52) | 5,576 (6.29) |
| Myocardial infarction | 210,005 (3.48) | 4,921 (5.56) |
| Cardiomyopathy | 153,043 (2.53) | 3,757 (4.24) |
| ***Cancer*** | 1,041,765 (17.24) | 23,904 (26.98) |
| Malignant tumor | 422,158 (6.99) | 9,186 (10.37) |
| Benign tumor | 795,279 (13.16) | 19,147 (21.61) |

Table S4: Descriptive statistics of variables from other sources.

| Covariates | Mean (SD) | | | | |
|---|---|---|---|---|---|
| | 2020<br>N = 2,580 | 2021<br>N = 2,669 | 2022<br>N = 2,663 | 2023<br>N = 2,332 | 2024<br>N = 1,720 |
| **County Health Rankings and Roadmaps** | | | | | |
| Rate poor health * | 17.85 (4.64) | 19.95 (4.93) | 20.44 (4.78) | 15.81 (4.26) | 17.38 (4.42) |
| Avg days physical unhealthy | 4.00 (0.69) | 4.38 (0.76) | 4.34 (0.73) | 3.49 (0.64) | 3.87 (0.66) |
| Smoking * | 17.45 (3.45) | 21.21 (4.10) | 20.30 (4.16) | 19.77 (4.14) | 18.57 (4.25) |
| Obesity * | 32.92 (5.43) | 33.66 (5.81) | 35.61 (4.30) | 35.96 (4.83) | 36.73 (4.94) |
| Food env index * | 7.56 (1.04) | 7.56 (1.02) | 7.53 (1.06) | 7.53 (1.04) | 7.73 (1.03) |
| Physical inactive * | 27.28 (5.74) | 26.73 (5.90) | 30.13 (5.68) | 25.35 (5.10) | 26.13 (5.43) |
| Exercise opportunity * | 64.93 (22.24) | 64.60 (22.19) | 57.09 (22.87) | 64.12 (22.34) | 66.66 (22.00) |
| Over drinking * | 17.56 (3.14) | 19.06 (3.40) | 19.01 (3.27) | 18.96 (3.20) | 16.82 (2.69) |
| Uninsured * | 10.70 (4.59) | 10.91 (4.67) | 11.34 (4.66) | 10.85 (4.43) | 10.24 (4.28) |
| Vaccinated * | 42.93 (9.09) | 44.25 (9.30) | 44.11 (9.31) | 46.67 (9.56) | 42.80 (9.48) |
| Completed high school * | 88.72 (6.78) | 87.15 (5.91) | 87.82 (5.60) | 88.25 (5.42) | 88.62 (5.23) |
| Rate college | 58.40 (11.59) | 58.55 (11.54) | 59.21 (11.51) | 59.50 (11.48) | 60.41 (11.49) |
| Unemployed * | 4.12 (1.42) | 3.99 (1.39) | 6.85 (2.19) | 4.74 (1.69) | 3.63 (1.14) |
| Children in poverty | 20.72 (8.80) | 19.66 (8.35) | 18.35 (8.02) | 19.58 (8.24) | 18.67 (8.28) |
| Income inequality * | 4.50 (0.75) | 4.50 (0.75) | 4.49 (0.76) | 4.53 (0.76) | 4.56 (0.76) |
| Rate social association * | 11.55 (5.36) | 11.45 (5.35) | 11.30 (5.17) | 10.91 (4.79) | 10.59 (4.52) |
| Avg daily PM25 * | 9.26 (1.87) | 7.81 (1.59) | 8.13 (1.64) | 7.73 (1.61) | 7.80 (1.59) |
| Severe housing * | 13.97 (4.07) | 13.65 (3.91) | 13.41 (3.84) | 13.24 (3.80) | 13.17 (3.87) |
| **Rural-Urban Continuum Codes 2023 *, No. (%)** | | | | | |
| 1 | 423 (16.40) | 428 (16.04) | 425 (15.97) | 411 (17.63) | 348 (20.24) |
| 2 | 356 (13.80) | 364 (13.64) | 368 (13.82) | 348 (14.93) | 303 (17.63) |
| 3 | 325 (12.60) | 330 (12.37) | 324 (12.17) | 298 (12.78) | 232 (13.50) |
| 4 | 191 (7.41) | 193 (7.23) | 192 (7.21) | 186 (7.98) | 134 (7.80) |
| 5 | 67 (2.60) | 69 (2.59) | 70 (2.63) | 64 (2.75) | 43 (2.50) |
| 6 | 325 (12.60) | 338 (12.67) | 339 (12.73) | 297 (12.74) | 208 (12.10) |
| 7 | 196 (7.60) | 212 (7.95) | 217 (8.15) | 172 (7.38) | 114 (6.63) |
| 8 | 339 (13.14) | 357 (13.38) | 362 (13.60) | 282 (12.10) | 180 (10.47) |
| 9 | 357 (13.84) | 377 (14.13) | 365 (13.71) | 273 (11.71) | 157 (9.13) |
| **Social Vulnerability Index** | | | | | |
| POV150 | 24.19 (8.26) | 24.22 (8.21) | 24.26 (8.11) | 24.05 (8.15) | 23.73 (8.33) |
| UNEMP * | 5.19 (2.30) | 5.18 (2.27) | 5.22 (2.30) | 5.28 (2.25) | 5.32 (2.20) |
| HBURD | 22.85 (5.09) | 22.81 (5.09) | 22.83 (5.04) | 23.12 (5.04) | 23.43 (5.15) |
| NOHSDP | 12.20 (5.78) | 12.20 (5.66) | 12.18 (5.60) | 12.10 (5.49) | 12.13 (5.49) |
| UNINSUR * | 8.93 (4.43) | 8.99 (4.49) | 8.95 (4.38) | 8.69 (4.14) | 8.52 (3.99) |
| DISABL | 15.89 (4.39) | 15.93 (4.40) | 15.98 (4.42) | 15.89 (4.45) | 15.69 (4.53) |
| SNGPNT * | 5.91 (2.23) | 5.90 (2.23) | 5.90 (2.19) | 5.97 (2.15) | 6.01 (2.09) |
| LIMENG | 1.60 (2.63) | 1.56 (2.44) | 1.55 (2.44) | 1.56 (2.38) | 1.65 (2.37) |
| MINRTY | 23.15 (19.40) | 23.10 (19.31) | 22.97 (19.07) | 23.35 (19.26) | 24.12 (19.17) |
| MUNIT * | 5.16 (6.07) | 5.07 (6.01) | 5.08 (6.00) | 5.39 (6.17) | 5.96 (6.66) |
| MOBILE * | 12.25 (9.56) | 12.36 (9.54) | 12.37 (9.55) | 12.12 (9.56) | 11.95 (9.76) |
| CROWD * | 2.21 (1.81) | 2.19 (1.64) | 2.20 (1.64) | 2.18 (1.63) | 2.17 (1.57) |
| NOVEH * | 6.09 (3.66) | 6.09 (3.62) | 6.07 (3.61) | 6.16 (3.68) | 6.29 (4.03) |
| GROUPQ * | 3.38 (4.17) | 3.38 (4.14) | 3.37 (4.07) | 3.29 (3.84) | 3.21 (3.78) |

***The set of selected variables using the method in Section S3**

For each comorbidity, we identified a representative medication and calculated the proportion of patients who had initiated that medication before a predefined cutoff date. For each comorbidity group ($X_c = 1$) and comparison group ($X_c = 0$), the medication proportion was calculated as the number of patients with medication initiation before the cutoff date divided by the total number of eligible patients in that group.

Table S5: Numbers and proportions of medication users by comorbidity status in the pre-infection cohort (illustrated using Cohort IV) (rows: comorbidity status [0/1]; columns: medications).

| **Comorbidity (Cohort IV)** | $X_c$ | **Hypertension drug** | **Chronic lung disease drug** | **Kidney disease drug** | **Liver disease drug** | **Depression drug** | **Cerebrovascular disease drug** | **Dementia drug** |
|---|---|---|---|---|---|---|---|---|
| Any comorbidities | 0 | 36,585 (1.2%) | 51,843 (1.7%) | 17,757 (0.6%) | 4,123 (0.1%) | 56,418 (1.9%) | 6,817 (0.2%) | 2,963 (0.1%) |
| | 1 | 113,287 (4.3%) | 63,640 (2.4%) | 49,780 (1.9%) | 5,352 (0.2%) | 65,130 (2.5%) | 24,353 (0.9%) | 6,600 (0.2%) |
| Hypertension | 0 | 66,204 (1.5%) | 192,056 (4.5%) | 55,253 (1.3%) | 16,624 (0.4%) | 158,290 (3.7%) | 19,343 (0.5%) | 8,340 (0.2%) |
| | 1 | 127,507 (8.5%) | 61,766 (4.1%) | 60,907 (4.0%) | 7,021 (0.5%) | 39,601 (2.6%) | 29,584 (2.0%) | 6,697 (0.4%) |
| Chronic lung disease | 0 | 317,014 (6.2%) | 186,550 (3.6%) | 149,842 (2.9%) | 30,972 (0.6%) | 197,268 (3.8%) | 74,342 (1.4%) | 22,109 (0.4%) |
| | 1 | 57,198 (7.7%) | 75,901 (10.3%) | 50,838 (6.9%) | 6,709 (0.9%) | 29,352 (4.0%) | 19,440 (2.6%) | 3,537 (0.5%) |
| Kidney disease | 0 | 343,821 (6.2%) | 360,335 (6.4%) | 172,273 (3.1%) | 36,389 (0.7%) | 234,753 (4.2%) | 80,248 (1.4%) | 21,913 (0.4%) |
| | 1 | 50,655 (13.7%) | 35,806 (9.7%) | 47,703 (12.9%) | 6,310 (1.7%) | 15,039 (4.1%) | 18,084 (4.9%) | 4,036 (1.1%) |
| Liver disease | 0 | 386,371 (6.8%) | 390,820 (6.9%) | 234,410 (4.1%) | 38,639 (0.7%) | 242,176 (4.2%) | 109,332 (1.9%) | 29,231 (0.5%) |
| | 1 | 29,963 (11.0%) | 29,497 (10.8%) | 21,770 (8.0%) | 4,524 (1.7%) | 12,584 (4.6%) | 7,006 (2.6%) | 885 (0.3%) |
| **Neuro psych disorders** | 0 | 288,270 (5.8%) | 271,747 (5.5%) | 160,384 (3.2%) | 27,732 (0.6%) | 114,287 (2.3%) | 60,948 (1.2%) | 7,719 (0.2%) |
| | 1 | 76,406 (8.2%) | 71,436 (7.6%) | 52,295 (5.6%) | 8,506 (0.9%) | 59,782 (6.4%) | 25,675 (2.7%) | 8,067 (0.9%) |
| Depression | 0 | 325,540 (6.3%) | 303,038 (5.9%) | 194,511 (3.8%) | 33,715 (0.7%) | 122,156 (2.4%) | 89,481 (1.7%) | 20,137 (0.4%) |
| | 1 | 57,826 (7.5%) | 60,240 (7.8%) | 38,362 (4.9%) | 7,129 (0.9%) | 57,751 (7.4%) | 15,417 (2.0%) | 4,348 (0.6%) |
| Cerebrovascular disease | 0 | 389,065 (6.7%) | 403,808 (7.0%) | 236,636 (4.1%) | 46,943 (0.8%) | 248,300 (4.3%) | 82,626 (1.4%) | 23,751 (0.4%) |
| | 1 | 29,322 (15.0%) | 24,550 (12.6%) | 23,725 (12.1%) | 3,645 (1.9%) | 9,658 (4.9%) | 18,478 (9.5%) | 3,219 (1.6%) |
| Dementia | 0 | 421,929 (7.1%) | 436,710 (7.4%) | 265,568 (4.5%) | 52,891 (0.9%) | 256,439 (4.3%) | 114,149 (1.9%) | 11,760 (0.2%) |
| | 1 | 9,716 (13.5%) | 7,610 (10.6%) | 9,228 (12.8%) | 1,498 (2.1%) | 5,044 (7.0%) | 4,959 (6.9%) | 8,209 (11.4%) |
| **Diabetes** | 0 | 255,419 (4.9%) | 319,160 (6.1%) | 155,388 (3.0%) | 33,163 (0.6%) | 213,087 (4.1%) | 64,042 (1.2%) | 20,874 (0.4%) |
| | 1 | 67,383 (9.6%) | 36,360 (5.2%) | 39,964 (5.7%) | 4,911 (0.7%) | 19,738 (2.8%) | 17,908 (2.5%) | 3,018 (0.4%) |
| Diabetes with complications | 0 | 304,666 (5.5%) | 356,467 (6.5%) | 181,315 (3.3%) | 37,666 (0.7%) | 231,142 (4.2%) | 75,670 (1.4%) | 23,586 (0.4%) |
| | 1 | 61,274 (14.1%) | 32,691 (7.5%) | 38,291 (8.8%) | 4,786 (1.1%) | 15,046 (3.5%) | 17,374 (4.0%) | 2,674 (0.6%) |
| Diabetes without complications | 0 | 258,889 (5.0%) | 321,527 (6.2%) | 158,348 (3.0%) | 33,597 (0.6%) | 214,116 (4.1%) | 65,563 (1.3%) | 21,155 (0.4%) |
| | 1 | 69,128 (10.1%) | 37,171 (5.4%) | 40,931 (6.0%) | 4,977 (0.7%) | 19,934 (2.9%) | 18,404 (2.7%) | 3,083 (0.4%) |
| **Cardiovascular** | 0 | 279,925 (5.3%) | 301,126 (5.7%) | 109,370 (2.1%) | 30,215 (0.6%) | 218,595 (4.1%) | 27,419 (0.5%) | 17,470 (0.3%) |
| | 1 | 72,272 (12.1%) | 51,917 (8.7%) | 59,291 (10.0%) | 6,599 (1.1%) | 20,884 (3.5%) | 28,438 (4.8%) | 5,175 (0.9%) |
| Coronary artery disease | 0 | 338,564 (6.0%) | 359,454 (6.4%) | 179,192 (3.2%) | 40,387 (0.7%) | 237,795 (4.2%) | 42,183 (0.8%) | 22,544 (0.4%) |
| | 1 | 48,998 (13.5%) | 34,890 (9.6%) | 40,812 (11.2%) | 4,528 (1.2%) | 13,147 (3.6%) | 28,027 (7.7%) | 3,168 (0.9%) |
| Heart failure | 0 | 355,668 (6.2%) | 362,432 (6.4%) | 155,630 (2.7%) | 40,095 (0.7%) | 241,573 (4.2%) | 78,347 (1.4%) | 24,547 (0.4%) |
| | 1 | 40,685 (15.2%) | 36,336 (13.5%) | 53,277 (19.9%) | 4,757 (1.8%) | 11,592 (4.3%) | 19,682 (7.3%) | 2,960 (1.1%) |
| Peripheral vascular disease | 0 | 387,761 (6.6%) | 405,710 (7.0%) | 230,749 (4.0%) | 47,618 (0.8%) | 252,195 (4.3%) | 87,167 (1.5%) | 26,794 (0.5%) |
| | 1 | 31,094 (18.2%) | 23,293 (13.7%) | 27,705 (16.2%) | 3,464 (2.0%) | 7,854 (4.6%) | 16,813 (9.9%) | 2,085 (1.2%) |
| Myocardial infarction | 0 | 388,162 (6.6%) | 403,603 (6.9%) | 229,778 (3.9%) | 47,157 (0.8%) | 252,648 (4.3%) | 79,262 (1.4%) | 27,460 (0.5%) |
| | 1 | 25,146 (17.0%) | 21,042 (14.2%) | 23,286 (15.8%) | 2,978 (2.0%) | 6,699 (4.5%) | 16,961 (11.5%) | 1,585 (1.1%) |
| Cardiomyopathy | 0 | 399,587 (6.8%) | 416,948 (7.1%) | 232,229 (3.9%) | 49,933 (0.8%) | 256,496 (4.4%) | 102,189 (1.7%) | 29,275 (0.5%) |
| | 1 | 20,922 (18.1%) | 16,152 (14.0%) | 24,833 (21.5%) | 2,138 (1.9%) | 4,814 (4.2%) | 10,272 (8.9%) | 847 (0.7%) |
| **Cancer** | 0 | 294,572 (5.9%) | 294,478 (5.9%) | 175,272 (3.5%) | 33,050 (0.7%) | 195,775 (3.9%) | 79,481 (1.6%) | 21,361 (0.4%) |
| | 1 | 84,429 (9.2%) | 69,973 (7.6%) | 48,911 (5.3%) | 8,135 (0.9%) | 38,989 (4.2%) | 22,216 (2.4%) | 4,101 (0.4%) |
| Malignant tumor | 0 | 373,587 (6.6%) | 377,834 (6.7%) | 222,511 (4.0%) | 43,327 (0.8%) | 240,988 (4.3%) | 100,099 (1.8%) | 25,576 (0.5%) |
| | 1 | 36,047 (10.0%) | 27,800 (7.7%) | 24,826 (6.9%) | 3,950 (1.1%) | 12,705 (3.5%) | 11,848 (3.3%) | 2,307 (0.6%) |
| Benign tumor | 0 | 330,758 (6.3%) | 335,964 (6.4%) | 210,744 (4.0%) | 41,061 (0.8%) | 209,953 (4.0%) | 92,930 (1.8%) | 24,757 (0.5%) |
| | 1 | 69,509 (9.9%) | 63,028 (8.9%) | 39,924 (5.7%) | 7,191 (1.0%) | 34,011 (4.8%) | 16,949 (2.4%) | 2,998 (0.4%) |

| **Comorbidity (Cohort I)** | $X_c$ | **Diabetes with complications drug** | **Diabetes without complications drug** | **Coronary artery disease drug** | **Heart failure drug** | **Peripheral vascular disease drug** | **Myocardial infarction drug** | **Cardio-myopathy drug** |
|---|---|---|---|---|---|---|---|---|
| Any comorbidities | 0 | 10,223 (0.3%) | 27,985 (0.9%) | 63,189 (2.1%) | 7,523 (0.2%) | 262 (0.0%) | 43,431 (1.4%) | 21,436 (0.7%) |
| | 1 | 22,759 (0.9%) | 79,075 (3.0%) | 151,220 (5.7%) | 30,179 (1.1%) | 1,209 (0.0%) | 59,785 (2.3%) | 49,284 (1.9%) |
| Hypertension | 0 | 54,675 (1.3%) | 86,699 (2.0%) | 161,361 (3.8%) | 16,896 (0.4%) | 875 (0.0%) | 129,481 (3.0%) | 57,067 (1.3%) |
| | 1 | 35,731 (2.4%) | 80,973 (5.4%) | 153,348 (10.2%) | 36,012 (2.4%) | 1,482 (0.1%) | 70,679 (4.7%) | 55,824 (3.7%) |

| Comorbidity (Cohort I) | $X_c$ | Diabetes with complications drug | Diabetes without complications drug | Coronary artery disease drug | Heart failure drug | Peripheral vascular disease drug | Myocardial infarction drug | Cardio-myopathy drug |
|---|---|---|---|---|---|---|---|---|
| Chronic lung disease | 0 | 148,153 (2.9%) | 216,465 (4.2%) | 428,575 (8.3%) | 85,068 (1.7%) | 3,093 (0.1%) | 310,343 (6.0%) | 162,631 (3.2%) |
| | 1 | 29,420 (4.0%) | 36,948 (5.0%) | 80,907 (10.9%) | 25,051 (3.4%) | 1,183 (0.2%) | 56,671 (7.7%) | 35,596 (4.8%) |
| Kidney disease | 0 | 153,864 (2.8%) | 241,762 (4.3%) | 471,635 (8.4%) | 80,582 (1.4%) | 3,637 (0.1%) | 356,443 (6.4%) | 181,502 (3.2%) |
| | 1 | 31,973 (8.6%) | 31,869 (8.6%) | 67,734 (18.3%) | 24,907 (6.7%) | 988 (0.3%) | 47,734 (12.9%) | 32,126 (8.7%) |
| Liver disease | 0 | 198,876 (3.5%) | 253,741 (4.4%) | 544,282 (9.5%) | 121,393 (2.1%) | 5,217 (0.1%) | 419,354 (7.4%) | 223,520 (3.9%) |
| | 1 | 19,447 (7.1%) | 23,832 (8.8%) | 37,261 (13.7%) | 9,998 (3.7%) | 315 (0.1%) | 30,387 (11.2%) | 16,237 (6.0%) |
| **Neuro psych disorders** | 0 | 136,026 (2.8%) | 197,257 (4.0%) | 388,051 (7.9%) | 84,548 (1.7%) | 2,854 (0.1%) | 278,038 (5.6%) | 153,764 (3.1%) |
| | 1 | 40,514 (4.3%) | 50,322 (5.4%) | 106,447 (11.4%) | 27,363 (2.9%) | 1,403 (0.1%) | 76,239 (8.1%) | 43,946 (4.7%) |
| Depression | 0 | 163,088 (3.2%) | 216,910 (4.2%) | 454,618 (8.8%) | 103,364 (2.0%) | 4,175 (0.1%) | 340,481 (6.6%) | 183,903 (3.6%) |
| | 1 | 31,990 (4.1%) | 39,778 (5.1%) | 76,413 (9.8%) | 18,704 (2.4%) | 783 (0.1%) | 55,466 (7.1%) | 31,573 (4.1%) |
| Cerebrovascular disease | 0 | 205,148 (3.5%) | 268,690 (4.6%) | 522,878 (9.0%) | 116,336 (2.0%) | 3,991 (0.1%) | 398,417 (6.9%) | 216,534 (3.7%) |
| | 1 | 19,094 (9.8%) | 16,324 (8.4%) | 49,092 (25.1%) | 14,302 (7.3%) | 984 (0.5%) | 40,311 (20.6%) | 20,801 (10.6%) |
| Dementia | 0 | 232,626 (3.9%) | 286,694 (4.8%) | 584,781 (9.9%) | 134,759 (2.3%) | 5,439 (0.1%) | 458,267 (7.7%) | 242,933 (4.1%) |
| | 1 | 6,425 (8.9%) | 5,308 (7.4%) | 15,335 (21.3%) | 4,192 (5.8%) | 232 (0.3%) | 12,835 (17.8%) | 7,332 (10.2%) |
| **Diabetes** | 0 | 57,213 (1.1%) | 58,819 (1.1%) | 345,213 (6.6%) | 71,882 (1.4%) | 2,817 (0.1%) | 294,853 (5.7%) | 158,783 (3.0%) |
| | 1 | 30,730 (4.4%) | 84,106 (11.9%) | 90,399 (12.8%) | 23,264 (3.3%) | 935 (0.1%) | 45,248 (6.4%) | 30,423 (4.3%) |
| Diabetes with complications | 0 | 89,695 (1.6%) | 130,470 (2.4%) | 417,193 (7.6%) | 85,036 (1.5%) | 3,206 (0.1%) | 343,140 (6.2%) | 182,510 (3.3%) |
| | 1 | 43,809 (10.1%) | 81,145 (18.7%) | 83,924 (19.3%) | 22,258 (5.1%) | 952 (0.2%) | 45,367 (10.4%) | 28,330 (6.5%) |
| Diabetes without complications | 0 | 59,966 (1.1%) | 62,335 (1.2%) | 350,420 (6.7%) | 73,534 (1.4%) | 2,944 (0.1%) | 298,337 (5.7%) | 160,898 (3.1%) |
| | 1 | 33,787 (4.9%) | 86,587 (12.6%) | 92,797 (13.5%) | 23,874 (3.5%) | 957 (0.1%) | 46,887 (6.8%) | 31,181 (4.5%) |
| **Cardiovascular** | 0 | 123,000 (2.3%) | 201,458 (3.8%) | 356,334 (6.7%) | 45,525 (0.9%) | 801 (0.0%) | 250,081 (4.7%) | 120,069 (2.3%) |
| | 1 | 36,476 (6.1%) | 47,628 (8.0%) | 108,137 (18.2%) | 34,398 (5.8%) | 1,348 (0.2%) | 65,919 (11.1%) | 47,020 (7.9%) |
| Coronary artery disease | 0 | 167,038 (3.0%) | 238,684 (4.3%) | 433,890 (7.7%) | 78,709 (1.4%) | 2,708 (0.0%) | 320,643 (5.7%) | 160,975 (2.9%) |
| | 1 | 25,945 (7.1%) | 29,282 (8.1%) | 82,654 (22.8%) | 27,353 (7.5%) | 1,275 (0.4%) | 54,400 (15.0%) | 37,508 (10.3%) |
| Heart failure | 0 | 173,472 (3.0%) | 254,021 (4.5%) | 490,945 (8.6%) | 75,096 (1.3%) | 3,991 (0.1%) | 362,733 (6.4%) | 181,367 (3.2%) |
| | 1 | 26,845 (10.0%) | 22,195 (8.3%) | 58,565 (21.8%) | 27,806 (10.4%) | 963 (0.4%) | 47,908 (17.9%) | 32,081 (12.0%) |
| Peripheral vascular disease | 0 | 196,775 (3.4%) | 256,020 (4.4%) | 531,381 (9.1%) | 113,720 (1.9%) | 1,266 (0.0%) | 413,714 (7.1%) | 218,605 (3.7%) |
| | 1 | 25,084 (14.7%) | 24,637 (14.4%) | 46,930 (27.5%) | 15,829 (9.3%) | 1,322 (0.8%) | 34,623 (20.3%) | 20,345 (11.9%) |
| Myocardial infarction | 0 | 201,467 (3.4%) | 270,856 (4.6%) | 530,449 (9.1%) | 107,908 (1.8%) | 4,456 (0.1%) | 399,288 (6.8%) | 206,831 (3.5%) |
| | 1 | 17,607 (11.9%) | 13,438 (9.1%) | 38,861 (26.3%) | 15,416 (10.4%) | 662 (0.4%) | 33,206 (22.5%) | 20,166 (13.6%) |
| Cardiomyopathy | 0 | 215,378 (3.7%) | 278,821 (4.7%) | 559,130 (9.5%) | 105,187 (1.8%) | 5,159 (0.1%) | 429,959 (7.3%) | 224,753 (3.8%) |
| | 1 | 13,196 (11.4%) | 9,416 (8.2%) | 28,205 (24.5%) | 18,171 (15.8%) | 381 (0.3%) | 24,994 (21.7%) | 15,275 (13.2%) |
| **Cancer** | 0 | 157,527 (3.2%) | 206,109 (4.1%) | 394,407 (7.9%) | 92,164 (1.8%) | 3,697 (0.1%) | 303,746 (6.1%) | 159,077 (3.2%) |
| | 1 | 36,491 (4.0%) | 53,475 (5.8%) | 122,682 (13.4%) | 26,605 (2.9%) | 1,121 (0.1%) | 78,699 (8.6%) | 46,512 (5.1%) |
| Malignant tumor | 0 | 200,325 (3.6%) | 257,878 (4.6%) | 511,833 (9.1%) | 116,137 (2.1%) | 4,662 (0.1%) | 398,281 (7.1%) | 207,240 (3.7%) |
| | 1 | 16,626 (4.6%) | 21,362 (5.9%) | 54,063 (14.9%) | 13,157 (3.6%) | 636 (0.2%) | 34,806 (9.6%) | 22,260 (6.1%) |
| Benign tumor | 0 | 184,046 (3.5%) | 227,933 (4.4%) | 449,625 (8.6%) | 107,383 (2.1%) | 4,418 (0.1%) | 350,136 (6.7%) | 186,420 (3.6%) |
| | 1 | 31,703 (4.5%) | 43,799 (6.2%) | 101,304 (14.4%) | 21,051 (3.0%) | 803 (0.1%) | 68,851 (9.8%) | 38,711 (5.5%) |

Table S6: Numbers and proportions of medication users by comorbidity status in the pre-LC Cohort II (rows: comorbidity status [0/1]; columns: medications).

| Comorbidity (Cohort II) | $X_c$ | Hypertension drug | Chronic lung disease drug | Kidney disease drug | Liver disease drug | Depression drug | Cerebrovascular disease drug | Dementia drug |
|---|---|---|---|---|---|---|---|---|
| Any comorbidities | 0 | 39,398 (1.3%) | 77,643 (2.6%) | 21,520 (0.7%) | 5,433 (0.2%) | 61,544 (2.0%) | 7,542 (0.2%) | 3,245 (0.1%) |
| | 1 | 127,510 (4.2%) | 80,028 (2.7%) | 58,281 (1.9%) | 6,713 (0.2%) | 75,142 (2.5%) | 27,631 (0.9%) | 7,857 (0.3%) |
| Hypertension | 0 | 72,687 (1.7%) | 262,125 (6.1%) | 70,479 (1.6%) | 22,280 (0.5%) | 176,446 (4.1%) | 22,434 (0.5%) | 9,540 (0.2%) |
| | 1 | 146,812 (8.4%) | 79,582 (4.6%) | 74,165 (4.3%) | 9,435 (0.5%) | 47,121 (2.7%) | 35,049 (2.0%) | 8,429 (0.5%) |
| Chronic lung disease | 0 | 351,941 (6.8%) | 282,579 (5.5%) | 186,210 (3.6%) | 42,546 (0.8%) | 221,205 (4.3%) | 87,359 (1.7%) | 26,177 (0.5%) |
| | 1 | 72,436 (8.1%) | 99,446 (11.1%) | 67,145 (7.5%) | 9,661 (1.1%) | 37,162 (4.2%) | 25,425 (2.8%) | 4,891 (0.5%) |
| Kidney disease | 0 | 382,599 (6.8%) | 491,019 (8.8%) | 217,298 (3.9%) | 50,168 (0.9%) | 263,179 (4.7%) | 94,435 (1.7%) | 25,956 (0.5%) |
| | 1 | 63,207 (14.1%) | 48,626 (10.9%) | 61,729 (13.8%) | 8,773 (2.0%) | 19,224 (4.3%) | 23,452 (5.2%) | 5,540 (1.2%) |

Table S6: Numbers and proportions of medication users by comorbidity status in the pre-LC Cohort II (rows: comorbidity status [0/1]; columns: medications).

| **Comorbidity (Cohort II)** | $X_c$ | **Hypertension drug** | **Chronic lung disease drug** | **Kidney disease drug** | **Liver disease drug** | **Depression drug** | **Cerebrovascular disease drug** | **Dementia drug** |
|---|---|---|---|---|---|---|---|---|
| Liver disease | 0 | 429,724 (7.5%) | 533,134 (9.3%) | 290,157 (5.1%) | 53,739 (0.9%) | 272,141 (4.8%) | 128,819 (2.3%) | 34,817 (0.6%) |
| | 1 | 37,478 (11.4%) | 39,324 (11.9%) | 28,894 (8.8%) | 6,090 (1.8%) | 15,910 (4.8%) | 9,150 (2.8%) | 1,245 (0.4%) |
| **Neuro psych disorders** | 0 | 320,310 (6.5%) | 379,893 (7.7%) | 200,467 (4.1%) | 38,774 (0.8%) | 127,354 (2.6%) | 71,501 (1.4%) | 8,789 (0.2%) |
| | 1 | 92,946 (8.5%) | 91,773 (8.4%) | 67,413 (6.2%) | 11,572 (1.1%) | 71,062 (6.5%) | 31,935 (2.9%) | 10,253 (0.9%) |
| Depression | 0 | 363,559 (7.1%) | 426,242 (8.3%) | 244,236 (4.8%) | 47,895 (0.9%) | 137,160 (2.7%) | 106,292 (2.1%) | 24,179 (0.5%) |
| | 1 | 69,756 (7.8%) | 76,466 (8.5%) | 49,151 (5.5%) | 9,628 (1.1%) | 68,916 (7.7%) | 19,457 (2.2%) | 5,785 (0.6%) |
| Cerebrovascular disease | 0 | 431,954 (7.5%) | 550,347 (9.5%) | 293,795 (5.1%) | 64,407 (1.1%) | 278,802 (4.8%) | 96,894 (1.7%) | 28,095 (0.5%) |
| | 1 | 36,842 (15.5%) | 32,819 (13.8%) | 31,129 (13.1%) | 5,165 (2.2%) | 12,190 (5.1%) | 22,993 (9.7%) | 4,326 (1.8%) |
| Dementia | 0 | 468,570 (7.9%) | 592,860 (10.0%) | 328,597 (5.5%) | 72,526 (1.2%) | 287,859 (4.9%) | 134,325 (2.3%) | 13,595 (0.2%) |
| | 1 | 14,299 (14.1%) | 12,481 (12.3%) | 14,480 (14.3%) | 2,565 (2.5%) | 7,101 (7.0%) | 7,665 (7.6%) | 10,834 (10.7%) |
| **Diabetes** | 0 | 283,285 (5.4%) | 434,279 (8.3%) | 194,060 (3.7%) | 45,280 (0.9%) | 238,714 (4.6%) | 75,260 (1.4%) | 24,657 (0.5%) |
| | 1 | 80,234 (9.7%) | 48,448 (5.9%) | 50,397 (6.1%) | 6,684 (0.8%) | 24,136 (2.9%) | 21,791 (2.6%) | 3,926 (0.5%) |
| Diabetes with complications | 0 | 338,513 (6.1%) | 485,407 (8.8%) | 226,473 (4.1%) | 51,550 (0.9%) | 259,419 (4.7%) | 89,252 (1.6%) | 27,956 (0.5%) |
| | 1 | 74,989 (14.4%) | 44,511 (8.5%) | 49,341 (9.5%) | 6,681 (1.3%) | 18,590 (3.6%) | 21,857 (4.2%) | 3,620 (0.7%) |
| Diabetes without complications | 0 | 287,209 (5.5%) | 437,690 (8.4%) | 197,739 (3.8%) | 45,916 (0.9%) | 239,895 (4.6%) | 77,097 (1.5%) | 25,019 (0.5%) |
| | 1 | 82,320 (10.2%) | 49,304 (6.1%) | 51,534 (6.4%) | 6,788 (0.8%) | 24,361 (3.0%) | 22,412 (2.8%) | 3,989 (0.5%) |
| **Cardiovascular** | 0 | 309,810 (5.8%) | 411,876 (7.7%) | 137,735 (2.6%) | 40,587 (0.8%) | 244,483 (4.6%) | 32,199 (0.6%) | 20,487 (0.4%) |
| | 1 | 88,934 (12.4%) | 69,333 (9.7%) | 74,963 (10.5%) | 9,363 (1.3%) | 26,255 (3.7%) | 33,743 (4.7%) | 6,964 (1.0%) |
| Coronary artery disease | 0 | 376,425 (6.7%) | 491,776 (8.8%) | 224,828 (4.0%) | 55,419 (1.0%) | 266,946 (4.8%) | 50,167 (0.9%) | 26,690 (0.5%) |
| | 1 | 60,222 (13.9%) | 46,636 (10.8%) | 52,302 (12.1%) | 6,332 (1.5%) | 16,480 (3.8%) | 33,813 (7.8%) | 4,262 (1.0%) |
| Heart failure | 0 | 394,264 (6.9%) | 496,241 (8.7%) | 195,735 (3.4%) | 54,507 (1.0%) | 270,879 (4.8%) | 91,540 (1.6%) | 29,027 (0.5%) |
| | 1 | 52,752 (15.6%) | 50,580 (14.9%) | 69,449 (20.5%) | 7,057 (2.1%) | 15,322 (4.5%) | 25,824 (7.6%) | 4,196 (1.2%) |
| Peripheral vascular disease | 0 | 431,965 (7.4%) | 555,419 (9.5%) | 288,493 (4.9%) | 65,637 (1.1%) | 283,723 (4.9%) | 103,429 (1.8%) | 31,895 (0.5%) |
| | 1 | 38,202 (18.9%) | 30,270 (15.0%) | 35,275 (17.4%) | 4,887 (2.4%) | 9,814 (4.9%) | 20,771 (10.3%) | 2,738 (1.4%) |
| Myocardial infarction | 0 | 430,685 (7.4%) | 549,755 (9.4%) | 284,694 (4.9%) | 64,467 (1.1%) | 283,940 (4.9%) | 92,805 (1.6%) | 32,562 (0.6%) |
| | 1 | 33,198 (17.2%) | 30,106 (15.6%) | 32,496 (16.8%) | 4,535 (2.3%) | 8,997 (4.7%) | 22,076 (11.4%) | 2,477 (1.3%) |
| Cardiomyopathy | 0 | 444,189 (7.5%) | 569,278 (9.7%) | 289,758 (4.9%) | 68,605 (1.2%) | 288,553 (4.9%) | 119,976 (2.0%) | 34,863 (0.6%) |
| | 1 | 26,850 (18.6%) | 22,821 (15.8%) | 32,973 (22.9%) | 3,336 (2.3%) | 6,535 (4.5%) | 13,748 (9.5%) | 1,275 (0.9%) |
| **Cancer** | 0 | 329,166 (6.6%) | 403,734 (8.1%) | 218,207 (4.4%) | 45,900 (0.9%) | 219,577 (4.4%) | 94,103 (1.9%) | 25,391 (0.5%) |
| | 1 | 98,933 (9.5%) | 86,636 (8.4%) | 59,640 (5.7%) | 10,511 (1.0%) | 46,335 (4.5%) | 26,701 (2.6%) | 5,052 (0.5%) |
| Malignant tumor | 0 | 415,799 (7.4%) | 514,879 (9.2%) | 275,378 (4.9%) | 59,508 (1.1%) | 271,053 (4.8%) | 118,114 (2.1%) | 30,424 (0.5%) |
| | 1 | 42,872 (10.5%) | 35,741 (8.7%) | 31,047 (7.6%) | 5,275 (1.3%) | 15,239 (3.7%) | 14,484 (3.5%) | 2,954 (0.7%) |
| Benign tumor | 0 | 369,934 (7.1%) | 461,013 (8.8%) | 263,295 (5.0%) | 57,313 (1.1%) | 235,982 (4.5%) | 110,090 (2.1%) | 29,520 (0.6%) |
| | 1 | 81,407 (10.2%) | 77,314 (9.6%) | 48,375 (6.0%) | 9,200 (1.1%) | 40,521 (5.1%) | 20,303 (2.5%) | 3,617 (0.5%) |

| **Comorbidity (Cohort II)** | $X_c$ | **Diabetes with complications drug** | **Diabetes without complications drug** | **Coronary artery disease drug** | **Heart failure drug** | **Peripheral vascular disease drug** | **Myocardial infarction drug** | **Cardiomyopathy drug** |
|---|---|---|---|---|---|---|---|---|
| Any comorbidities | 0 | 15,833 (0.5%) | 30,505 (1.0%) | 70,072 (2.3%) | 8,313 (0.3%) | 282 (0.0%) | 57,573 (1.9%) | 25,282 (0.8%) |
| | 1 | 29,190 (1.0%) | 90,137 (3.0%) | 172,899 (5.7%) | 34,117 (1.1%) | 1,345 (0.0%) | 74,181 (2.5%) | 57,280 (1.9%) |
| Hypertension | 0 | 79,598 (1.9%) | 97,906 (2.3%) | 182,654 (4.3%) | 19,328 (0.4%) | 968 (0.0%) | 167,900 (3.9%) | 69,471 (1.6%) |
| | 1 | 46,798 (2.7%) | 94,608 (5.4%) | 180,877 (10.4%) | 42,053 (2.4%) | 1,737 (0.1%) | 90,043 (5.2%) | 67,356 (3.9%) |
| Chronic lung disease | 0 | 210,397 (4.1%) | 241,220 (4.7%) | 488,430 (9.5%) | 99,559 (1.9%) | 3,473 (0.1%) | 395,266 (7.7%) | 197,106 (3.8%) |
| | 1 | 42,018 (4.7%) | 46,807 (5.2%) | 104,237 (11.7%) | 32,440 (3.6%) | 1,515 (0.2%) | 77,744 (8.7%) | 47,517 (5.3%) |
| Kidney disease | 0 | 222,004 (4.0%) | 270,200 (4.8%) | 537,434 (9.6%) | 93,895 (1.7%) | 4,088 (0.1%) | 451,173 (8.1%) | 221,101 (4.0%) |
| | 1 | 43,504 (9.7%) | 40,230 (9.0%) | 86,568 (19.3%) | 31,421 (7.0%) | 1,283 (0.3%) | 64,292 (14.4%) | 42,244 (9.4%) |
| Liver disease | 0 | 280,653 (4.9%) | 282,591 (5.0%) | 622,382 (10.9%) | 141,833 (2.5%) | 5,901 (0.1%) | 530,252 (9.3%) | 272,553 (4.8%) |
| | 1 | 26,411 (8.0%) | 29,368 (8.9%) | 47,388 (14.4%) | 13,055 (4.0%) | 406 (0.1%) | 40,178 (12.2%) | 21,503 (6.5%) |
| **Neuro psych disorders** | 0 | 197,808 (4.0%) | 220,497 (4.5%) | 442,318 (9.0%) | 98,889 (2.0%) | 3,203 (0.1%) | 355,815 (7.2%) | 187,935 (3.8%) |
| | 1 | 54,175 (5.0%) | 61,007 (5.6%) | 131,532 (12.0%) | 34,396 (3.1%) | 1,717 (0.2%) | 99,192 (9.1%) | 56,246 (5.1%) |

| Comorbidity (Cohort II) | $X_c$ | Diabetes with complications drug | Diabetes without complications drug | Coronary artery disease drug | Heart failure drug | Peripheral vascular disease drug | Myocardial infarction drug | Cardiomyopathy drug |
|---|---|---|---|---|---|---|---|---|
| Depression | 0 | 237,582 (4.6%) | 242,835 (4.7%) | 523,019 (10.2%) | 121,988 (2.4%) | 4,725 (0.1%) | 437,343 (8.5%) | 227,101 (4.4%) |
| | 1 | 41,966 (4.7%) | 47,795 (5.3%) | 93,741 (10.5%) | 23,329 (2.6%) | 1,005 (0.1%) | 71,537 (8.0%) | 39,928 (4.5%) |
| Cerebrovascular disease | 0 | 289,438 (5.0%) | 299,455 (5.2%) | 595,929 (10.3%) | 135,982 (2.3%) | 4,498 (0.1%) | 504,045 (8.7%) | 264,314 (4.6%) |
| | 1 | 26,241 (11.0%) | 20,492 (8.6%) | 62,183 (26.1%) | 18,428 (7.7%) | 1,186 (0.5%) | 53,057 (22.3%) | 27,169 (11.4%) |
| Dementia | 0 | 324,305 (5.5%) | 319,312 (5.4%) | 666,793 (11.2%) | 157,623 (2.7%) | 6,130 (0.1%) | 575,935 (9.7%) | 295,637 (5.0%) |
| | 1 | 10,787 (10.7%) | 7,758 (7.7%) | 23,186 (22.9%) | 6,607 (6.5%) | 362 (0.4%) | 20,457 (20.2%) | 11,781 (11.6%) |
| **Diabetes** | 0 | 88,505 (1.7%) | 65,517 (1.3%) | 393,827 (7.6%) | 83,895 (1.6%) | 3,173 (0.1%) | 374,167 (7.2%) | 193,379 (3.7%) |
| | 1 | 40,282 (4.9%) | 97,901 (11.8%) | 109,198 (13.2%) | 28,351 (3.4%) | 1,122 (0.1%) | 59,286 (7.2%) | 38,370 (4.6%) |
| Diabetes with complications | 0 | 137,303 (2.5%) | 146,197 (2.7%) | 476,943 (8.6%) | 99,417 (1.8%) | 3,608 (0.1%) | 435,429 (7.9%) | 222,656 (4.0%) |
| | 1 | 57,628 (11.1%) | 97,813 (18.8%) | 104,262 (20.0%) | 27,880 (5.3%) | 1,166 (0.2%) | 60,187 (11.5%) | 36,698 (7.0%) |
| Diabetes without complications | 0 | 92,588 (1.8%) | 69,446 (1.3%) | 400,018 (7.7%) | 85,851 (1.6%) | 3,324 (0.1%) | 378,755 (7.2%) | 196,079 (3.8%) |
| | 1 | 43,993 (5.4%) | 100,955 (12.5%) | 112,086 (13.9%) | 29,067 (3.6%) | 1,160 (0.1%) | 61,269 (7.6%) | 39,266 (4.9%) |
| **Cardiovascular** | 0 | 176,689 (3.3%) | 225,055 (4.2%) | 403,563 (7.6%) | 53,424 (1.0%) | 909 (0.0%) | 316,828 (6.0%) | 144,635 (2.7%) |
| | 1 | 49,614 (6.9%) | 58,707 (8.2%) | 133,479 (18.7%) | 41,417 (5.8%) | 1,566 (0.2%) | 86,950 (12.2%) | 59,445 (8.3%) |
| Coronary artery disease | 0 | 239,037 (4.3%) | 266,547 (4.8%) | 495,477 (8.8%) | 93,137 (1.7%) | 3,079 (0.1%) | 408,242 (7.3%) | 197,330 (3.5%) |
| | 1 | 35,150 (8.1%) | 36,028 (8.3%) | 102,162 (23.6%) | 33,671 (7.8%) | 1,549 (0.4%) | 71,566 (16.5%) | 47,311 (10.9%) |
| Heart failure | 0 | 246,039 (4.3%) | 282,948 (5.0%) | 558,022 (9.8%) | 88,079 (1.5%) | 4,506 (0.1%) | 458,742 (8.1%) | 218,783 (3.8%) |
| | 1 | 38,247 (11.3%) | 29,196 (8.6%) | 77,311 (22.8%) | 35,042 (10.3%) | 1,306 (0.4%) | 66,068 (19.5%) | 43,564 (12.9%) |
| Peripheral vascular disease | 0 | 281,786 (4.8%) | 286,493 (4.9%) | 609,015 (10.4%) | 133,877 (2.3%) | 1,459 (0.0%) | 524,833 (9.0%) | 268,059 (4.6%) |
| | 1 | 32,846 (16.2%) | 30,079 (14.9%) | 58,227 (28.8%) | 19,817 (9.8%) | 1,542 (0.8%) | 44,850 (22.2%) | 25,908 (12.8%) |
| Myocardial infarction | 0 | 284,475 (4.9%) | 301,677 (5.2%) | 603,983 (10.3%) | 126,225 (2.2%) | 5,023 (0.1%) | 504,138 (8.6%) | 251,649 (4.3%) |
| | 1 | 25,662 (13.3%) | 17,891 (9.3%) | 52,076 (27.0%) | 20,599 (10.7%) | 907 (0.5%) | 46,252 (24.0%) | 27,615 (14.3%) |
| Cardiomyopathy | 0 | 303,438 (5.2%) | 310,779 (5.3%) | 638,683 (10.8%) | 124,182 (2.1%) | 5,820 (0.1%) | 543,534 (9.2%) | 273,486 (4.6%) |
| | 1 | 19,023 (13.2%) | 12,389 (8.6%) | 37,418 (26.0%) | 22,821 (15.8%) | 539 (0.4%) | 34,627 (24.0%) | 21,044 (14.6%) |
| **Cancer** | 0 | 224,272 (4.5%) | 230,590 (4.6%) | 454,746 (9.1%) | 108,849 (2.2%) | 4,173 (0.1%) | 388,887 (7.8%) | 195,998 (3.9%) |
| | 1 | 46,461 (4.5%) | 62,984 (6.1%) | 144,678 (13.9%) | 31,818 (3.1%) | 1,324 (0.1%) | 96,895 (9.3%) | 56,116 (5.4%) |
| Malignant tumor | 0 | 281,341 (5.0%) | 287,718 (5.1%) | 586,113 (10.4%) | 136,501 (2.4%) | 5,252 (0.1%) | 503,901 (9.0%) | 253,287 (4.5%) |
| | 1 | 21,984 (5.4%) | 25,523 (6.2%) | 64,890 (15.9%) | 16,017 (3.9%) | 769 (0.2%) | 43,975 (10.7%) | 27,475 (6.7%) |
| Benign tumor | 0 | 261,892 (5.0%) | 254,888 (4.9%) | 518,887 (9.9%) | 126,896 (2.4%) | 4,992 (0.1%) | 447,896 (8.6%) | 230,423 (4.4%) |
| | 1 | 39,746 (5.0%) | 51,696 (6.4%) | 119,219 (14.9%) | 25,175 (3.1%) | 945 (0.1%) | 83,956 (10.5%) | 46,491 (5.8%) |

Table S7: Four census regions of the U.S.

| Census regions | State | Census regions | State | |
|---|---|---|---|---|
| **Northeast** | | **Midwest** | | |
| | Connecticut<br>Maine<br>Massachusetts<br>New Hampshire<br>Rhode Island<br>Vermont<br>New Jersey<br>New York<br>Pennsylvania | | Illinois<br>Indiana<br>Michigan<br>Ohio<br>Wisconsin<br>Iowa<br>Kansas<br>Minnesota<br>Missouri | Nebraska<br>North Dakota<br>South Dakota |
| **West** | | **South** | | |
| | Arizona<br>Colorado<br>Idaho<br>Montana<br>Nevada | | Delaware<br>District of Columbia<br>Florida<br>Georgia<br>Maryland | Arkansas<br>Louisiana<br>Oklahoma<br>Texas |

Table S7: Four census regions of the U.S.

| Census regions | State | Census regions | State |
|---|---|---|---|
| | New Mexico | | North Carolina |
| | Utah | | South Carolina |
| | Wyoming | | Virginia |
| | Alaska | | West Virginia |
| | California | | Alabama |
| | Hawaii | | Kentucky |
| | Oregon | | Mississippi |
| | Washington | | Tennessee |

### S3 Select minimally redundant variables

For county-level covariables from CHRR, SVI, and RUCC, we use the following procedure to obtain their minimal redundant variables for developing DML models:

(a) **Compute correlations**: Calculate the Pearson correlation matrix **C**, among all variables.

(b) **Identify correlated group**: Pick one target variable and collect all variables whose absolute correlation with it exceeds the threshold (e.g., 0.7).

(c) **Compare redundancy**: For the target variable and its correlated variables, to compute their average absolute correlations with all other variables.

(d) **Select representative**: Add the variable with the smallest average correlation (i.e., less redundant) to the candidate list.

(e) Repeat steps (b)-(d) until all variables are processed.

This procedure resulted in a minimally redundant set of variables (Supplementary **Table** S4). These variables, along with the demographic characteristics in Supplementary **Table** S3 and the 14 medications in Supplementary **Tables** S5–S6, were included as adjustment covariates in the development of our ensemble DML models according to **Algorithm** 1 in the main text (**Table** S8).

Table S8: Adjustment covariables for developing DML.

| No. | Full name | Definition |
|---|---|---|
| 1 | Age | >=18 |
| 2 | Sex | Categorical variable indicating patient sex (Female, Male, Other, Unknown). |
| 3 | Race | Categorical variable indicating patient race/ethnicity (White, Black or African American, Asian, American Indian or Alaska Native, Native Hawaiian or Other Pacific Islander, Hispanic or Latino, Other, Unknown). |
| 4 | Count of doses | Count of COVID-19 vaccine doses received by the patient. |
| 5 | BMI | Body mass index (BMI) |
| 6 | Rate poor health | Percentage of adults reporting fair or poor health (age-adjusted) |
| 7 | Smoking | Percentage of adults who are current smokers (age-adjusted). |
| 8 | Obesity | Percentage of the adult population (age 18 and older) that reports a body mass index (BMI) greater than or equal to 30 kg/m2 (age-adjusted). |
| 9 | Food env. index | Index of factors that contribute to a healthy food environment, from 0 (worst) to 10 (best) |
| 10 | Physical inactive | Percentage of adults that report no leisure-time physical activity |
| 11 | Exercise opportunity | Percentage of the population with access to places for physical activity |
| 12 | Over drinking | Percentage of adults reporting binge or heavy drinking (age-adjusted) |
| 13 | Uninsured | Percentage of population under age 65 without health insurance |
| 14 | Vaccinated | Percentage of fee-for-service (FFS) Medicare enrollees who had an annual flu vaccination |
| 15 | Completed high school | Percentage of adults age 25 and over with a high school diploma or equivalent |
| 16 | Unemployment | Percentage of population ages 16 and older unemployed but seeking work. |
| 17 | Income inequality | Ratio of household income at the 80th percentile to income at the 20th percentile |
| 18 | Rate social association | Number of membership associations per 10,000 population. |
| 19 | PM25 | Average daily density of fine particulate matter in micrograms per cubic meter (PM2.5). |

Table S8: Adjustment covariables for developing DML.

| No. | Full name | Definition |
|---|---|---|
| 20 | Severe housing | Percentage of households with at least 1 of 4 housing problems: overcrowding, high housing costs, lack of kitchen facilities, or lack of plumbing facilities. |
| 21 | RUCC | Rural-Urban Continuum Codes for 2023, a county-level classification scheme developed by the United States Department of Agriculture (USDA) that categorizes counties based on population size, degree of urbanization, and adjacency to metropolitan areas. The codes range from 1 (counties in large metropolitan areas) to 9 (completely rural counties not adjacent to a metro area). |
| 22 | MOBILE | Percentage of mobile homes estimate |
| 23 | UNINSUR | Percentage uninsured in the total civilian noninstitutionalized population estimate |
| 24 | NOVEH | Percentage of households with no vehicle available estimate |
| 25 | CROWD | Percentage of occupied housing units with more people than rooms estimate |
| 26 | UNEMP | Unemployment Rate estimate |
| 27 | SNGPNT | Percentage of single-parent households with children under 18 estimates |
| 28 | MUNIT | Percentage of housing in structures with 10 or more units estimate |
| 29 | GROUPQ | Percentage of persons in group quarters estimate |
| 30 | Health insurance | Reflecting access to healthcare coverage |
| 31 | Poverty status | Socioeconomic disadvantage related to poverty |
| | **Comorbidity** | **Drug** |
| 32 | Hypertension | Lisinopril |
| 33 | Chronic lung disease | Albuterol |
| 34 | Kidney disease | Furosemide |
| 35 | Liver disease | Lactulose |
| 36 | Depression | Sertraline |
| 37 | Cerebrovascular disease | Clopidogrel |
| 38 | Dementia | Donepezil |
| 39 | Diabetes without complications | Metformin |
| 40 | Diabetes with complications | Insulin |
| 41 | Coronary artery disease | Atorvastatin |
| 42 | Heart failure | Carvedilol |
| 43 | Peripheral vascular disease | Cilostazol |
| 44 | Myocardial infarction | Aspirin |
| 45 | Cardiomyopathy | Metoprolol |
| | **Comorbidity** | **Included 15 comorbidity indicators (excluding target comorbidity)** |
| 46 | Hypertension | Diagnosed before the target comorbidity |
| 47 | Chronic lung disease | Diagnosed before the target comorbidity |
| 48 | Kidney disease | Diagnosed before the target comorbidity |
| 49 | Liver disease | Diagnosed before the target comorbidity |
| 50 | Depression | Diagnosed before the target comorbidity |
| 51 | Cerebrovascular disease | Diagnosed before the target comorbidity |
| 52 | Dementia | Diagnosed before the target comorbidity |
| 53 | Diabetes without complications | Diagnosed before the target comorbidity |
| 54 | Diabetes with complications | Diagnosed before the target comorbidity |
| 55 | Coronary artery disease | Diagnosed before the target comorbidity |
| 56 | Heart failure | Diagnosed before the target comorbidity |
| 57 | Peripheral vascular disease | Diagnosed before the target comorbidity |
| 58 | Myocardial infarction | Diagnosed before the target comorbidity |
| 59 | Cardiomyopathy | Diagnosed before the target comorbidity |
| 60 | Malignant tumor | Diagnosed before the target comorbidity |
| 61 | Benign tumor | Diagnosed before the target comorbidity |

### S4 Developing DML models

Based on K-fold cross-validation, we implemented the following estimation procedure for each specific preexisting comorbidity, where $K = 5$. To support robust estimation in the subgroup analyses, we used stratified sampling when assigning cross-validation folds across regions (Table S6), calendar years, and the number of preexisting comorbidities, thereby helping ensure adequate representation of these characteristics within each fold.

#### S4.1 Details of subpopulation analysis

Taking the multimorbidity subgroup as an example, we counted the number of additional preexisting comorbidities relative to each index comorbidity and classified individuals into three groups: 0, 1, or $\geq 2$ additional comorbidities. Specifically, for each index comorbidity, the remaining comorbidities were coded according to whether they had been documented before the index comorbidity date. This preprocessing step was completed before assigning each individual to a cross-validation fold (i.e., before the stratified sampling above).

Using the full analytical cohort, we applied Steps 1–4 of Algorithm 1 of the main text to estimate propensity scores and predict potential outcomes within each cross-validation fold. We then summarized the predicted propensity scores and potential outcomes obtained in Step 5 of Algorithm 1 across the multimorbidity subgroups (0, 1, or $\geq 2$ additional comorbidities). Within each subgroup, we applied the orthogonal score procedure in Step 6 of Algorithm 1 to estimate $\mathrm{aR}_1$ and $\mathrm{aR}_0$, respectively.

In this work, Steps 1–4 of Algorithm 1 only needed to be performed once, whereas the operations in Steps 5–6 were repeated across the other subpopulation analyses, including regional and temporal trend analyses, to improve computational efficiency. In particular, temporal trends in risk estimates were obtained by applying Steps 5–7 to generate year-specific $\mathrm{aR}_1$, $\mathrm{aR}_0$, aARs, and aRRs.

#### S4.2 Additional implementation details of DML

Logistic regression, GBM, and RF were implemented using the *h2o.glm*, *h2o.gbm*, and *h2o.randomForest* functions in the *h2o* R package, respectively. These three models were used as base learners in a stacked ensemble, with a binomial generalized linear model (GLM) used as the metalearner in Steps 1-4 Algorithm 1 of the main text. Within each outer-fold training sample, the internal 3-fold cross-validation procedure was implemented for each base learner. This inner cross-validation was used to generate out-of-fold predictions from the three models for training the stacked-ensemble metalearner. The machine-learning ensemble was embedded within the outer 5-fold DML cross-fitting procedure described in Algorithm 1 of the Main text.

We used prespecified model settings rather than performing an additional data-adaptive hyperparameter search within each DML fold due to the computational burden. For GBM, we used 200 trees, a maximum tree depth of 6, and a learning rate of 0.05, with a Bernoulli distribution for binary classification of the comorbidity exposure or LC outcome. For RF, we used 200 trees and a maximum tree depth of 6 and enabled `binomial_double_trees = TRUE` for our the binary classification problems. Other parameters were left at their default values in the *h2o* implementation.

For reproducibility, the overall random seed was initialized at 123. Within each outer DML fold, different deterministic seed offsets were used for the propensity-score model and the two potential-outcome models. Specifically, the propensity-score ensemble used a seed of $123 + 1000k$, while the potential outcome models for $X_c = 1$ and $X_c = 0$ used $123 + 2000k$ and $123 + 3000k$, respectively, for outer fold $k$. Within each ensemble, the seeds for individual base learners were further incremented according to the learner index, and the metalearner used an additional offset of 999.

For numerical stability, predicted the propensity scores in Step 2 were truncated to the interval $[0.01, 0.99]$ before being used in the doubly robust estimating equations (Imbens, 2024). In addition, when a training subset contained fewer than 30 observations, contained only one response class, or contained two or fewer observations in the smaller response class, model fitting was replaced by the corresponding empirical mean response probability, which was likewise truncated to $[0.01, 0.99]$. These safeguards were primarily included to avoid unstable estimation in sparse training subsets.

#### S4.3 Interpretation of marginal causal risk

Under appropriate identification assumptions, the reported aARs may be interpreted as marginal causal risk contrasts. However, we acknowledge that these interpretations should be made with caution for two key reasons. First, comorbidity status as an exposure may not correspond to a single well-defined and modifiable causal intervention, because a given comorbidity can arise through heterogeneous pathways and may vary in duration, severity, treatment, and clinical presentation. Second, our estimates may remain subject to biases inherent in EHR data, including potential misclassification and inaccuracies in recorded diagnosis dates and disease onset for conditions such as COVID-19 infection, preexisting comorbidities, and LC.

Nevertheless, to clarify the conditions under which our DML approach may help reduce confounding bias and potentially support a causal interpretation of the resulting estimates, we have added the following standard identification assumptions (Igelström et al., 2022; Imbens, 2024):

**(A1) Consistency**: the observed outcome for an individual equals the potential outcome corresponding to the exposure that the individual actually experienced.

**(A2) No interference**: Individual $i$'s Long COVID outcome depends only on their own comorbidity exposure, not on the comorbidity status of other individuals.

**(A3) Positivity**: For all values of the measured covariates $\boldsymbol{X}_{-c}$ and $\boldsymbol{Z}$ with positive probability, $0 < P(X_c = 1 \mid \boldsymbol{X}_{-c}, \boldsymbol{Z}) < 1$.

**(A4) Conditional exchangeability (ignorability)**: Conditional on the measured covariates $\boldsymbol{X}_{-c}$ and $\boldsymbol{Z}$, the comorbidity exposure $X_c$ is independent of the potential outcomes.

Although the causal identification assumptions cannot all be empirically verified using observed EHR data in N3C, we assessed their plausibility where possible. In practice, (A1) may be affected by inaccuracies in disease diagnosis timing and exposure misclassification. To evaluate the potential impact of comorbidity exposure misclassification, we constructed alternative cohorts using different temporal definitions of comorbidity exposure and compared the resulting estimates across Cohorts I-IV. The assumption (A2) is generally plausible in this setting because the comorbidities considered are noninfectious conditions, such that one individual's comorbidity status would not be expected to directly affect another individual's Long COVID outcome. We assessed (A3) by examining propensity score overlap between exposed and unexposed individuals. We also evaluated covariate balance to assess the adequacy of measured confounding control. However, (A4) cannot be empirically verified in the presence of potential unmeasured confounding. Therefore, we additionally used E-values to assess the robustness of our findings to unmeasured confounding.

### S5 Additional results

#### S5.1 Tables

Table S9: Number (percentage = Number × 100/total number of the subpopulation, %) of each medical condition among individuals under 65 and those at 65 and older across the U.S.

| Conditions | 2020 | | 2021 | | 2022 | | 2023 | | 2024 | | Total |
|---|---|---|---|---|---|---|---|---|---|---|---|
| | <65 | >=65 | <65 | >=65 | <65 | >=65 | <65 | >=65 | <65 | >=65 | |
| Any comorbidities | 279,995 (24.59) | 216,744 (55.80) | 445,136 (31.32) | 272,079 (62.37) | 589,900 (41.51) | 443,561 (73.35) | 179,052 (54.09) | 183,163 (80.23) | 59,691 (59.92) | 51,628 (84.48) | 2,720,949 (44.38) |
| Hypertension | 123,358 (10.83) | 161,033 (41.46) | 193,812 (13.64) | 202,565 (46.44) | 251,716 (17.71) | 335,450 (55.47) | 82,090 (24.80) | 142,553 (62.44) | 28,244 (28.35) | 41,187 (67.39) | 1,562,008 (25.48) |
| Chronic lung disease | 65,826 (5.78) | 58,769 (15.13) | 115,553 (8.13) | 78,178 (17.92) | 165,450 (11.64) | 135,214 (22.36) | 53,811 (16.26) | 59,550 (26.09) | 18,786 (18.86) | 17,915 (29.31) | 769,052 (12.54) |
| Kidney disease | 21,344 (1.87) | 47,022 (12.11) | 35,238 (2.48) | 58,632 (13.44) | 49,550 (3.49) | 104,468 (17.28) | 15,803 (4.77) | 46,432 (20.34) | 5,545 (5.57) | 13,896 (22.74) | 397,930 (6.49) |
| Liver disease | 26,236 (2.30) | 17,704 (4.56) | 46,209 (3.25) | 25,424 (5.83) | 66,790 (4.70) | 44,265 (7.32) | 22,830 (6.90) | 18,767 (8.22) | 8,088 (8.12) | 5,953 (9.74) | 282,266 (4.60) |
| **Neuro psych disorders** | 85,387 (7.50) | 69,044 (17.77) | 152,261 (10.71) | 84,289 (19.32) | 227,281 (15.99) | 157,113 (25.98) | 74,502 (22.51) | 71,409 (31.28) | 26,568 (26.67) | 21,249 (34.77) | 969,103 (15.81) |
| Depression | 80,329 (7.05) | 41,982 (10.81) | 143,828 (10.12) | 55,073 (12.63) | 214,708 (15.11) | 102,393 (16.93) | 70,288 (21.23) | 46,642 (20.43) | 25,144 (25.24) | 13,825 (22.62) | 794,212 (12.96) |
| Cerebrovascular disease | 7,254 (0.64) | 24,804 (6.39) | 12,670 (0.89) | 32,531 (7.46) | 20,183 (1.42) | 62,691 (10.37) | 7,254 (2.19) | 29,655 (12.99) | 2,605 (2.61) | 9,124 (14.93) | 208,771 (3.41) |
| Dementia | 831 (0.07) | 19,406 (5.00) | 1,185 (0.08) | 14,301 (3.28) | 1,907 (0.13) | 27,617 (4.57) | 754 (0.23) | 13,043 (5.71) | 260 (0.26) | 3,833 (6.27) | 83,137 (1.36) |
| **Diabetes** | 66,787 (5.87) | 78,608 (20.24) | 96,927 (6.82) | 94,423 (21.65) | 122,585 (8.63) | 143,921 (23.80) | 40,064 (12.10) | 60,025 (26.29) | 13,977 (14.03) | 17,791 (29.11) | 735,108 (11.99) |
| Diabetes without complications | 65,288 (5.73) | 75,698 (19.49) | 94,973 (6.68) | 91,356 (20.94) | 120,259 (8.46) | 139,763 (23.11) | 39,422 (11.91) | 58,449 (25.60) | 13,798 (13.85) | 17,412 (28.49) | 716,418 (11.69) |
| Diabetes with complications | 37,176 (3.26) | 50,394 (12.97) | 53,946 (3.80) | 61,916 (14.19) | 69,788 (4.91) | 98,269 (16.25) | 23,450 (7.08) | 42,132 (18.46) | 8,163 (8.19) | 12,602 (20.62) | 457,836 (7.47) |
| **Cardiovascular** | 28,758 (2.53) | 80,777 (20.80) | 50,005 (3.52) | 103,055 (23.63) | 68,201 (4.80) | 173,876 (28.75) | 23,435 (7.08) | 75,950 (33.27) | 8,144 (8.17) | 22,158 (36.26) | 634,359(10.35) |
| Coronary artery disease | 13,713 (1.20) | 51,446 (13.24) | 23,619 (1.66) | 68,318 (15.66) | 33,478 (2.36) | 116,152 (19.21) | 11,559 (3.49) | 50,212 (21.99) | 4,021 (4.04) | 14,717 (24.08) | 387,235 (6.32) |
| Heart failure | 13,623 (1.20) | 38,209 (9.84) | 23,225 (1.63) | 46,583 (10.68) | 31,746 (2.23) | 78,758 (13.02) | 10,999 (3.32) | 35,457 (15.53) | 3,831 (3.85) | 10,665 (17.45) | 293,096 (4.78) |
| Peripheral vascular disease | 7,241 (0.64) | 23,248 (5.98) | 12,607 (0.89) | 30,403 (6.97) | 18,083 (1.27) | 52,513 (8.68) | 6,483 (1.96) | 23,726 (10.39) | 2,214 (2.22) | 6,906 (11.30) | 183,424 (2.99) |
| Myocardial infarction | 7,026 (0.62) | 18,488 (4.76) | 13,256 (0.93) | 25,080 (5.75) | 18,268 (1.29) | 43,499 (7.19) | 6,512 (1.97) | 19,623 (8.60) | 2,241 (2.25) | 5,995 (9.81) | 159,988 (2.61) |
| Cardiomyopathy | 6,998 (0.61) | 13,586 (3.50) | 12,211 (0.86) | 18,139 (4.16) | 16,553 (1.16) | 31,295 (5.18) | 5,546 (1.68) | 13,832 (6.06) | 1,980 (1.99) | 4,355 (7.13) | 124,495 (2.03) |
| **Cancer** | 74,637 (6.55) | 69,488 (17.89) | 123,771 (8.71) | 101,593 (23.29) | 188,374 (13.25) | 192,983 (31.91) | 61,324 (18.53) | 85,680 (37.53) | 20,645 (20.72) | 25,105 (41.08) | 943,600 (15.39) |
| Malignant tumor | 20,399 (1.79) | 38,712 (9.97) | 33,934 (2.39) | 56,923 (13.05) | 49,784 (3.50) | 102,451 (16.94) | 16,085 (4.86) | 44,726 (19.59) | 5,306 (5.33) | 13,026 (21.31) | 381,346 (6.22) |
| Benign tumor | 60,418 (5.31) | 43,277 (11.14) | 100,874 (7.10) | 65,529 (15.02) | 158,099 (11.12) | 137,671 (22.77) | 52,128 (15.75) | 63,402 (27.77) | 17,711 (17.78) | 18,805 (30.77) | 717,914 (11.71) |

Table S10: Absolute yearly number (percentage = Number*100/total number of the subpopulation, %) of each medical condition among individuals in urban or rural areas across the U.S.

| Conditions | 2020 | | 2021 | | 2022 | | 2023 | | 2024 | |
|---|---|---|---|---|---|---|---|---|---|---|
| | Rural | Urban | Rural | Urban | Rural | Urban | Rural | Urban | Rural | Urban |
| Any comorbidities | 61,535 (29.86) | 432,716 (32.93) | 110,155 (35.26) | 601,662 (39.30) | 144,871 (47.86) | 871,988 (51.52) | 56,215 (63.21) | 298,153 (65.05) | 17,989 (71.74) | 88,247 (68.83) |
| Hypertension | 37,580 (18.24) | 245,234 (18.67) | 65,014 (20.81) | 328,170 (21.43) | 89,377 (29.53) | 486,646 (28.75) | 37,864 (42.58) | 181,234 (39.54) | 12,434 (49.58) | 53,320 (41.59) |
| Chronic lung disease | 15,677 (7.61) | 108,203 (8.24) | 30,102 (9.64) | 161,842 (10.57) | 45,010 (14.87) | 250,517 (14.80) | 19,392 (21.81) | 91,482 (19.96) | 6,431 (25.65) | 28,540 (22.26) |
| Kidney disease | 9,389 (4.56) | 58,431 (4.45) | 15,218 (4.87) | 77,516 (5.06) | 23,074 (7.62) | 126,721 (7.49) | 10,793 (12.14) | 49,265 (10.75) | 3,551 (14.16) | 14,383 (11.22) |
| Liver disease | 5,450 (2.65) | 38,243 (2.91) | 11,778 (3.77) | 59,274 (3.87) | 16,190 (5.35) | 92,789 (5.48) | 6,667 (7.50) | 33,857 (7.39) | 2,333 (9.30) | 10,943 (8.53) |
| **Neuro psych disorders** | 20,039 (9.73) | 133,883 (10.19) | 38,200 (12.23) | 197,261 (12.88) | 56,979 (18.82) | 323,202 (19.10) | 23,612 (26.55) | 119,969 (26.18) | 7,869 (31.38) | 38,367 (29.92) |
| Depression | 15,970 (7.75) | 106,013 (8.07) | 32,411 (10.38) | 165,753 (10.83) | 47,095 (15.56) | 267,441 (15.80) | 18,765 (21.10) | 96,812 (21.12) | 6,213 (24.78) | 31,853 (24.84) |
| Cerebrovascular disease | 4,368 (2.12) | 27,551 (2.10) | 7,357 (2.36) | 37,498 (2.45) | 12,615 (4.17) | 68,855 (4.07) | 6,306 (7.09) | 29,783 (6.50) | 2,194 (8.75) | 8,930 (6.96) |
| Dementia | 2,735 (1.33) | 17,400 (1.32) | 2,480 (0.79) | 12,868 (0.84) | 4,417 (1.46) | 24,142 (1.43) | 2,498 (2.81) | 10,717 (2.34) | 710 (2.83) | 2,968 (2.31) |

Table S10: Absolute yearly number (percentage = Number*100/total number of the subpopulation, %) of each medical condition among individuals in urban or rural areas across the U.S.

| Conditions | 2020 | | 2021 | | 2022 | | 2023 | | 2024 | |
|---|---|---|---|---|---|---|---|---|---|---|
| | Rural | Urban | Rural | Urban | Rural | Urban | Rural | Urban | Rural | Urban |
| **Diabetes** | 19,440 (9.43) | 125,082 (9.52) | 32,735 (10.48) | 156,914 (10.25) | 41,913 (13.85) | 219,024 (12.94) | 17,765 (19.98) | 79,617 (17.37) | 5,831 (23.25) | 24,191 (18.87) |
| Diabetes without complications | 18,739 (9.09) | 121,386 (9.24) | 31,809 (10.18) | 152,856 (9.98) | 40,744 (13.46) | 213,823 (12.63) | 17,339 (19.50) | 77,881 (16.99) | 5,698 (22.72) | 23,798 (18.56) |
| Diabetes with complications | 11,223 (5.45) | 75,703 (5.76) | 19,173 (6.14) | 95,527 (6.24) | 25,872 (8.55) | 138,139 (8.16) | 11,534 (12.97) | 52,014 (11.35) | 3,771 (15.04) | 15,649 (12.20) |
| **Cardiovascular** | 15,837 (7.69) | 93,082 (7.08) | 26,575 (8.51) | 125,280 (8.18) | 39,740 (13.13) | 197,874 (11.69) | 18,244 (20.52) | 78,744 (17.18) | 5,926 (23.63) | 22,757 (17.75) |
| Coronary artery disease | 9,782 (4.75) | 55,036 (4.19) | 16,464 (5.27) | 74,848 (4.89) | 25,288 (8.35) | 121,783 (7.20) | 11,562 (13.00) | 48,798 (10.65) | 3,727 (14.86) | 14,105 (11.00) |
| Heart failure | 7,542 (3.66) | 43,941 (3.34) | 11,933 (3.82) | 57,184 (3.73) | 18,074 (5.97) | 90,035 (5.32) | 8,796 (9.89) | 36,439 (7.95) | 2,902 (11.57) | 10,738 (8.37) |
| Peripheral vascular disease | 4,451 (2.16) | 25,858 (1.97) | 7,664 (2.45) | 35,003 (2.29) | 11,944 (3.95) | 57,335 (3.39) | 5,809 (6.53) | 23,649 (5.16) | 1,886 (7.52) | 6,668 (5.20) |
| Myocardial infarction | 4,363 (2.12) | 20,989 (1.60) | 7,718 (2.47) | 30,300 (1.98) | 11,596 (3.83) | 48,957 (2.89) | 5,430 (6.11) | 20,117 (4.39) | 1,744 (6.95) | 6,055 (4.72) |
| Cardiomyopathy | 2,749 (1.33) | 17,631 (1.34) | 4,780 (1.53) | 25,146 (1.64) | 7,263 (2.40) | 39,096 (2.31) | 3,391 (3.81) | 15,253 (3.33) | 1,163 (4.64) | 4,656 (3.63) |
| **Cancer** | 17,265 (8.38) | 126,163 (9.60) | 30,746 (9.84) | 193,157 (12.62) | 44,043 (14.55) | 331,713 (19.60) | 18,411 (20.70) | 125,684 (27.42) | 6,104 (24.34) | 37,660 (29.37) |
| Malignant tumor | 8,183 (3.97) | 50,534 (3.85) | 13,805 (4.42) | 76,364 (4.99) | 20,364 (6.73) | 129,317 (7.64) | 8,867 (9.97) | 50,632 (11.05) | 2,891 (11.53) | 14,527 (11.33) |
| Benign tumor | 11,332 (5.50) | 91,997 (7.00) | 21,169 (6.78) | 144,309 (9.43) | 31,203 (10.31) | 260,806 (15.41) | 13,051 (14.68) | 100,482 (21.92) | 4,446 (17.73) | 30,702 (23.94) |

Table S11: $R_0$ and $aR_0$ (%) of LC across preexisting comorbidities.

| Comorbidity | Cohort I | | Cohort II | | Cohort III | | Cohort IV | |
|---|---|---|---|---|---|---|---|---|
| | $R_0$ | $aR_0$ | $R_0$ | $aR_0$ | $R_0$ | $aR_0$ | $R_0$ | $aR_0$ |
| Any comorbidities | 0.764 | 1.099 (1.085, 1.113) | 0.833 | 1.401 (1.383, 1.419) | 0.845 | 1.242 (1.227, 1.257) | 0.647 | 1.008 (0.993, 1.023) |
| Hypertension | 1.095 | 1.219 (1.207, 1.232) | 1.139 | 1.338 (1.324, 1.353) | 1.134 | 1.275 (1.262, 1.288) | 1.044 | 1.168 (1.155, 1.181) |
| Chronic lung disease | 1.216 | 1.292 (1.281, 1.302) | 1.23 | 1.342 (1.331, 1.354) | 1.251 | 1.329 (1.318, 1.340) | 1.134 | 1.216 (1.205, 1.226) |
| Kidney disease | 1.371 | 1.409 (1.399, 1.419) | 1.385 | 1.435 (1.424, 1.446) | 1.383 | 1.423 (1.413, 1.433) | 1.352 | 1.391 (1.381, 1.402) |
| Liver disease | 1.399 | 1.434 (1.424, 1.445) | 1.407 | 1.453 (1.442, 1.463) | 1.408 | 1.444 (1.434, 1.454) | 1.381 | 1.420 (1.396, 1.444) |
| ***Neuro psych disorders*** | 1.193 | 1.285 (1.274, 1.295) | 1.223 | 1.346 (1.334, 1.357) | 1.223 | 1.322 (1.311, 1.333) | 1.151 | 1.255 (1.244, 1.266) |
| Depression | 1.233 | 1.310 (1.299, 1.320) | 1.255 | 1.357 (1.345, 1.368) | 1.258 | 1.337 (1.327, 1.348) | 1.198 | 1.279 (1.268, 1.290) |
| Cerebrovascular disease | 1.413 | 1.443 (1.433, 1.453) | 1.423 | 1.460 (1.450, 1.470) | 1.421 | 1.451 (1.441, 1.461) | 1.404 | 1.435 (1.425, 1.445) |
| Dementia | 1.464 | 1.475 (1.462, 1.488) | 1.471 | 1.498 (1.483, 1.512) | 1.467 | 1.476 (1.464, 1.488) | 1.463 | 1.482 (1.467, 1.497) |
| ***Diabetes*** | 1.319 | 1.387 (1.374, 1.399) | 1.339 | 1.436 (1.421, 1.450) | 1.336 | 1.402 (1.389, 1.415) | 1.294 | 1.346 (1.333, 1.359) |
| Diabetes without complications | 1.322 | 1.378 (1.366, 1.391) | 1.342 | 1.438 (1.424, 1.452) | 1.339 | 1.409 (1.396, 1.421) | 1.297 | 1.357 (1.344, 1.370) |
| Diabetes with complications | 1.363 | 1.408 (1.398, 1.419) | 1.372 | 1.431 (1.420, 1.442) | 1.375 | 1.424 (1.413, 1.434) | 1.336 | 1.382 (1.371, 1.392) |
| ***Cardiovascular*** | 1.305 | 1.366 (1.355, 1.377) | 1.323 | 1.417 (1.406, 1.429) | 1.328 | 1.399 (1.389, 1.410) | 1.26 | 1.330 (1.319, 1.341) |
| Coronary artery disease | 1.375 | 1.418 (1.407, 1.429) | 1.389 | 1.451 (1.438, 1.464) | 1.387 | 1.434 (1.423, 1.445) | 1.358 | 1.406 (1.394, 1.418) |
| Heart failure | 1.387 | 1.429 (1.417, 1.440) | 1.396 | 1.459 (1.445, 1.473) | 1.401 | 1.446 (1.434, 1.457) | 1.357 | 1.402 (1.390, 1.414) |
| Peripheral vascular disease | 1.42 | 1.442 (1.428, 1.456) | 1.426 | 1.459 (1.446, 1.472) | 1.425 | 1.451 (1.440, 1.461) | 1.412 | 1.438 (1.428, 1.449) |
| Myocardial infarction | 1.429 | 1.449 (1.439, 1.459) | 1.435 | 1.469 (1.456, 1.481) | 1.435 | 1.459 (1.449, 1.469) | 1.413 | 1.437 (1.427, 1.447) |
| Cardiomyopathy | 1.439 | 1.454 (1.444, 1.464) | 1.443 | 1.463 (1.451, 1.474) | 1.445 | 1.462 (1.452, 1.472) | 1.428 | 1.445 (1.435, 1.455) |
| ***Cancer*** | 1.266 | 1.351 (1.340, 1.362) | 1.296 | 1.403 (1.391, 1.415) | 1.287 | 1.377 (1.366, 1.389) | 1.249 | 1.343 (1.331, 1.354) |
| Malignant tumor | 1.402 | 1.432 (1.422, 1.442) | 1.414 | 1.447 (1.437, 1.458) | 1.41 | 1.436 (1.426, 1.446) | 1.398 | 1.424 (1.414, 1.434) |
| Benign tumor | 1.307 | 1.379 (1.368, 1.390) | 1.33 | 1.422 (1.410, 1.433) | 1.322 | 1.398 (1.387, 1.409) | 1.293 | 1.376 (1.365, 1.387) |

Table S12: $R_1$ and $aR_1$ (%) of LC across preexisting comorbidities.

| Comorbidity | Cohort I | | Cohort II | | Cohort III | | Cohort IV | |
|---|---|---|---|---|---|---|---|---|
| | $R_1$ | $aR_1$ | $R_1$ | $aR_1$ | $R_1$ | $aR_1$ | $R_1$ | $aR_1$ |
| Any comorbidities | 2.377 | 2.024 (2.006, 2.041) | 2.116 | 1.791 (1.775, 1.806) | 2.274 | 1.922 (1.905, 1.939) | 2.274 | 1.912 (1.894, 1.929) |
| Hypertension | 2.606 | 2.295 (2.266, 2.325) | 2.298 | 1.985 (1.960, 2.011) | 2.492 | 2.147 (2.119, 2.175) | 2.492 | 2.146 (2.117, 2.175) |
| Chronic lung disease | 3.309 | 2.785 (2.745, 2.825) | 2.869 | 2.434 (2.398, 2.469) | 3.062 | 2.551 (2.513, 2.589) | 3.062 | 2.534 (2.496, 2.573) |
| Kidney disease | 3.033 | 2.605 (2.545, 2.665) | 2.572 | 2.254 (2.202, 2.306) | 2.848 | 2.446 (2.388, 2.504) | 2.848 | 2.436 (2.378, 2.494) |

Table S12: $R_1$ and $aR_1$ (%) of LC across preexisting comorbidities.

| **Comorbidity** | **Cohort I** | | **Cohort II** | | **Cohort III** | | **Cohort IV** | |
|---|---|---|---|---|---|---|---|---|
| | $\mathbf{R_1}$ | $\mathbf{aR_1}$ | $\mathbf{R_1}$ | $\mathbf{aR_1}$ | $\mathbf{R_1}$ | $\mathbf{aR_1}$ | $\mathbf{R_1}$ | $\mathbf{aR_1}$ |
| Liver disease | 3.025 | 2.592 (2.534, 2.651) | 2.606 | 2.241 (2.192, 2.291) | 2.822 | 2.394 (2.338, 2.450) | 2.822 | 2.384 (2.330, 2.438) |
| ***Neuro psych disorders*** | 2.99 | 2.559 (2.525, 2.592) | 2.602 | 2.220 (2.191, 2.250) | 2.828 | 2.385 (2.352, 2.417) | 2.828 | 2.379 (2.347, 2.412) |
| Depression | 3.089 | 2.679 (2.642, 2.717) | 2.722 | 2.346 (2.312, 2.379) | 2.924 | 2.510 (2.474, 2.546) | 2.924 | 2.500 (2.464, 2.536) |
| Cerebrovascular disease | 3.225 | 2.898 (2.813, 2.982) | 2.664 | 2.424 (2.351, 2.497) | 3.011 | 2.628 (2.548, 2.709) | 3.011 | 2.632 (2.551, 2.713) |
| Dementia | 2.085 | 2.203 (2.121, 2.286) | 1.5 | 1.674 (1.599, 1.749) | 1.881 | 1.923 (1.846, 2.000) | 1.881 | 1.904 (1.827, 1.981) |
| ***Diabetes*** | 2.63 | 2.182 (2.144, 2.219) | 2.31 | 1.956 (1.921, 1.991) | 2.499 | 2.088 (2.051, 2.125) | 2.499 | 2.088 (2.050, 2.125) |
| Diabetes without complications | 2.642 | 2.235 (2.197, 2.274) | 2.318 | 1.948 (1.913, 1.983) | 2.509 | 2.082 (2.045, 2.119) | 2.509 | 2.078 (2.040, 2.115) |
| Diabetes with complications | 2.88 | 2.492 (2.439, 2.545) | 2.53 | 2.195 (2.148, 2.241) | 2.731 | 2.332 (2.280, 2.383) | 2.731 | 2.322 (2.270, 2.373) |
| ***Cardiovascular*** | 3.004 | 2.721 (2.670, 2.771) | 2.592 | 2.383 (2.339, 2.428) | 2.797 | 2.476 (2.427, 2.524) | 2.797 | 2.463 (2.414, 2.511) |
| Coronary artery disease | 2.988 | 2.788 (2.727, 2.849) | 2.556 | 2.457 (2.403, 2.511) | 2.806 | 2.600 (2.542, 2.658) | 2.806 | 2.595 (2.538, 2.652) |
| Heart failure | 3.309 | 2.854 (2.785, 2.922) | 2.764 | 2.466 (2.407, 2.526) | 3.019 | 2.534 (2.470, 2.598) | 3.019 | 2.527 (2.463, 2.591) |
| Peripheral vascular disease | 3.272 | 2.903 (2.821, 2.985) | 2.805 | 2.506 (2.432, 2.580) | 3.096 | 2.676 (2.596, 2.755) | 3.096 | 2.678 (2.599, 2.757) |
| Myocardial infarction | 3.205 | 2.724 (2.643, 2.804) | 2.603 | 2.272 (2.203, 2.341) | 2.941 | 2.424 (2.343, 2.506) | 2.941 | 2.420 (2.338, 2.501) |
| Cardiomyopathy | 3.147 | 2.861 (2.765, 2.957) | 2.664 | 2.464 (2.380, 2.548) | 2.88 | 2.544 (2.447, 2.641) | 2.88 | 2.538 (2.440, 2.635) |
| ***Cancer*** | 2.621 | 2.344 (2.311, 2.377) | 2.321 | 2.119 (2.090, 2.149) | 2.505 | 2.261 (2.230, 2.293) | 2.505 | 2.251 (2.220, 2.283) |
| Malignant tumor | 2.574 | 2.346 (2.291, 2.402) | 2.278 | 2.146 (2.095, 2.197) | 2.452 | 2.256 (2.202, 2.311) | 2.452 | 2.251 (2.197, 2.305) |
| Benign tumor | 2.718 | 2.457 (2.420, 2.494) | 2.398 | 2.198 (2.165, 2.231) | 2.604 | 2.347 (2.311, 2.383) | 2.604 | 2.337 (2.301, 2.373) |

Table S13: Sensitivity analyses based on different data settings used for DML development.

| Comorbidity | aAR(%) | | | |
|---|---|---|---|---|
| | Non-imputed data excluding decedents | | Imputed data including decedents | |
| | Cohort I | Cohort II | Cohort I | Cohort II |
| Any comorbidities | 0.676 (0.645, 0.707) | -0.114 (-0.150, -0.078) | 0.890 (0.868, 0.913) | 0.341 (0.318, 0.365) |
| Hypertension | 0.835 (0.796, 0.875) | 0.286 (0.245, 0.326) | 1.022 (0.991, 1.053) | 0.593 (0.564, 0.622) |
| Chronic lung disease | 1.329 (1.281, 1.378) | 0.857 (0.812, 0.902) | 1.397 (1.357, 1.437) | 0.991 (0.956, 1.027) |
| Kidney disease | 0.996 (0.926, 1.067) | 0.458 (0.396, 0.519) | 1.057 (0.999, 1.115) | 0.691 (0.641, 0.742) |
| Liver disease | 0.889 (0.819, 0.959) | 0.497 (0.435, 0.560) | 1.076 (1.019, 1.133) | 0.679 (0.631, 0.727) |
| ***Neuro psych disorders*** | 1.069 (1.028, 1.111) | 0.614 (0.575, 0.654) | 1.177 (1.143, 1.211) | 0.791 (0.760, 0.822) |
| Depression | 1.158 (1.113, 1.204) | 0.728 (0.685, 0.771) | 1.294 (1.257, 1.332) | 0.911 (0.877, 0.945) |
| Cerebrovascular disease | 1.154 (1.056, 1.253) | 0.711 (0.625, 0.796) | 1.283 (1.201, 1.364) | 0.866 (0.796, 0.937) |
| Dementia | 0.535 (0.419, 0.652) | 0.296 (0.190, 0.402) | 0.517 (0.439, 0.594) | 0.094 (0.022, 0.165) |
| ***Diabetes*** | 0.670 (0.620, 0.721) | 0.157 (0.106, 0.208) | 0.809 (0.770, 0.848) | 0.451 (0.414, 0.488) |
| Diabetes without complications | 0.650 (0.600, 0.700) | 0.137 (0.086, 0.187) | 0.789 (0.750, 0.828) | 0.445 (0.408, 0.481) |
| Diabetes with complications | 0.851 (0.786, 0.917) | 0.484 (0.425, 0.543) | 0.973 (0.922, 1.025) | 0.643 (0.598, 0.688) |
| ***Cardiovascular*** | 1.185 (1.123, 1.246) | 0.725 (0.670, 0.781) | 1.216 (1.166, 1.267) | 0.838 (0.794, 0.882) |
| Coronary artery disease | 1.231 (1.156, 1.306) | 0.723 (0.656, 0.789) | 1.279 (1.220, 1.338) | 0.917 (0.863, 0.970) |
| Heart failure | 1.263 (1.181, 1.346) | 0.798 (0.725, 0.871) | 1.242 (1.177, 1.307) | 0.835 (0.778, 0.892) |
| Peripheral vascular disease | 1.205 (1.107, 1.303) | 0.810 (0.719, 0.902) | 1.261 (1.183, 1.338) | 0.907 (0.835, 0.978) |
| Myocardial infarction | 0.979 (0.879, 1.079) | 0.495 (0.410, 0.579) | 1.100 (1.019, 1.180) | 0.650 (0.586, 0.715) |
| Cardiomyopathy | 1.131 (1.013, 1.250) | 0.723 (0.623, 0.822) | 1.233 (1.137, 1.329) | 0.853 (0.775, 0.931) |
| ***Cancer*** | 0.764 (0.723, 0.806) | 0.351 (0.312, 0.390) | 0.983 (0.949, 1.017) | 0.676 (0.645, 0.707) |
| Malignant tumor | 0.681 (0.613, 0.749) | 0.377 (0.314, 0.439) | 0.857 (0.803, 0.912) | 0.600 (0.550, 0.650) |
| Benign tumor | 0.795 (0.750, 0.840) | 0.418 (0.376, 0.460) | 1.053 (1.016, 1.091) | 0.754 (0.720, 0.788) |

| Comorbidity | aRR | | | |
|---|---|---|---|---|
| | Non-imputed data excluding decedents | | Imputed data including decedents | |
| | Cohort I | Cohort II | Cohort I | Cohort II |
| Any comorbidities | 1.423 (1.399, 1.448) | 0.947 (0.931, 0.963) | 1.818 (1.790, 1.847) | 1.243 (1.224, 1.262) |
| Hypertension | 1.498 (1.471, 1.525) | 1.149 (1.127, 1.171) | 1.850 (1.819, 1.880) | 1.446 (1.421, 1.471) |
| Chronic lung disease | 1.770 (1.739, 1.801) | 1.468 (1.441, 1.495) | 2.097 (2.062, 2.132) | 1.745 (1.716, 1.775) |
| Kidney disease | 1.526 (1.488, 1.565) | 1.226 (1.196, 1.258) | 1.761 (1.718, 1.804) | 1.487 (1.452, 1.524) |
| Liver disease | 1.462 (1.425, 1.500) | 1.254 (1.222, 1.287) | 1.762 (1.721, 1.804) | 1.473 (1.439, 1.508) |
| ***Neuro psych disorders*** | 1.619 (1.593, 1.646) | 1.334 (1.311, 1.357) | 1.925 (1.895, 1.956) | 1.594 (1.568, 1.620) |
| Depression | 1.660 (1.632, 1.689) | 1.395 (1.371, 1.421) | 2.003 (1.971, 2.035) | 1.680 (1.653, 1.708) |
| Cerebrovascular disease | 1.597 (1.546, 1.649) | 1.362 (1.319, 1.407) | 1.903 (1.846, 1.962) | 1.602 (1.553, 1.653) |
| Dementia | 1.273 (1.214, 1.334) | 1.148 (1.096, 1.203) | 1.356 (1.304, 1.411) | 1.064 (1.016, 1.113) |
| ***Diabetes*** | 1.362 (1.334, 1.391) | 1.077 (1.052, 1.102) | 1.599 (1.569, 1.630) | 1.318 (1.291, 1.345) |
| Diabetes without complications | 1.350 (1.322, 1.379) | 1.067 (1.042, 1.092) | 1.581 (1.550, 1.612) | 1.313 (1.286, 1.340) |
| Diabetes with complications | 1.451 (1.415, 1.487) | 1.249 (1.219, 1.281) | 1.701 (1.663, 1.740) | 1.454 (1.422, 1.488) |
| ***Cardiovascular*** | 1.644 (1.609, 1.679) | 1.373 (1.344, 1.403) | 1.899 (1.861, 1.939) | 1.595 (1.563, 1.629) |
| Coronary artery disease | 1.648 (1.607, 1.689) | 1.359 (1.325, 1.394) | 1.915 (1.872, 1.960) | 1.641 (1.603, 1.681) |
| Heart failure | 1.662 (1.618, 1.707) | 1.407 (1.369, 1.446) | 1.880 (1.833, 1.928) | 1.578 (1.538, 1.620) |
| Peripheral vascular disease | 1.624 (1.573, 1.676) | 1.414 (1.368, 1.463) | 1.886 (1.831, 1.943) | 1.631 (1.580, 1.683) |
| Myocardial infarction | 1.503 (1.452, 1.556) | 1.250 (1.208, 1.294) | 1.769 (1.713, 1.828) | 1.448 (1.404, 1.495) |
| Cardiomyopathy | 1.579 (1.519, 1.642) | 1.367 (1.317, 1.419) | 1.861 (1.794, 1.930) | 1.592 (1.538, 1.649) |
| ***Cancer*** | 1.418 (1.394, 1.443) | 1.178 (1.158, 1.199) | 1.738 (1.710, 1.766) | 1.488 (1.464, 1.513) |
| Malignant tumor | 1.354 (1.319, 1.391) | 1.192 (1.160, 1.224) | 1.610 (1.570, 1.650) | 1.421 (1.385, 1.457) |
| Benign tumor | 1.428 (1.402, 1.454) | 1.213 (1.191, 1.235) | 1.775 (1.745, 1.805) | 1.539 (1.513, 1.565) |

Table S14: Adjusted attributable risk and relative risk (95% CIs) and sensitivity analysis using E–value for pre-infection comorbidities in Cohorts III and IV.

| Preexisting comorbidities | aARs (%; 95% CIs) | | aRR (95% CIs) | | E value | |
|---|---|---|---|---|---|---|
| | Cohort III | Cohort IV | Cohort III | Cohort IV | Cohort III | Cohort IV |
| Any comorbidities | 0.680 (0.657, 0.703) | 0.904 (0.881, 0.926) | 1.547 (1.524, 1.571) | 1.896 (1.864, 1.929) | 2.468 | 3.200 |
| Hypertension | 0.872 (0.841, 0.903) | 0.978 (0.946, 1.009) | 1.684 (1.656, 1.712) | 1.837 (1.806, 1.869) | 2.756 | 3.077 |
| Chronic lung disease | 1.222 (1.183, 1.262) | 1.319 (1.279, 1.358) | 1.920 (1.887, 1.953) | 2.085 (2.049, 2.121) | 3.249 | 3.589 |
| Kidney disease | 1.023 (0.964, 1.082) | 1.045 (0.986, 1.104) | 1.719 (1.677, 1.762) | 1.751 (1.708, 1.795) | 2.830 | 2.897 |
| Liver disease | 0.950 (0.893, 1.007) | 0.964 (0.905, 1.023) | 1.658 (1.618, 1.699) | 1.679 (1.632, 1.727) | 2.702 | 2.746 |
| ***Neuro psych disorders*** | 1.063 (1.029, 1.097) | 1.125 (1.091, 1.159) | 1.804 (1.776, 1.833) | 1.896 (1.866, 1.927) | 3.009 | 3.200 |
| Depression | 1.173 (1.135, 1.210) | 1.221 (1.183, 1.259) | 1.877 (1.846, 1.908) | 1.955 (1.922, 1.987) | 3.160 | 3.321 |
| Cerebrovascular disease | 1.178 (1.096, 1.259) | 1.197 (1.116, 1.279) | 1.812 (1.755, 1.870) | 1.835 (1.778, 1.893) | 3.024 | 3.072 |
| Dementia | 0.447 (0.369, 0.525) | 0.421 (0.343, 0.500) | 1.303 (1.251, 1.357) | 1.284 (1.232, 1.339) | 1.930 | 1.889 |
| ***Diabetes*** | 0.686 (0.647, 0.725) | 0.742 (0.702, 0.781) | 1.489 (1.460, 1.519) | 1.551 (1.520, 1.583) | 2.342 | 2.476 |
| Diabetes without complications | 0.673 (0.634, 0.712) | 0.721 (0.681, 0.760) | 1.478 (1.449, 1.508) | 1.531 (1.500, 1.563) | 2.318 | 2.433 |
| Diabetes with complications | 0.908 (0.855, 0.961) | 0.940 (0.887, 0.993) | 1.638 (1.600, 1.676) | 1.680 (1.641, 1.720) | 2.660 | 2.749 |
| ***Cardiovascular*** | 1.076 (1.027, 1.126) | 1.133 (1.083, 1.182) | 1.769 (1.732, 1.807) | 1.852 (1.813, 1.891) | 2.935 | 3.107 |
| Coronary artery disease | 1.166 (1.107, 1.225) | 1.189 (1.130, 1.247) | 1.813 (1.771, 1.857) | 1.846 (1.802, 1.890) | 3.028 | 3.095 |
| Heart failure | 1.089 (1.023, 1.154) | 1.125 (1.060, 1.190) | 1.753 (1.707, 1.800) | 1.802 (1.755, 1.851) | 2.902 | 3.005 |
| Peripheral vascular disease | 1.225 (1.145, 1.305) | 1.240 (1.160, 1.319) | 1.845 (1.789, 1.902) | 1.862 (1.806, 1.919) | 3.093 | 3.129 |
| Myocardial infarction | 0.965 (0.883, 1.048) | 0.983 (0.901, 1.065) | 1.662 (1.605, 1.720) | 1.684 (1.627, 1.743) | 2.710 | 2.758 |
| Cardiomyopathy | 1.082 (0.984, 1.179) | 1.093 (0.995, 1.191) | 1.740 (1.674, 1.808) | 1.757 (1.689, 1.826) | 2.874 | 2.909 |
| ***Cancer*** | 0.884 (0.850, 0.918) | 0.908 (0.875, 0.942) | 1.642 (1.615, 1.669) | 1.677 (1.649, 1.704) | 2.668 | 2.742 |
| Malignant tumor | 0.821 (0.765, 0.876) | 0.827 (0.772, 0.882) | 1.572 (1.533, 1.611) | 1.580 (1.541, 1.621) | 2.519 | 2.538 |
| Benign tumor | 0.949 (0.912, 0.987) | 0.961 (0.924, 0.999) | 1.679 (1.650, 1.708) | 1.699 (1.670, 1.729) | 2.747 | 2.788 |

#### S5.2 Figures

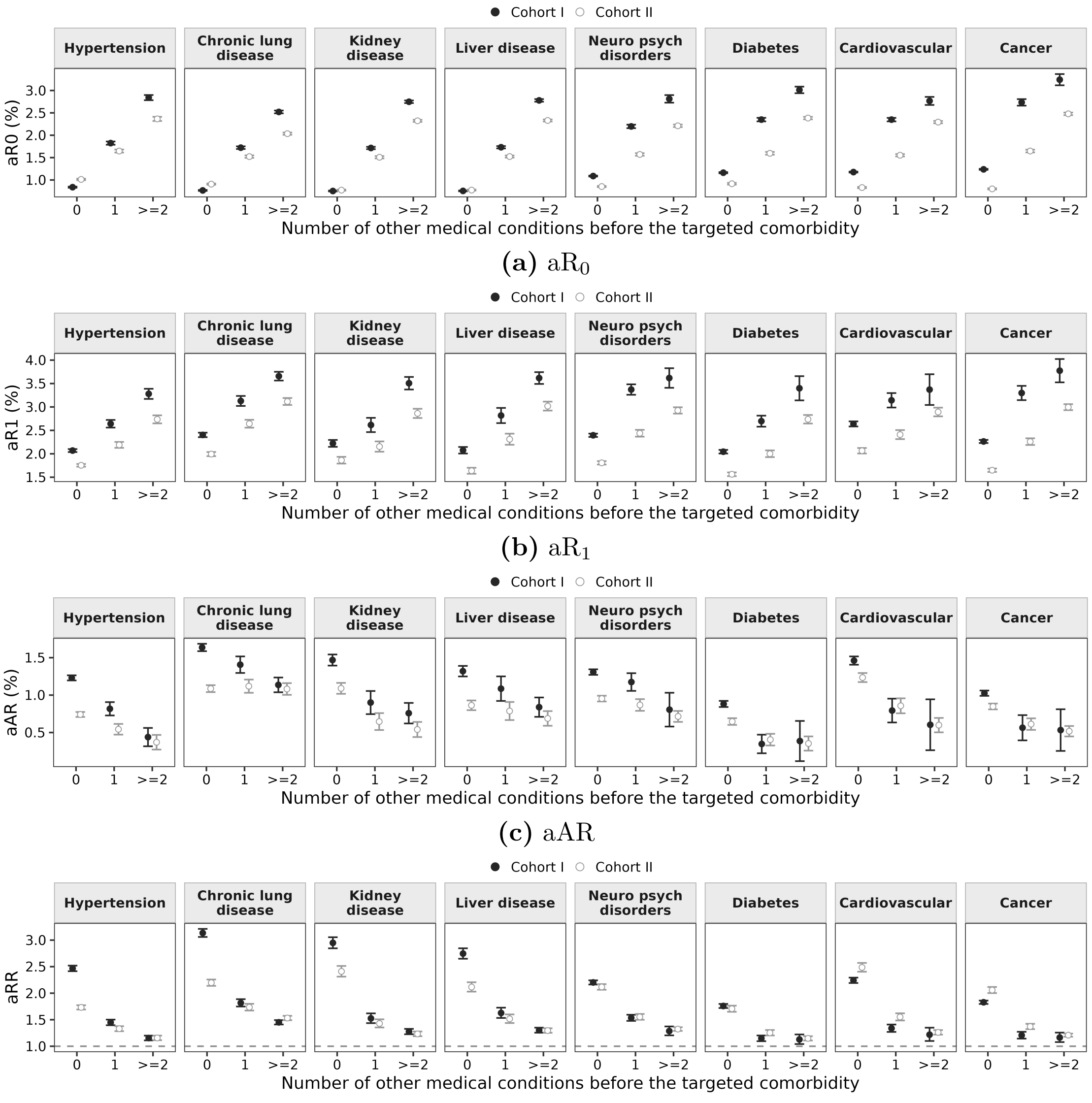


**(a)** $aR_0$

**(b)** $aR_1$

**(c)** aAR

**(d)** aRR

Figure S2: Risks of the target comorbidity by numbers of additional comorbidities.

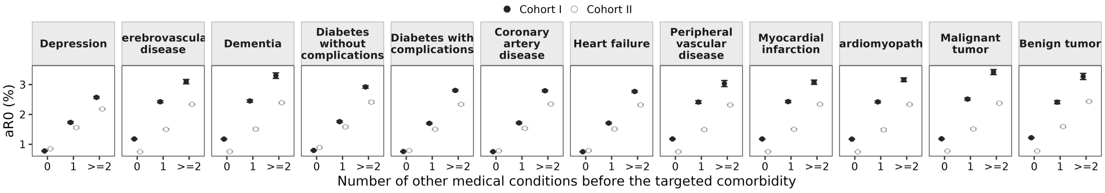


**(a)** $aR_0$

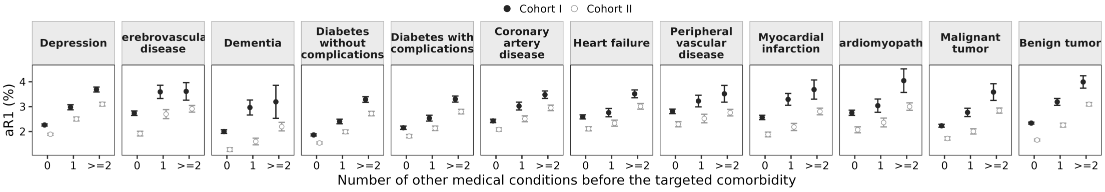


**(b)** $aR_1$

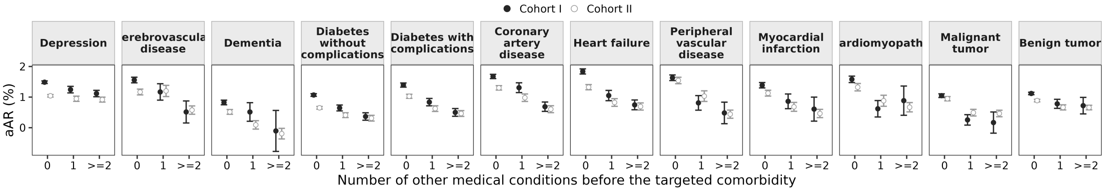


**(c)** aAR

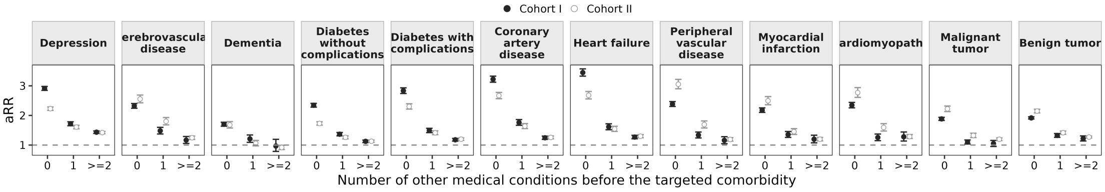


**(d)** aRR

Figure S3: Risks of the target comorbidity by numbers of additional comorbidities.

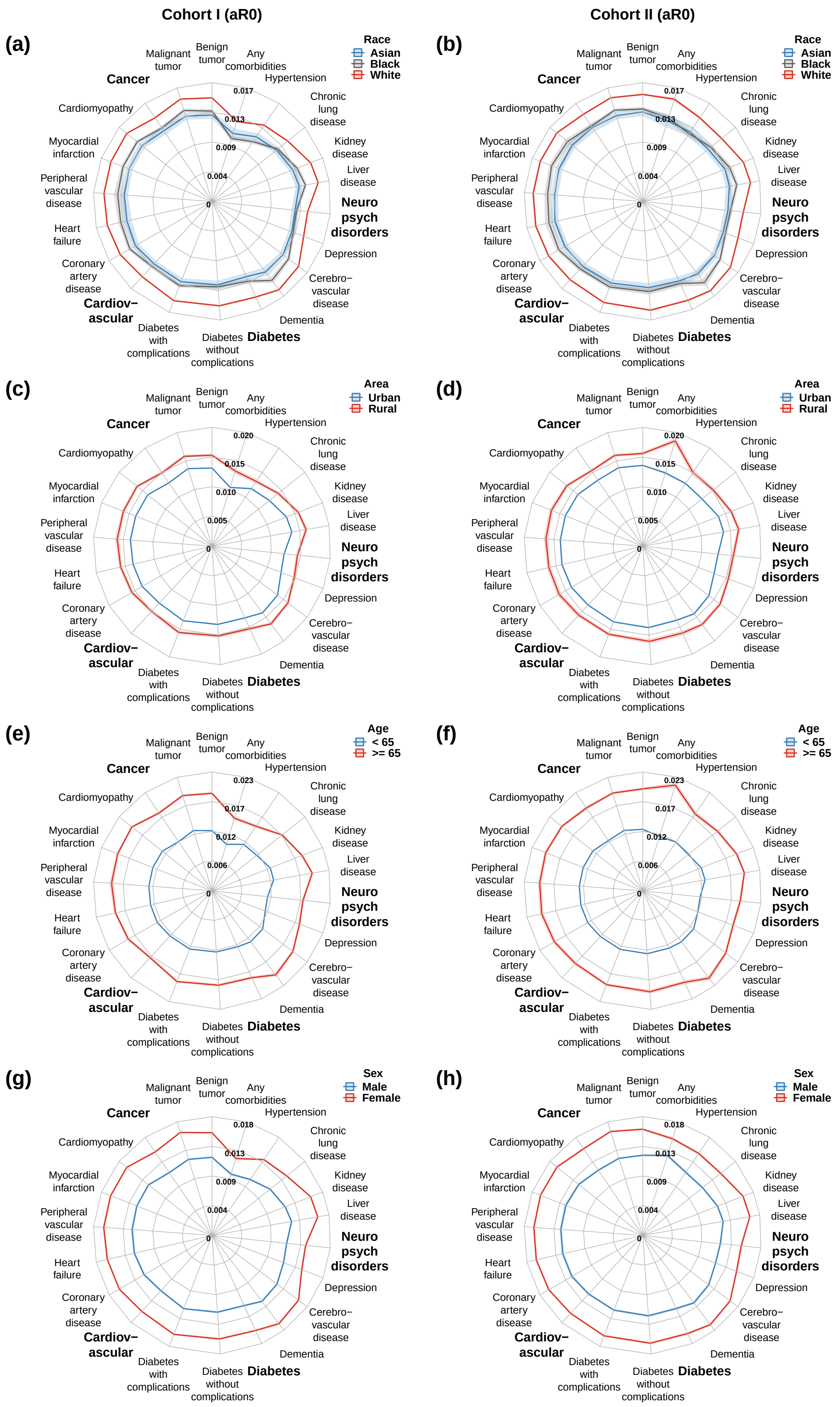


Figure S4: Estimated $aR_0$ of Long COVID associated with preexisting comorbidities across subpopulations, with 95% CIs.

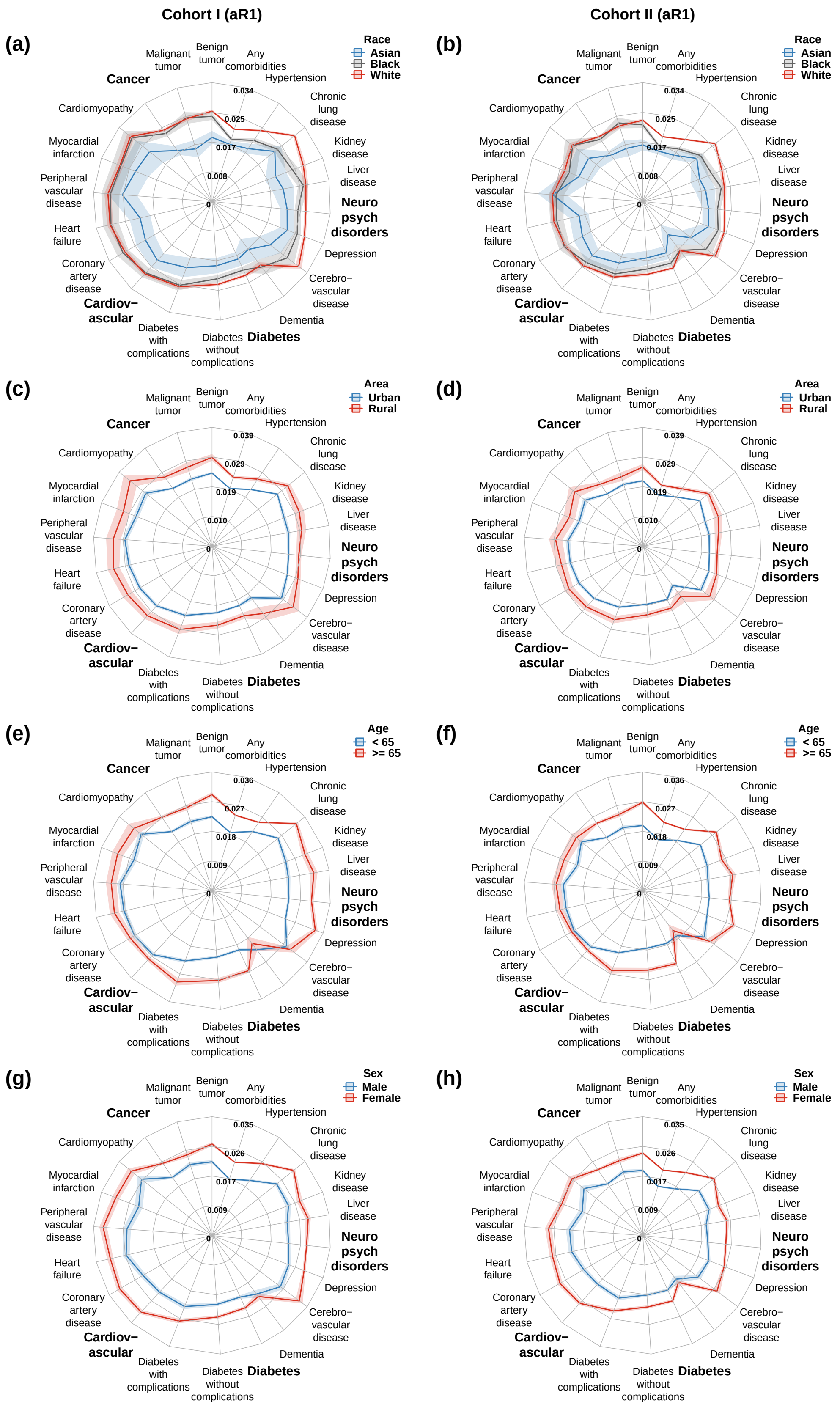


Figure S5: Estimated $aR_1$ of Long COVID associated with preexisting comorbidities across subpopulations, with 95% CIs.

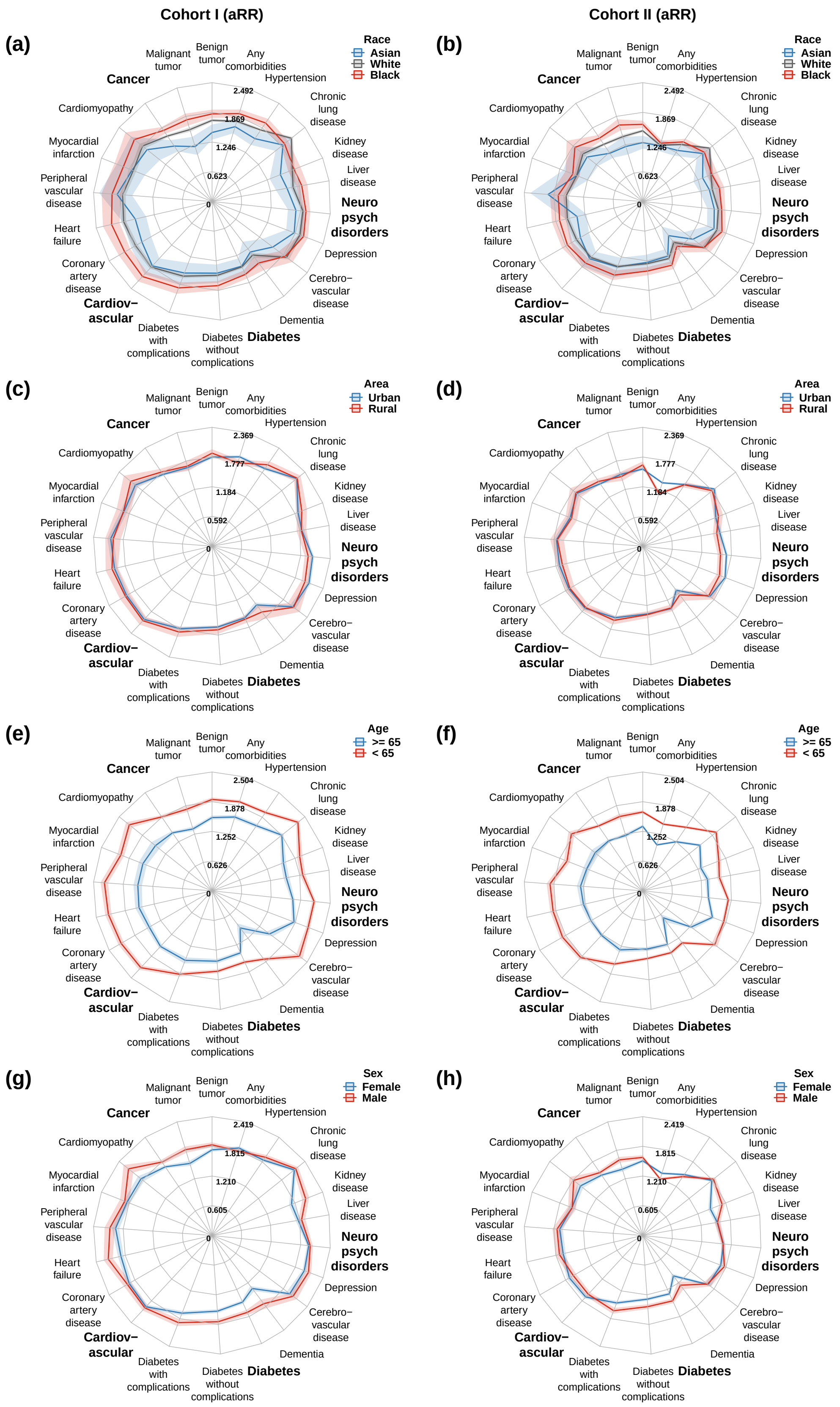


Figure S6: Estimated aRRs of Long COVID associated with preexisting comorbidities across subpopulations, with 95% CIs.

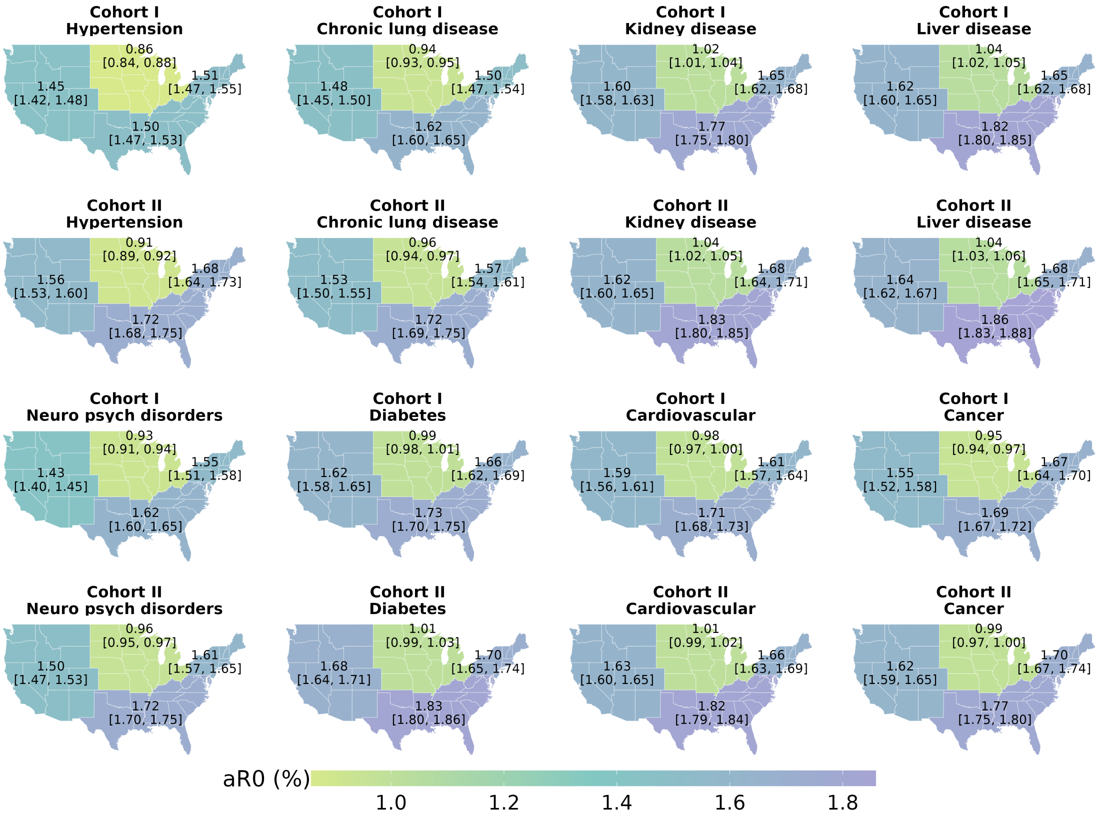


**(a)** $aR_0$

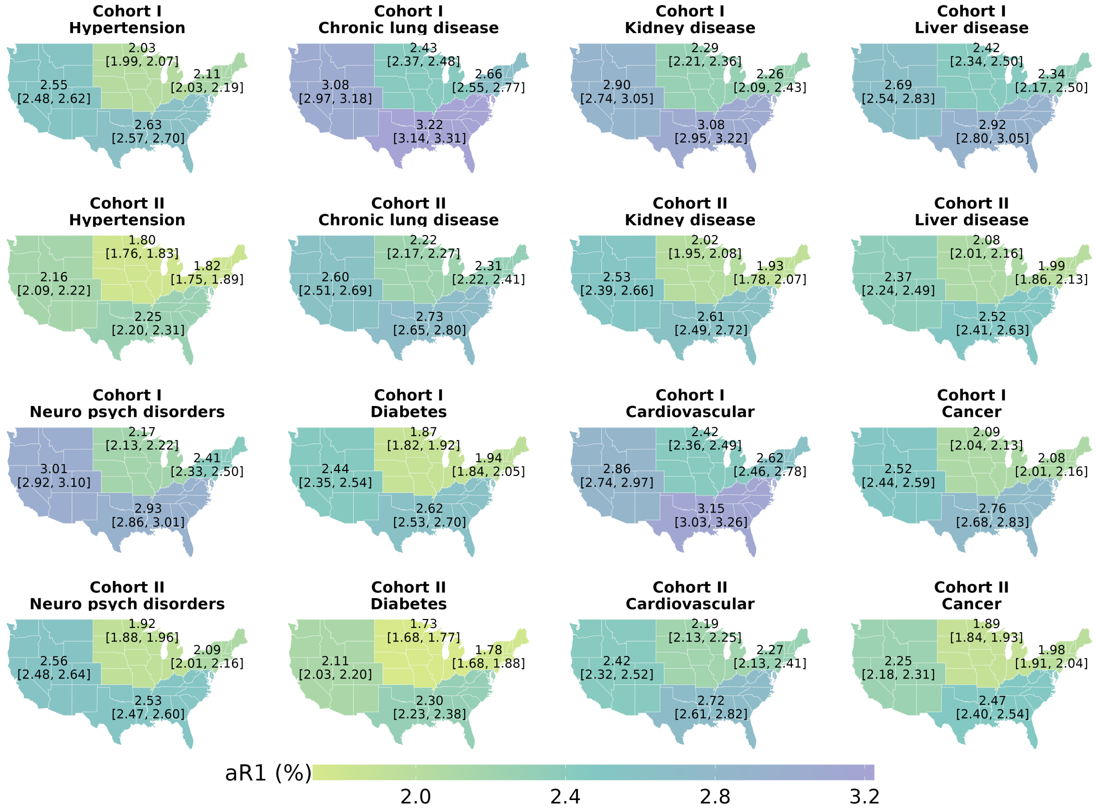


**(b)** $aR_1$

Figure S7: The risks of Long COVID associated with preexisting comorbidities across census regions, along with their 95% CIs.

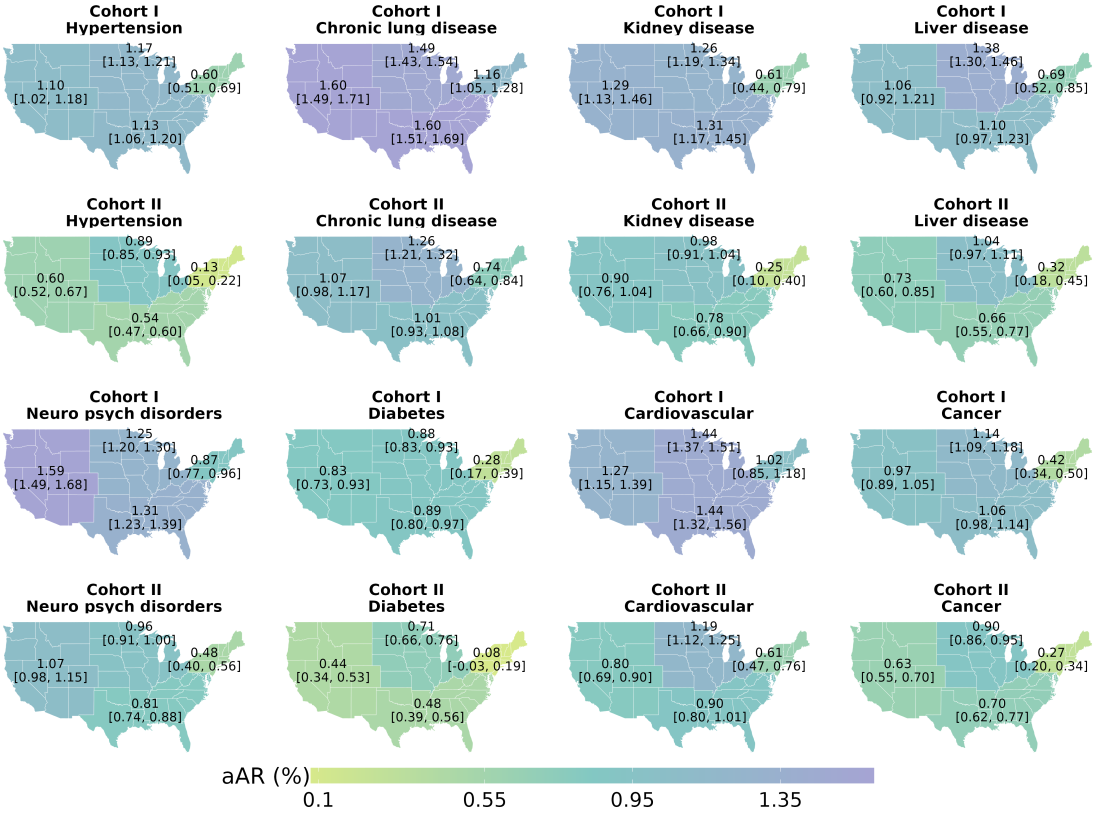


Figure S8: The aARs of Long COVID associated with preexisting comorbidities across census regions, along with their 95% CIs.

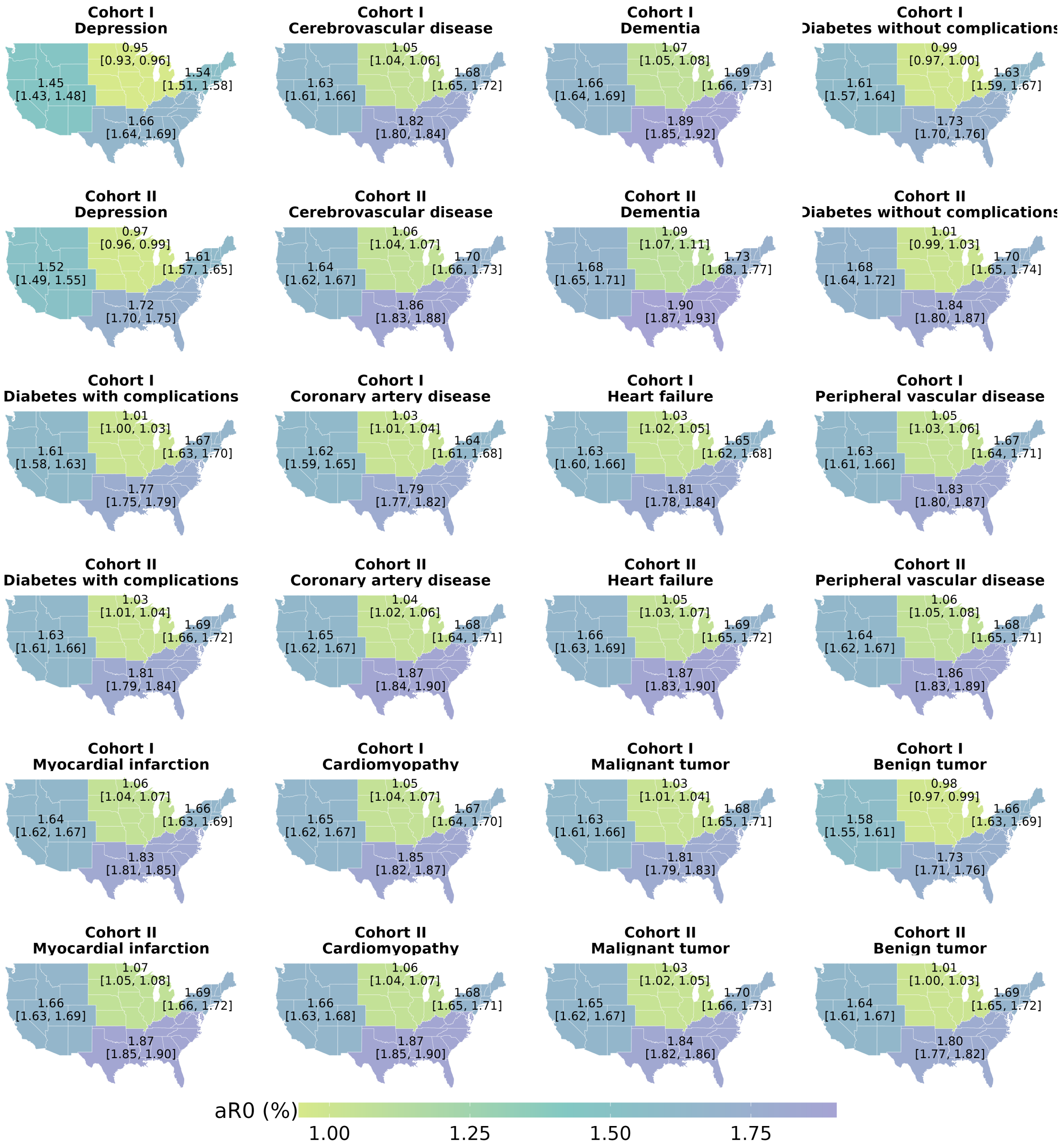


Figure S9: The $aR_0$ of Long COVID associated with the remaining 13 comorbidities across census regions, along with their 95% CIs.

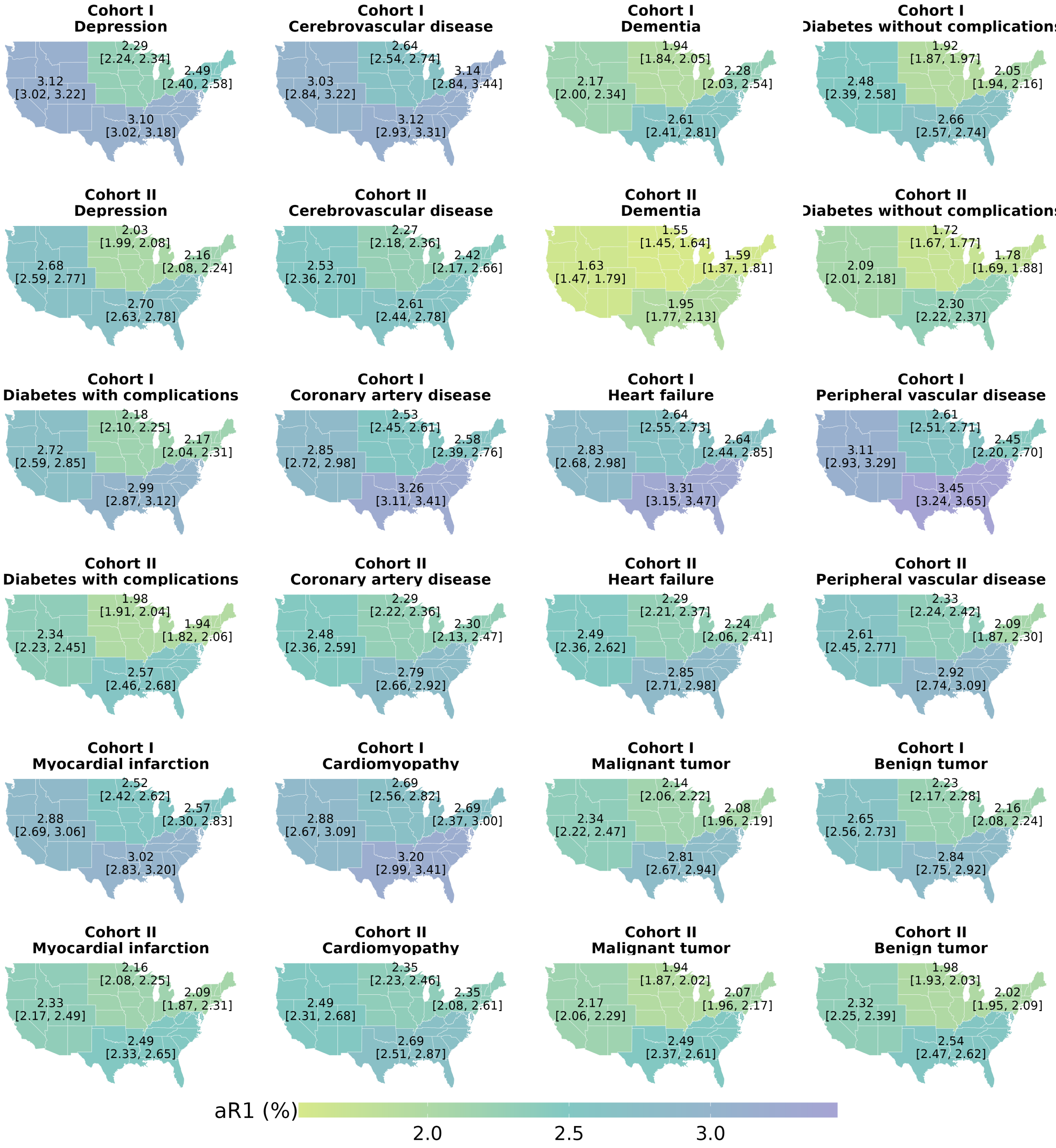


Figure S10: The $aR_1$ of Long COVID associated with the remaining 13 comorbidities across census regions, along with their 95% CIs.

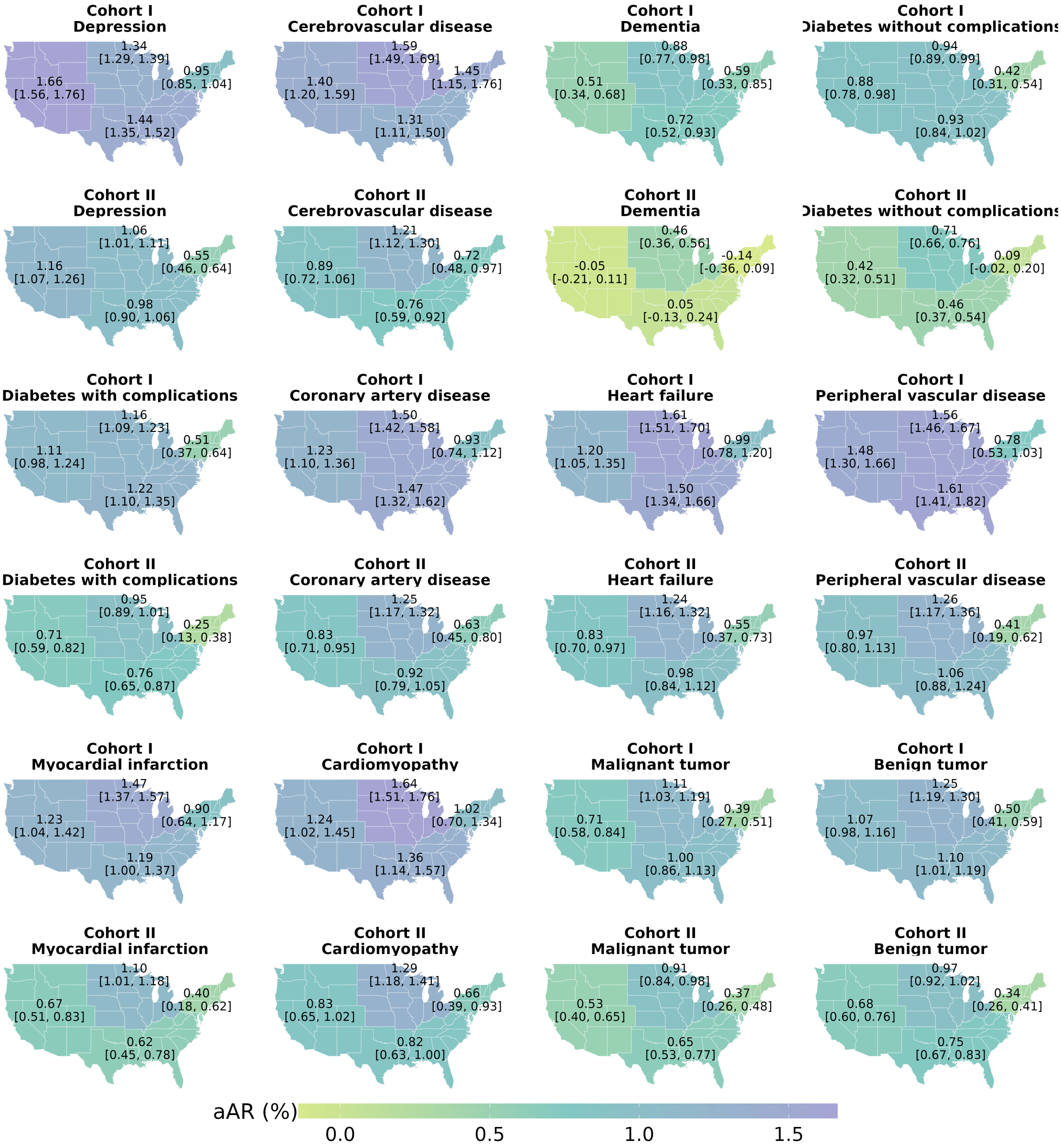


Figure S11: The aAR of Long COVID associated with the remaining 13 comorbidities across census regions, along with their 95% CIs.

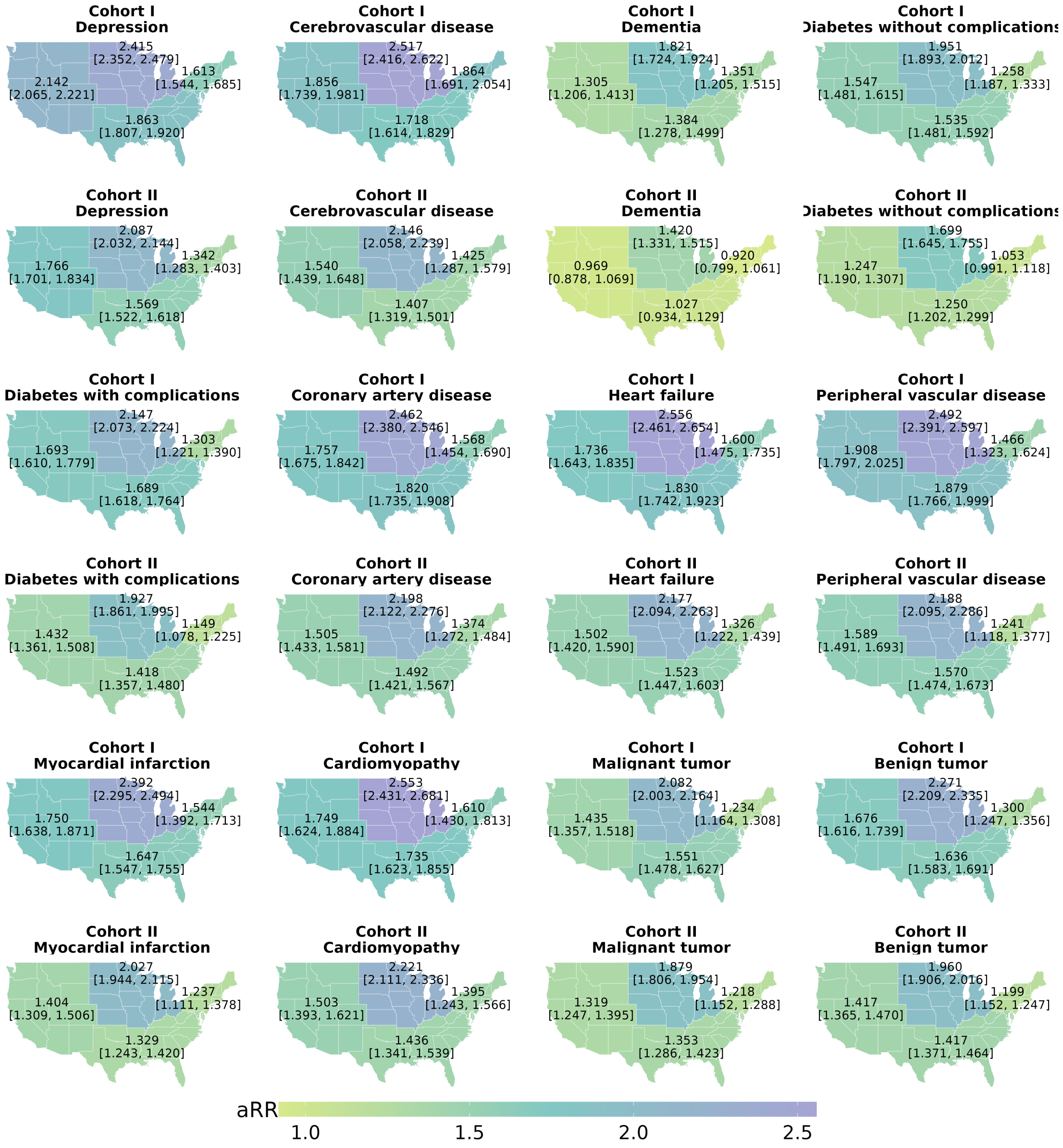


Figure S12: The aRR of Long COVID associated with the remaining 13 comorbidities across census regions, along with their 95% CIs.

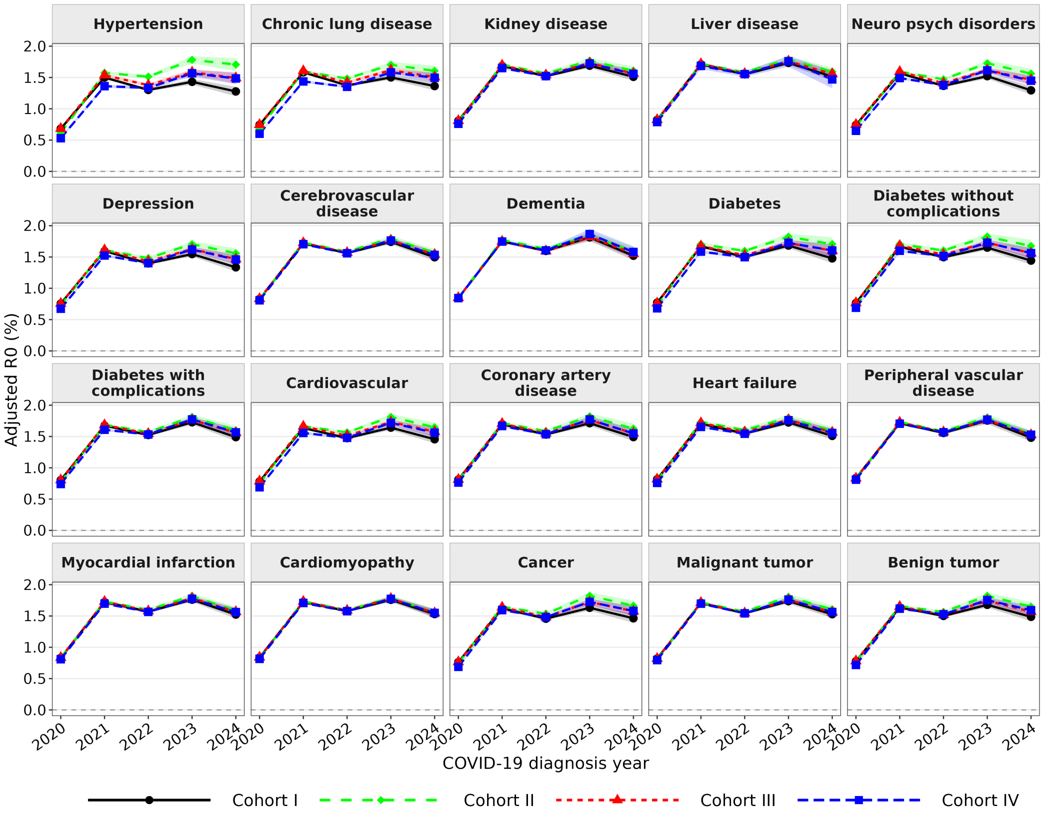


Figure S13: Temporal trends in $aR_0$ for LC, with the 95% CIs.

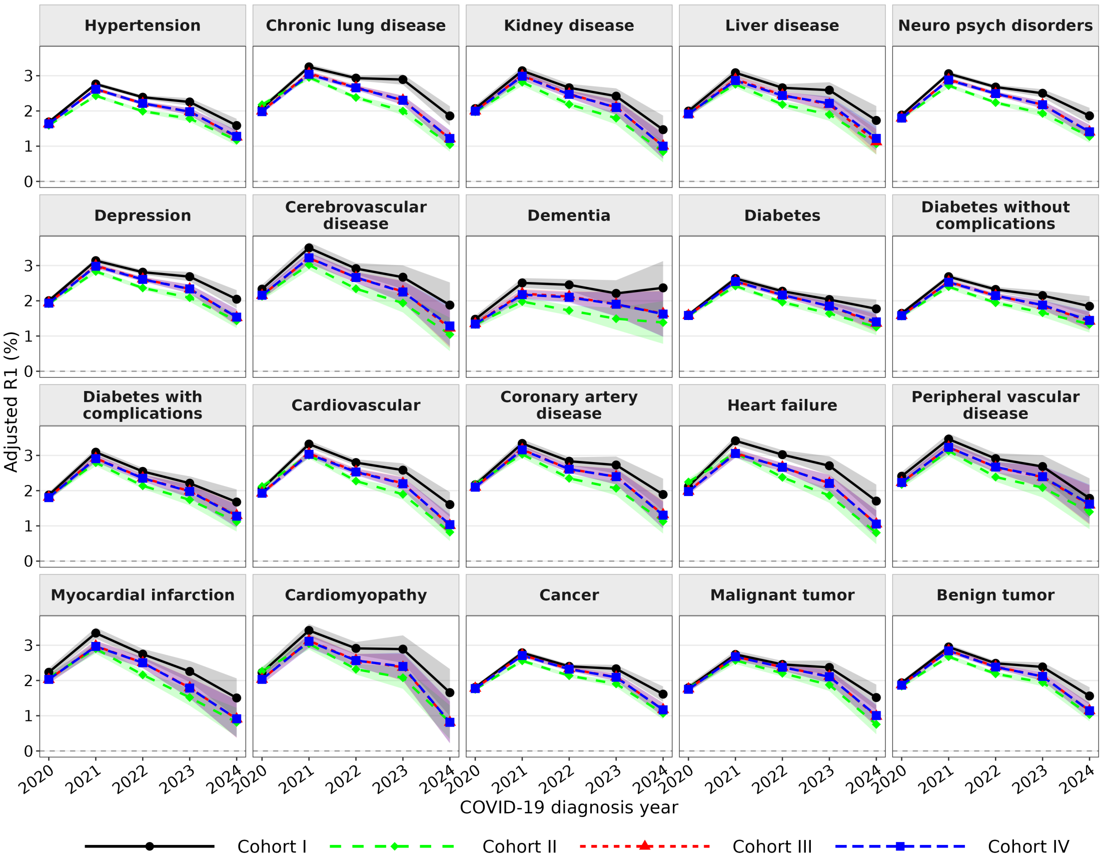


Figure S14: Temporal trends in $aR_1$ for LC, with the 95% CIs.

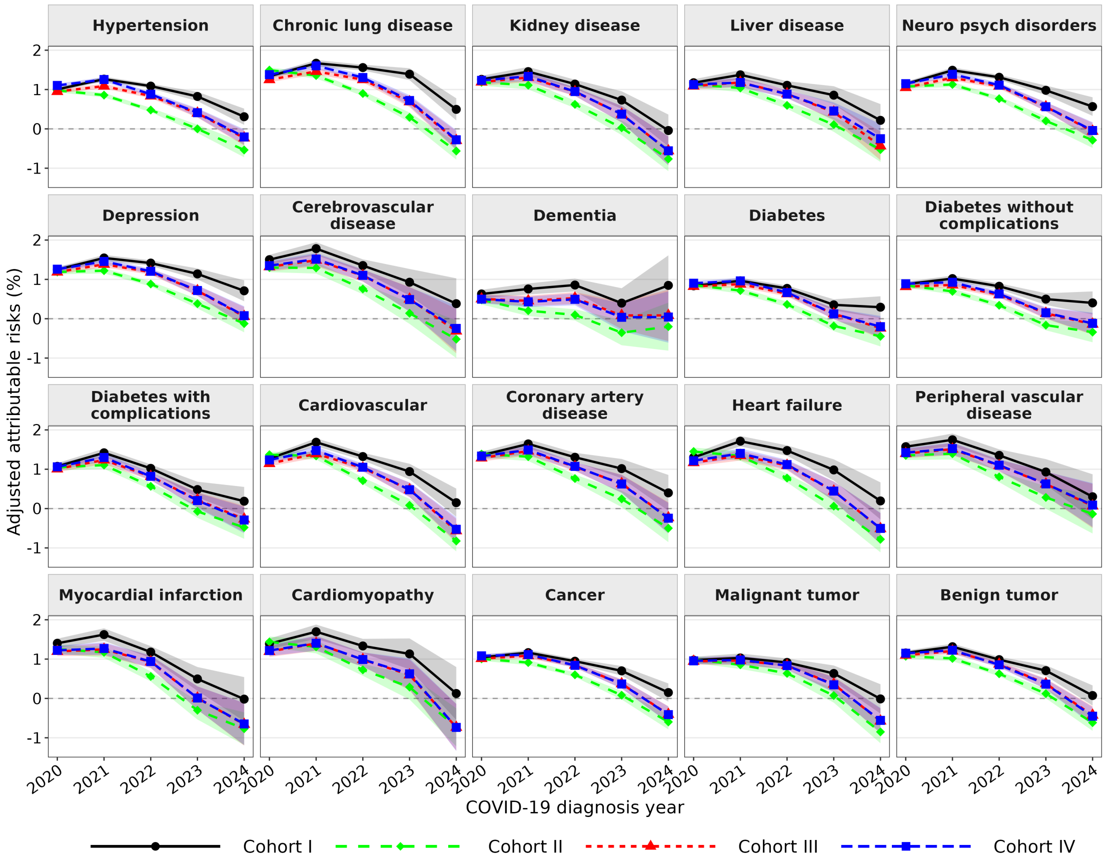


Figure S15: Temporal trends in aAR for LC, with the 95% CIs.

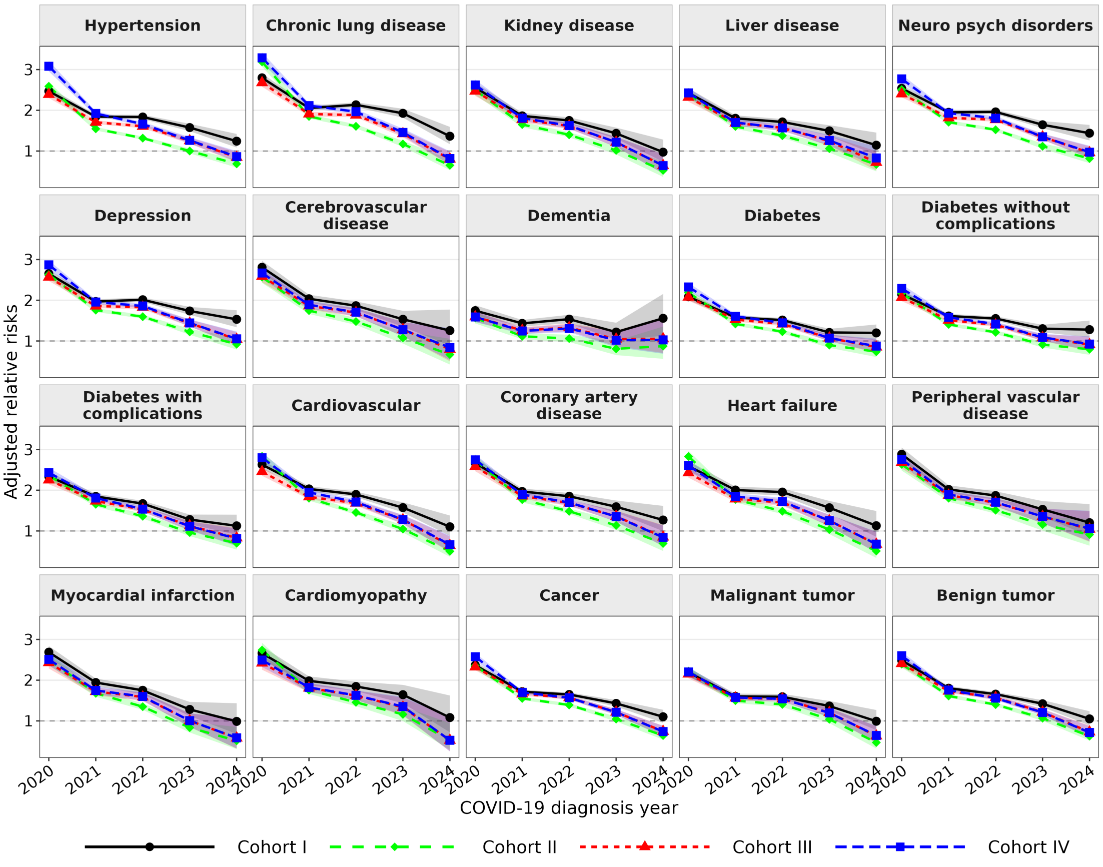


Figure S16: Temporal trends in aRR for LC, with the 95% CIs.

### S6 Model diagnosis

In this observational study, patients with and without a target comorbidity were generally not exchangeable at baseline with respect to the covariates, including age, region, calendar year of COVID-19 infection, other comorbidities, medication histories, and subgroup characteristics, as described in the manuscript and Supplementary **Table** S8. To address this, we employed a propensity score–based doubly robust framework to achieve covariate balance as closely as possible between exposed and unexposed groups. Specifically, exposure probabilities were estimated using cross-fitted ensemble models as described in **Algorithm** 1 of the main text. Covariate balance was then evaluated by calculating standardized mean differences (SMDs) before and after inverse probability weighting. The resulting unweighted and weighted SMDs were used to assess whether the propensity score model sufficiently improved comparability between exposed and unexposed groups, thereby supporting the interpretability of the adjusted risk estimates. The results are presented in **Figures** S17-S20 for Cohorts I-IV. Furthermore, we assessed propensity score overlap to evaluate the positivity assumption in **Figures** S21-S22 for Cohorts I-IV.

#### S6.1 Covariate balance

##### S6.1.1 Cohort I

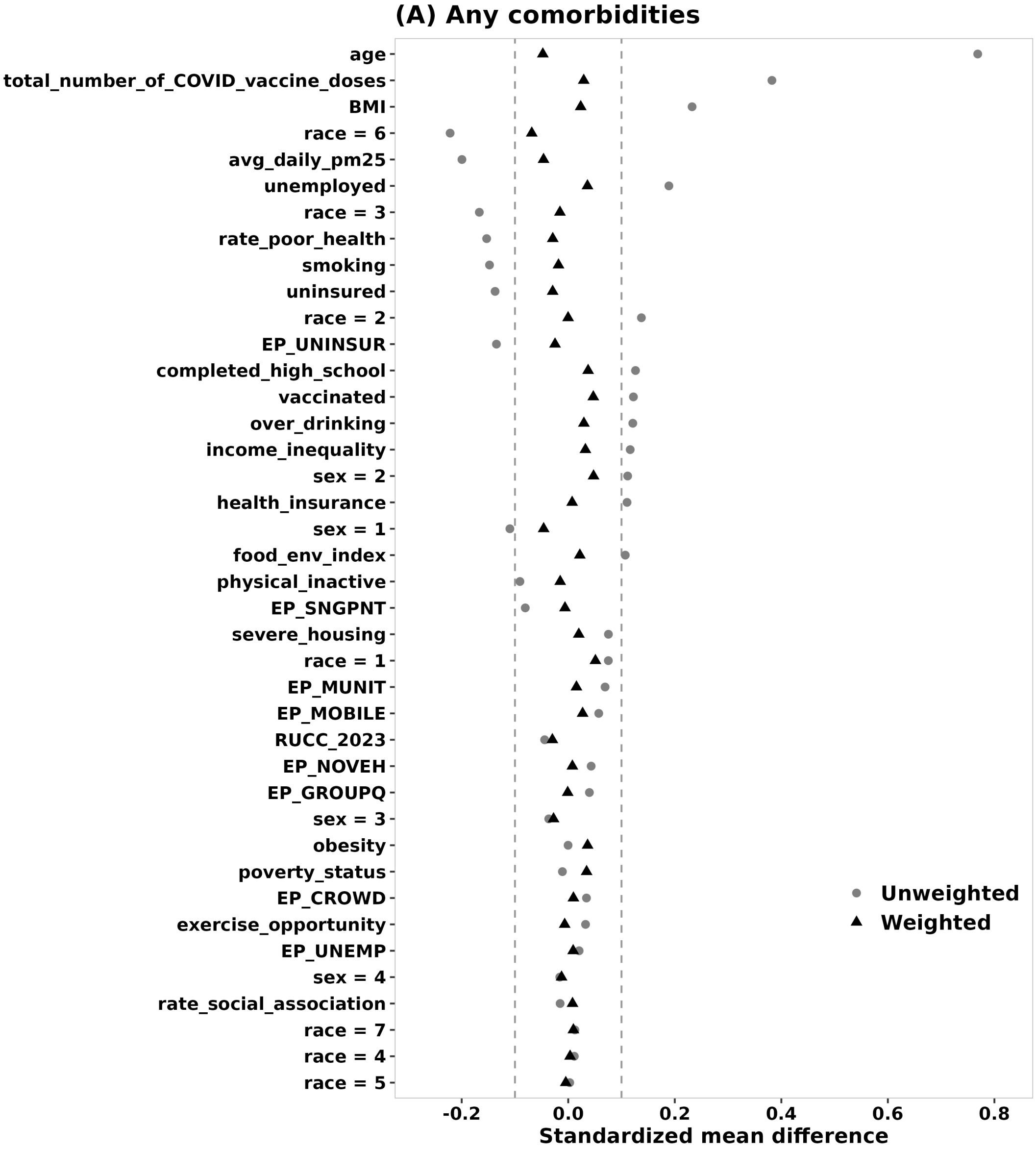


Figure S17: Covariate balance before and after inverse probability weighting under Cohort I.

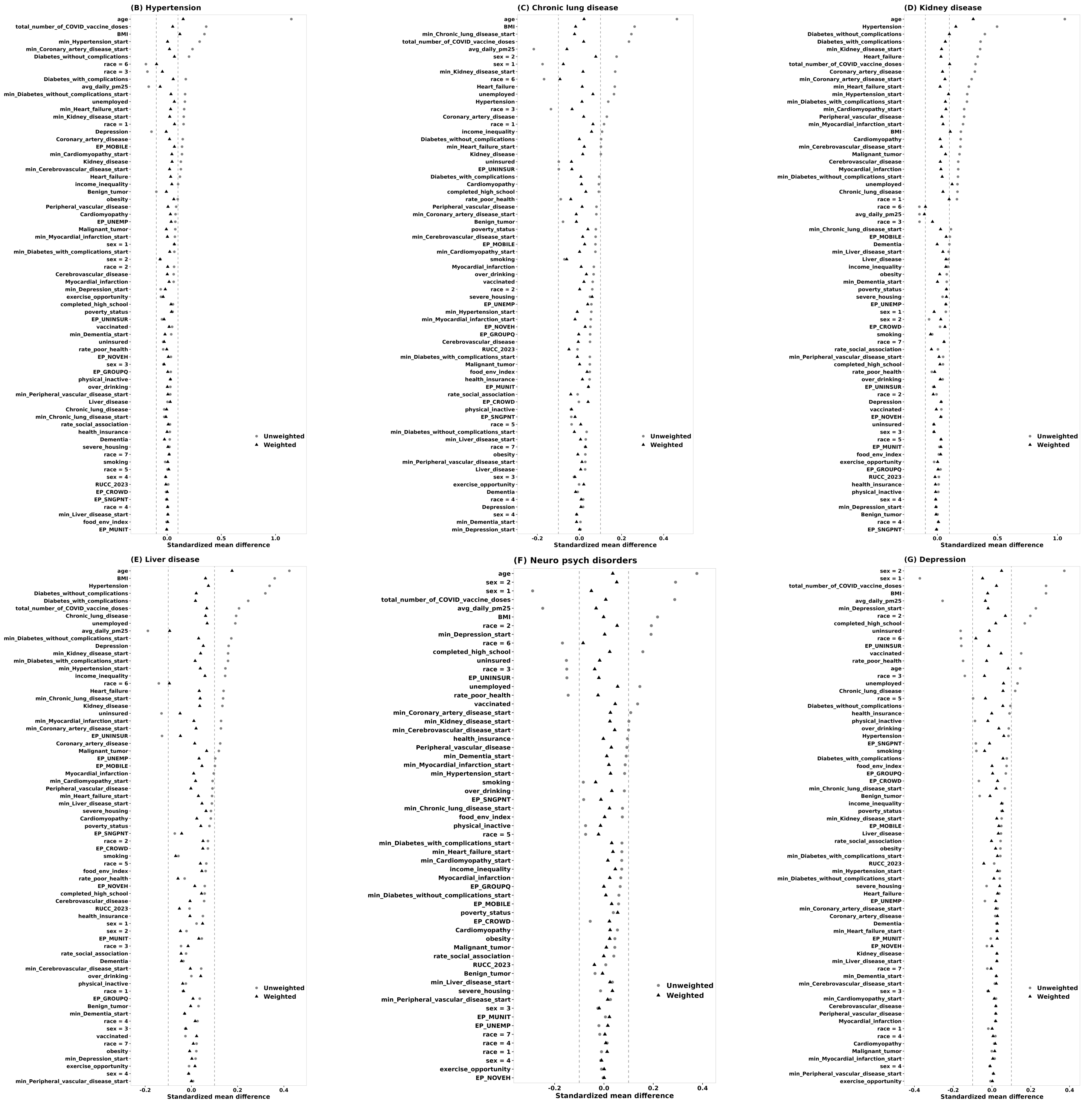

Figure S17: Covariate balance under Cohort I (continued).

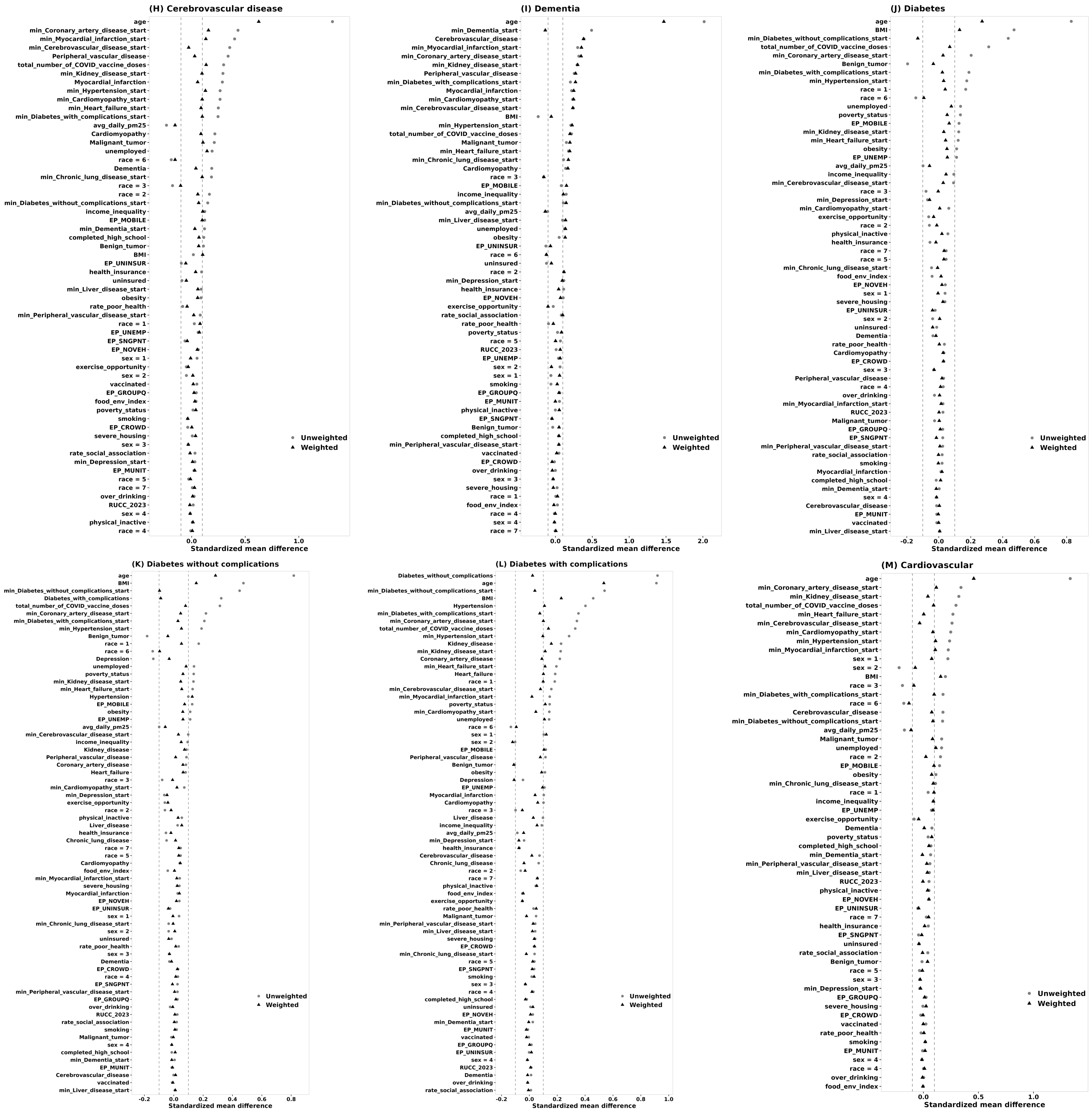


Figure S17: Covariate balance under Cohort I (continued).

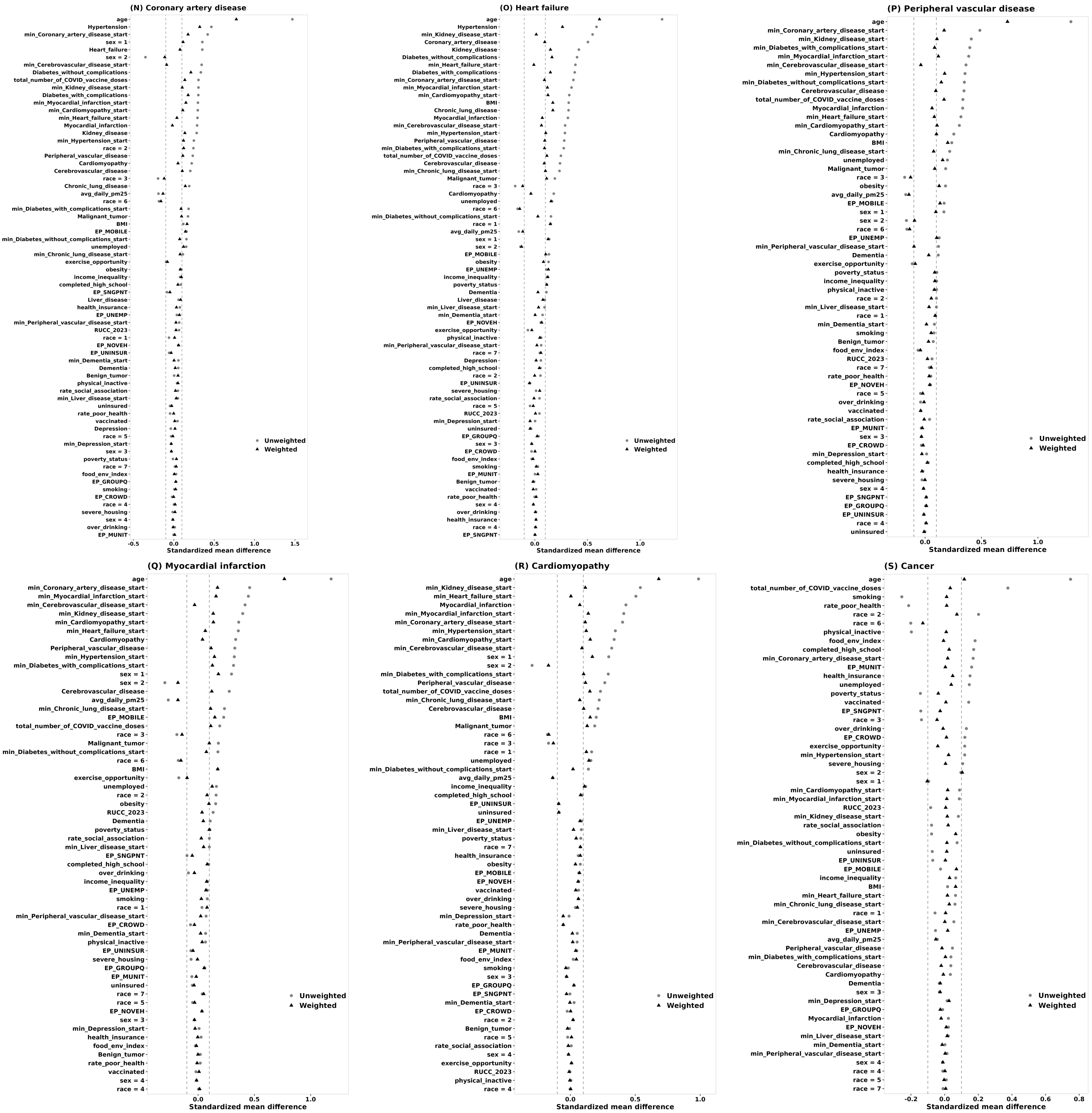


Figure S17: Covariate balance under Cohort I (continued).

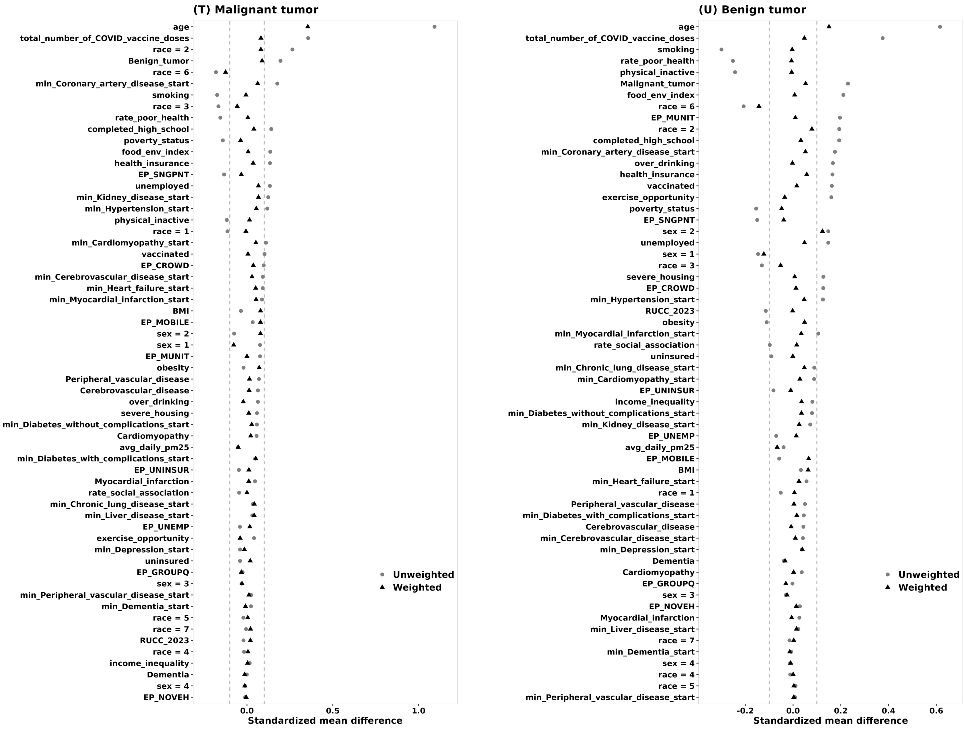


Figure S17: Covariate balance under Cohort I (continued).

##### S6.1.2 Cohort II

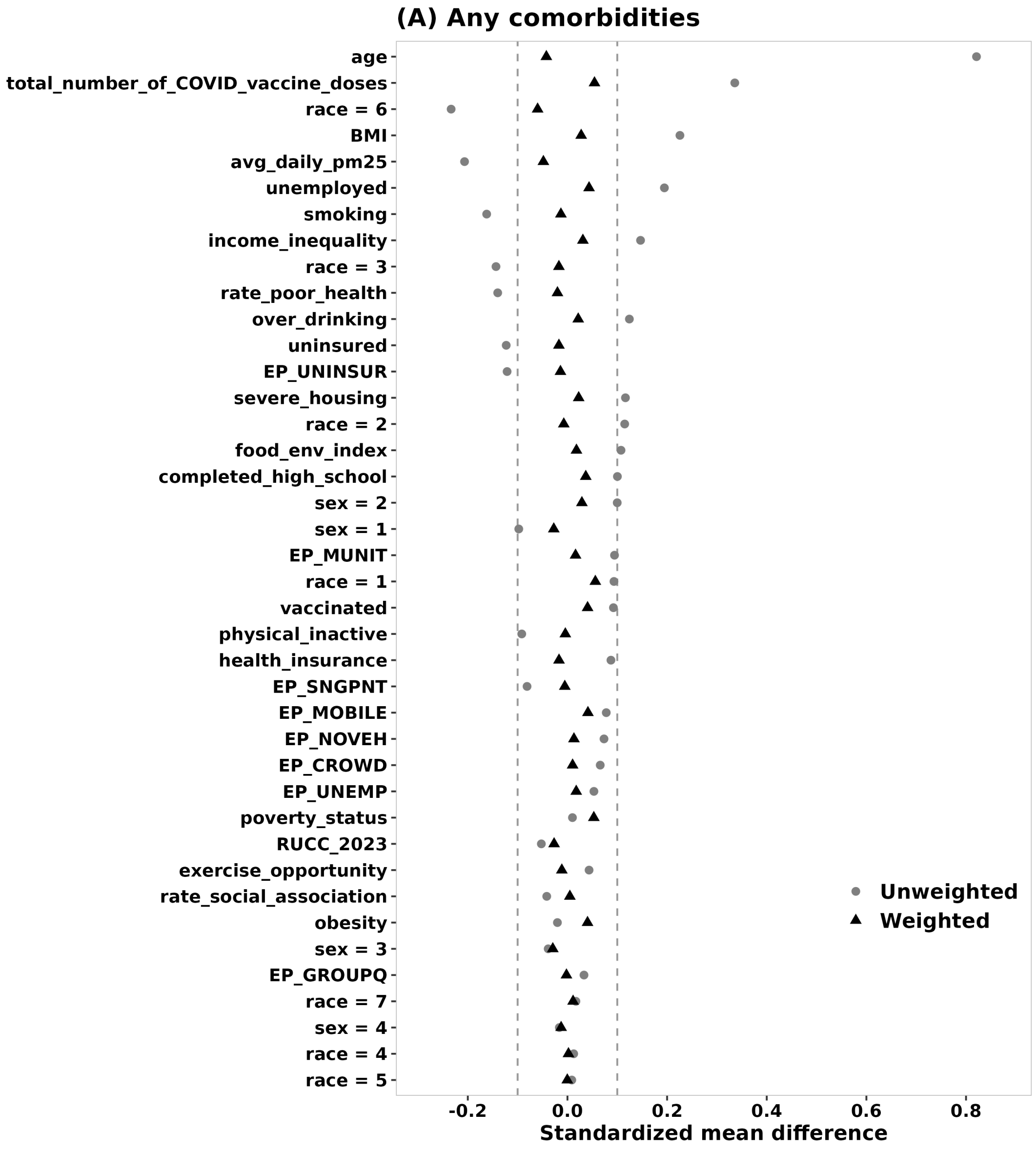


Figure S18: Covariate balance before and after inverse probability weighting under Cohort II.

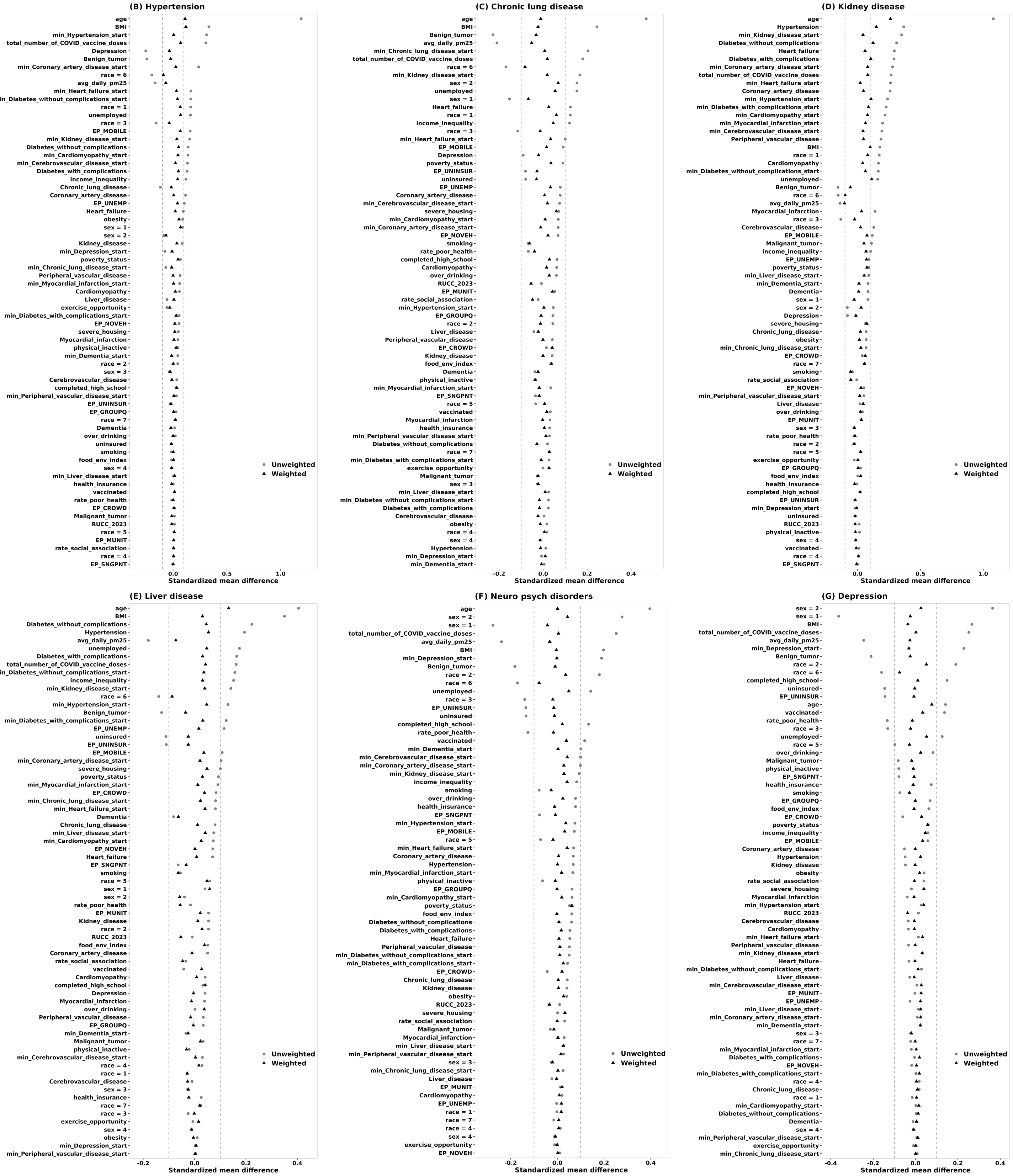

Figure S18: Covariate balance under Cohort II (continued).

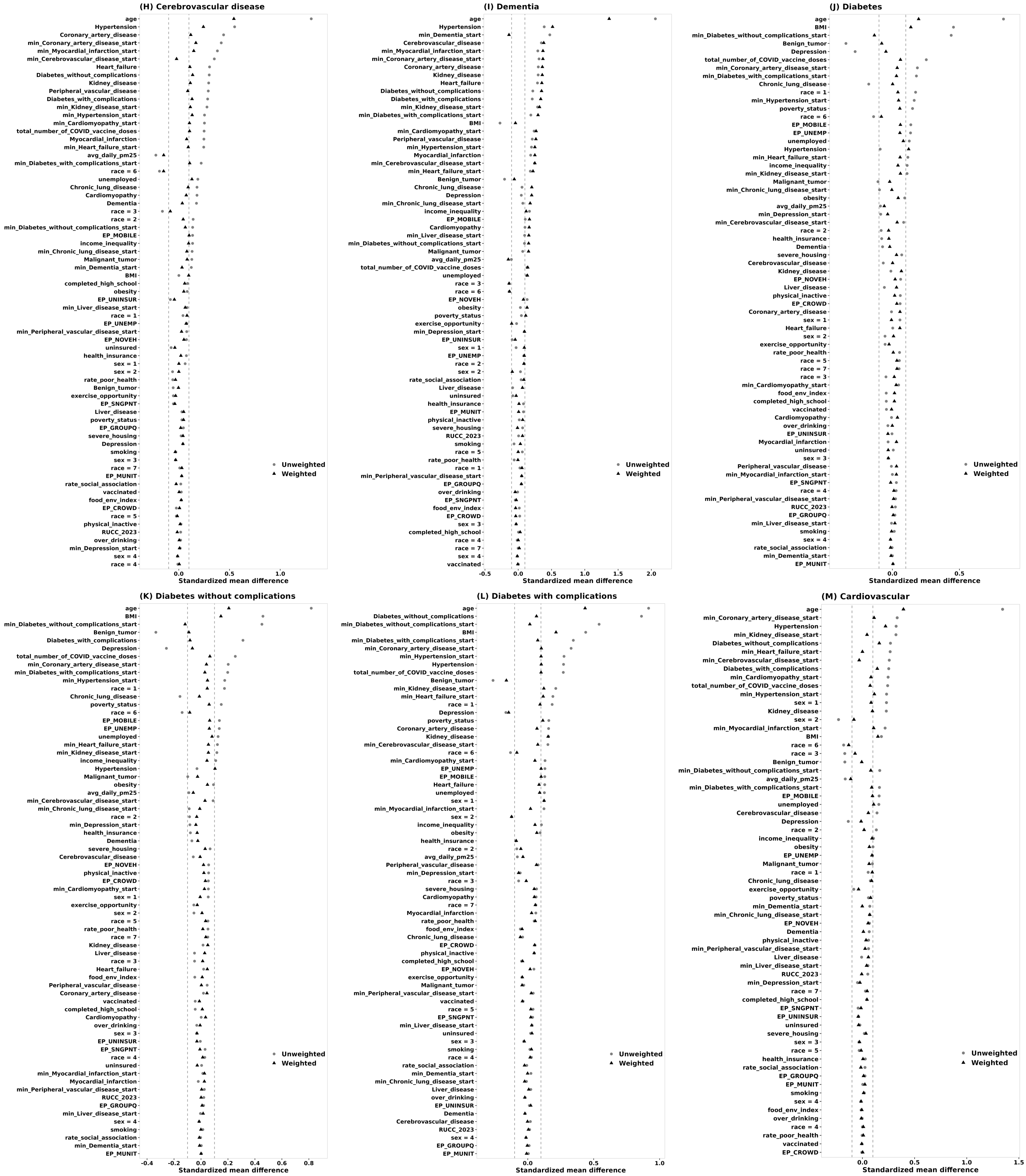


Figure S18: Covariate balance under Cohort II (continued).

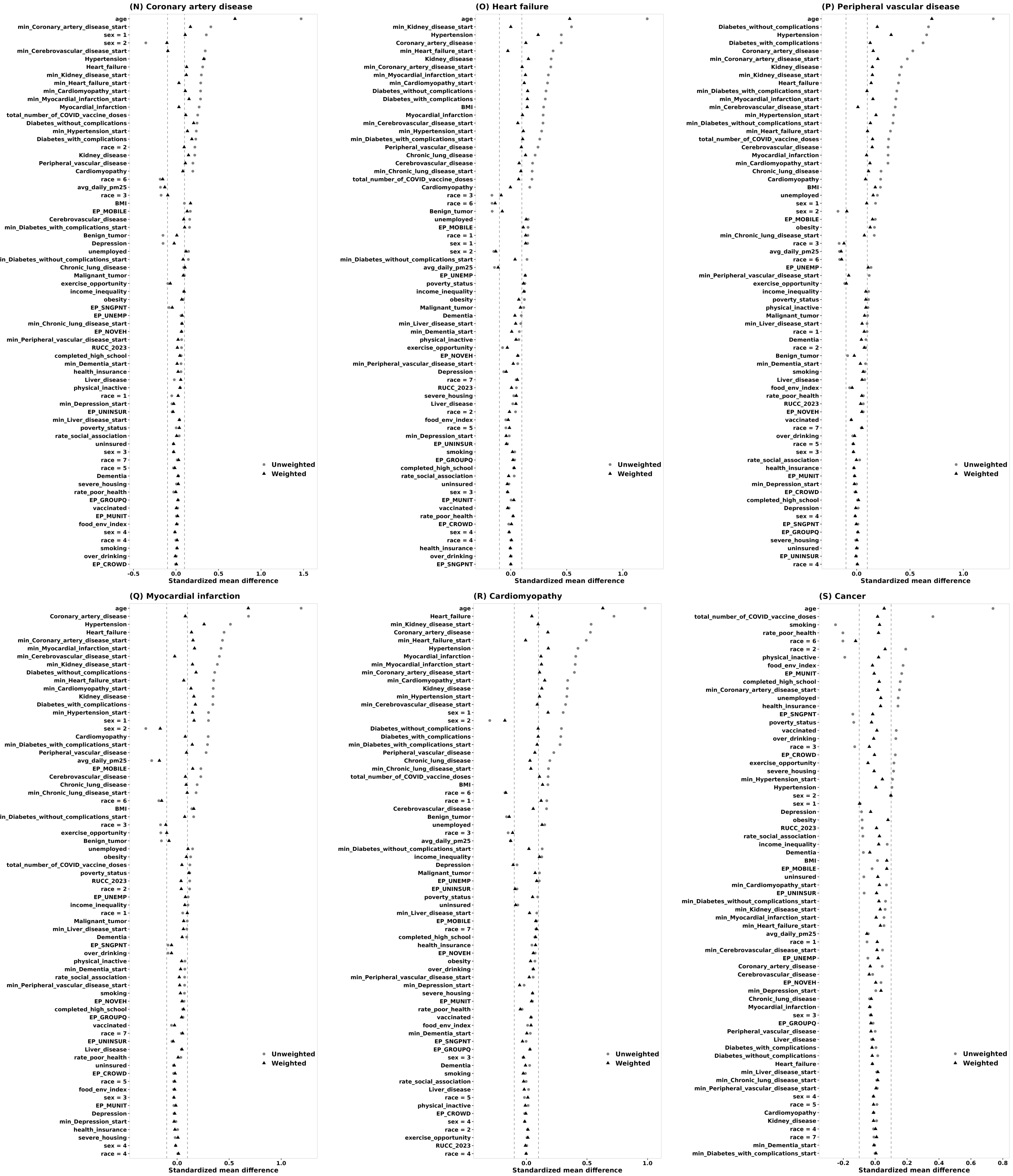


Figure S18: Covariate balance under Cohort II (continued).

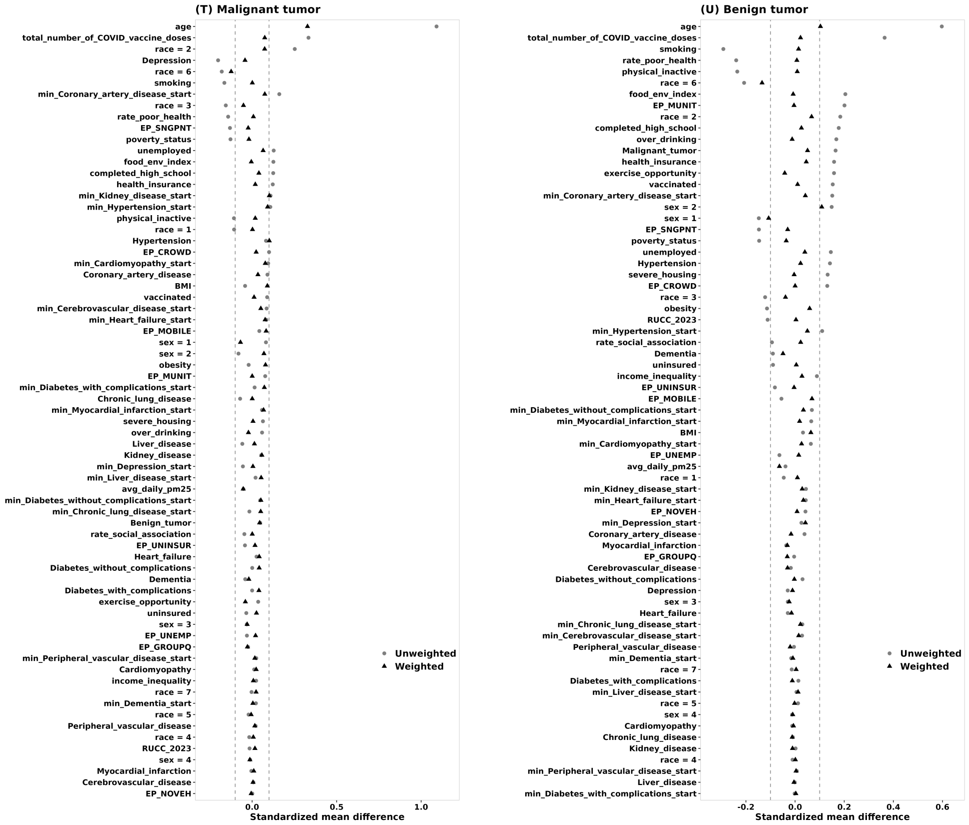


Figure S18: Covariate balance under Cohort II (continued).

##### S6.1.3 Cohort III

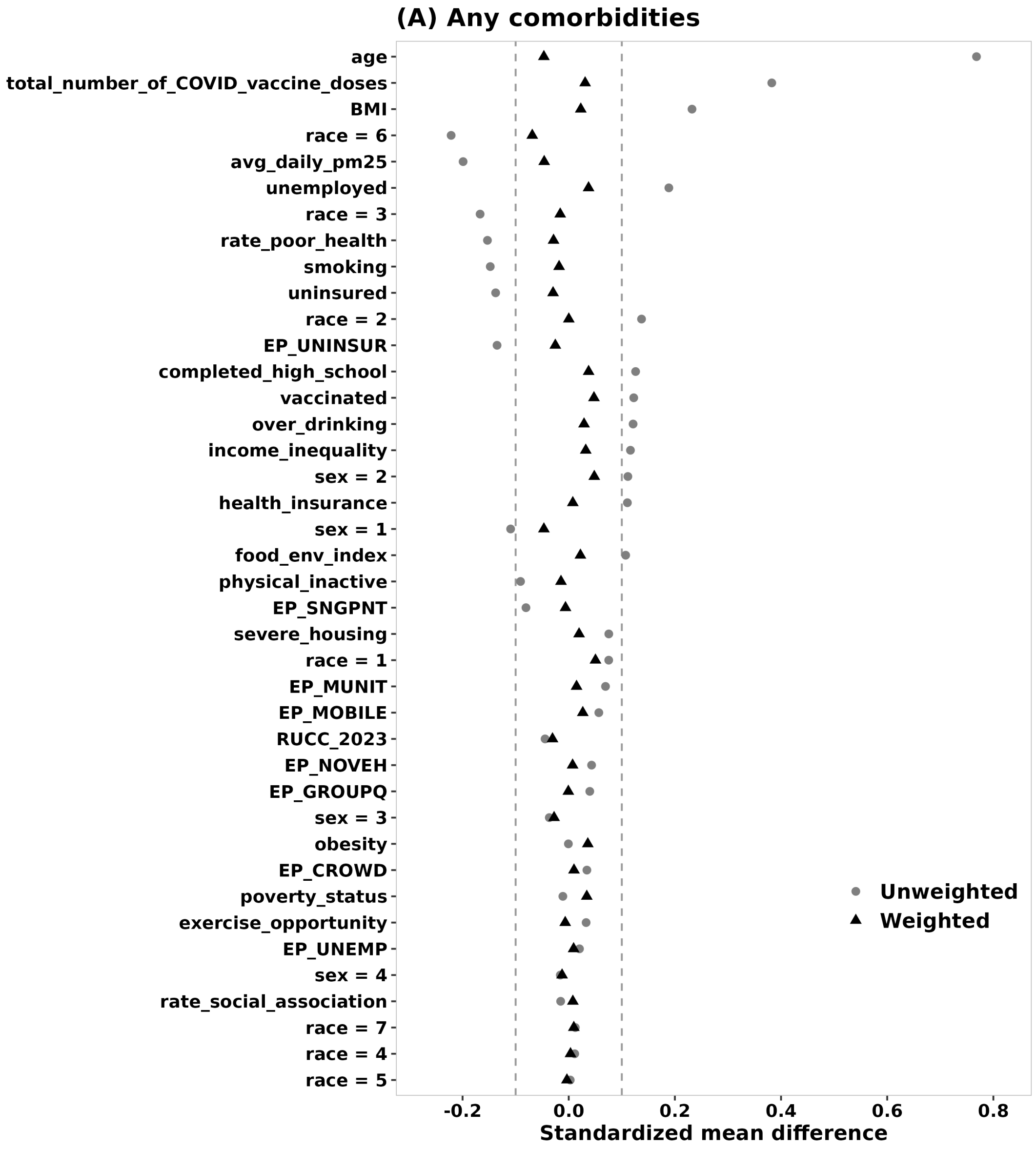


Figure S19: Covariate balance before and after inverse probability weighting under Cohort III.

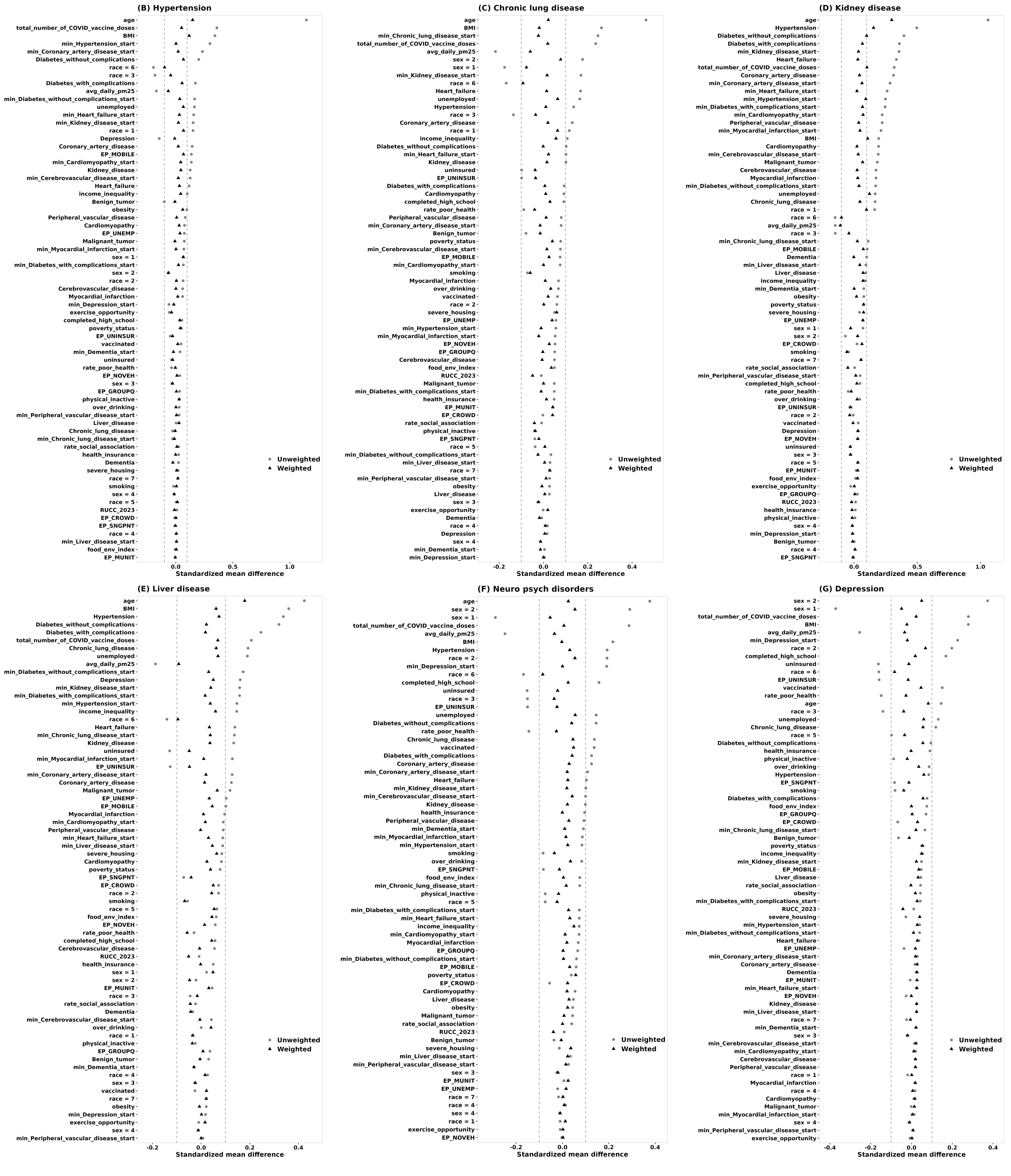

Figure S19: Covariate balance under Cohort III (continued).

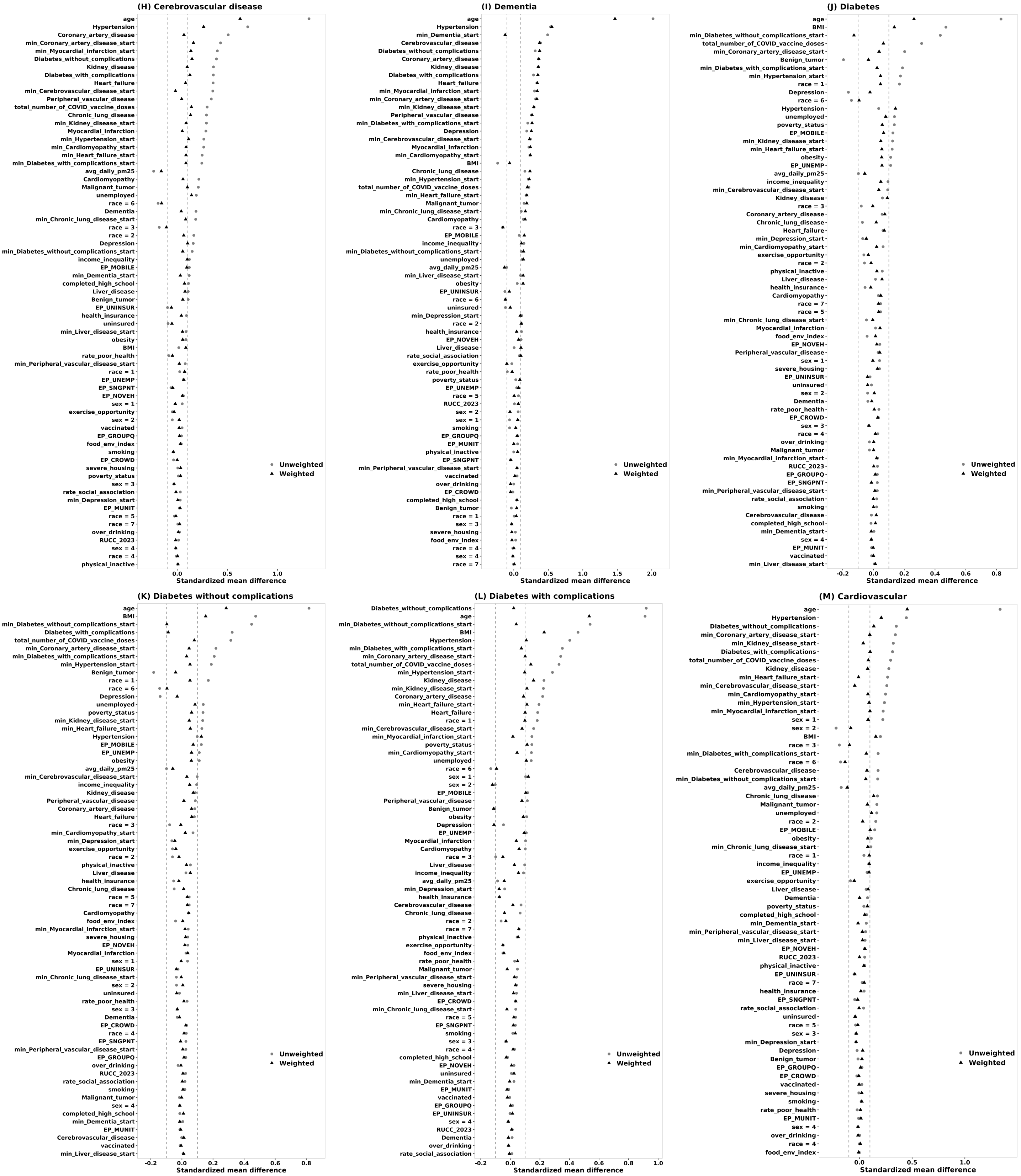


Figure S19: Covariate balance under Cohort III (continued).

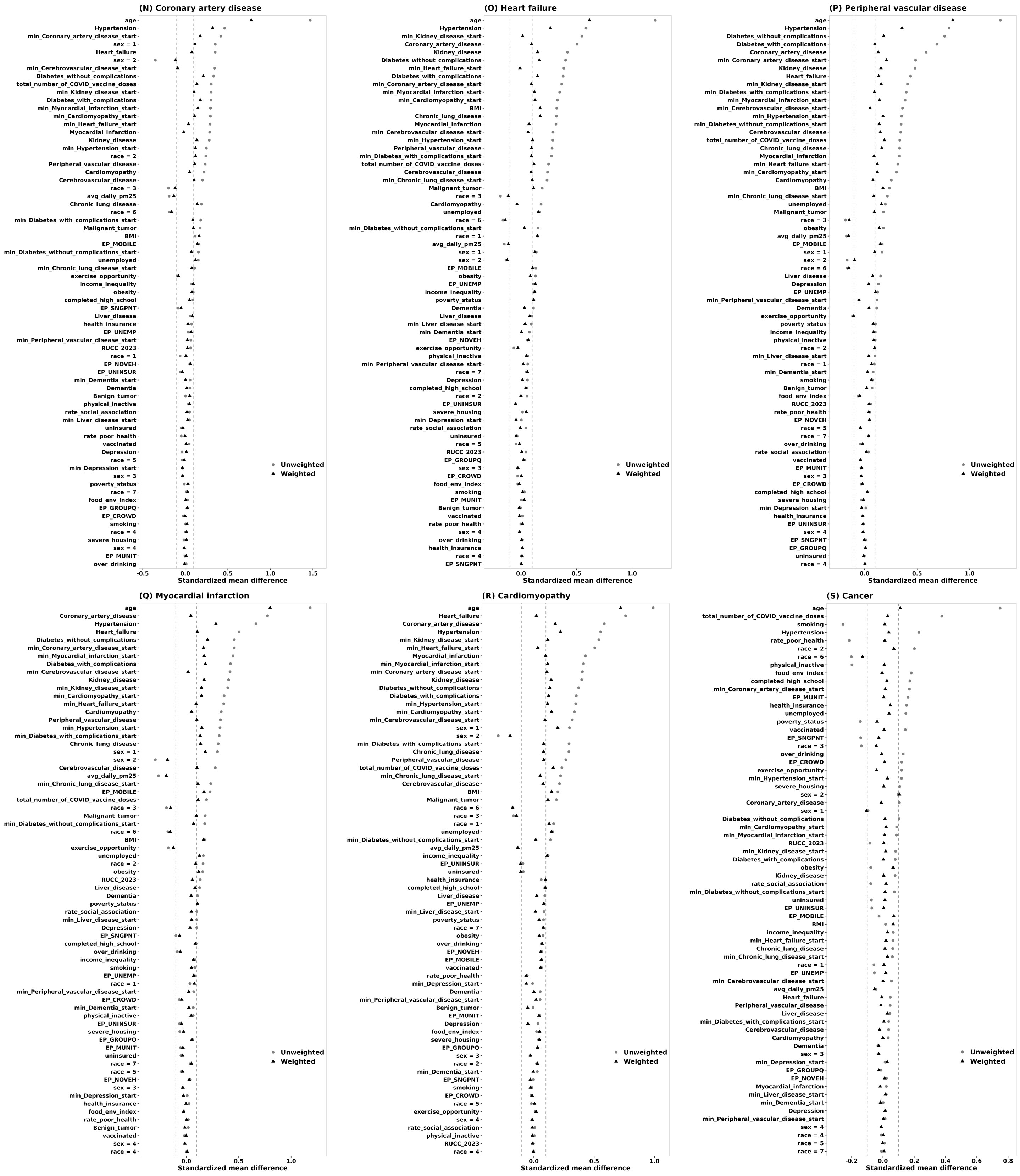


Figure S19: Covariate balance under Cohort III (continued).

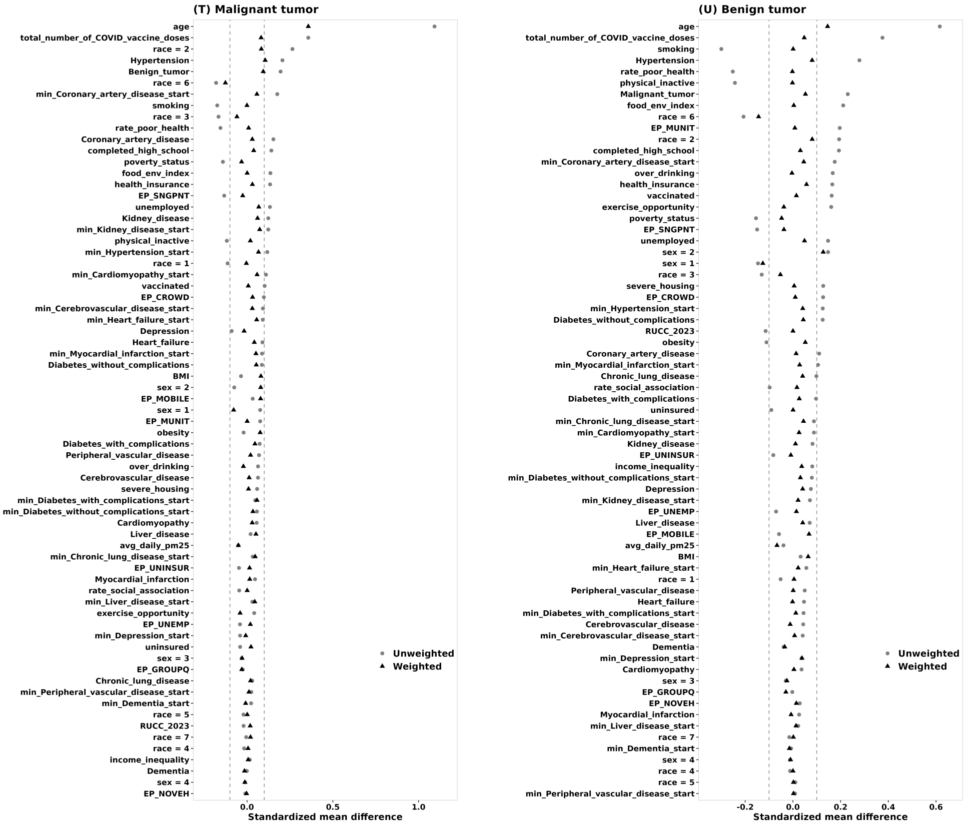


Figure S19: Covariate balance under Cohort III (continued).

##### S6.1.4 Cohort IV

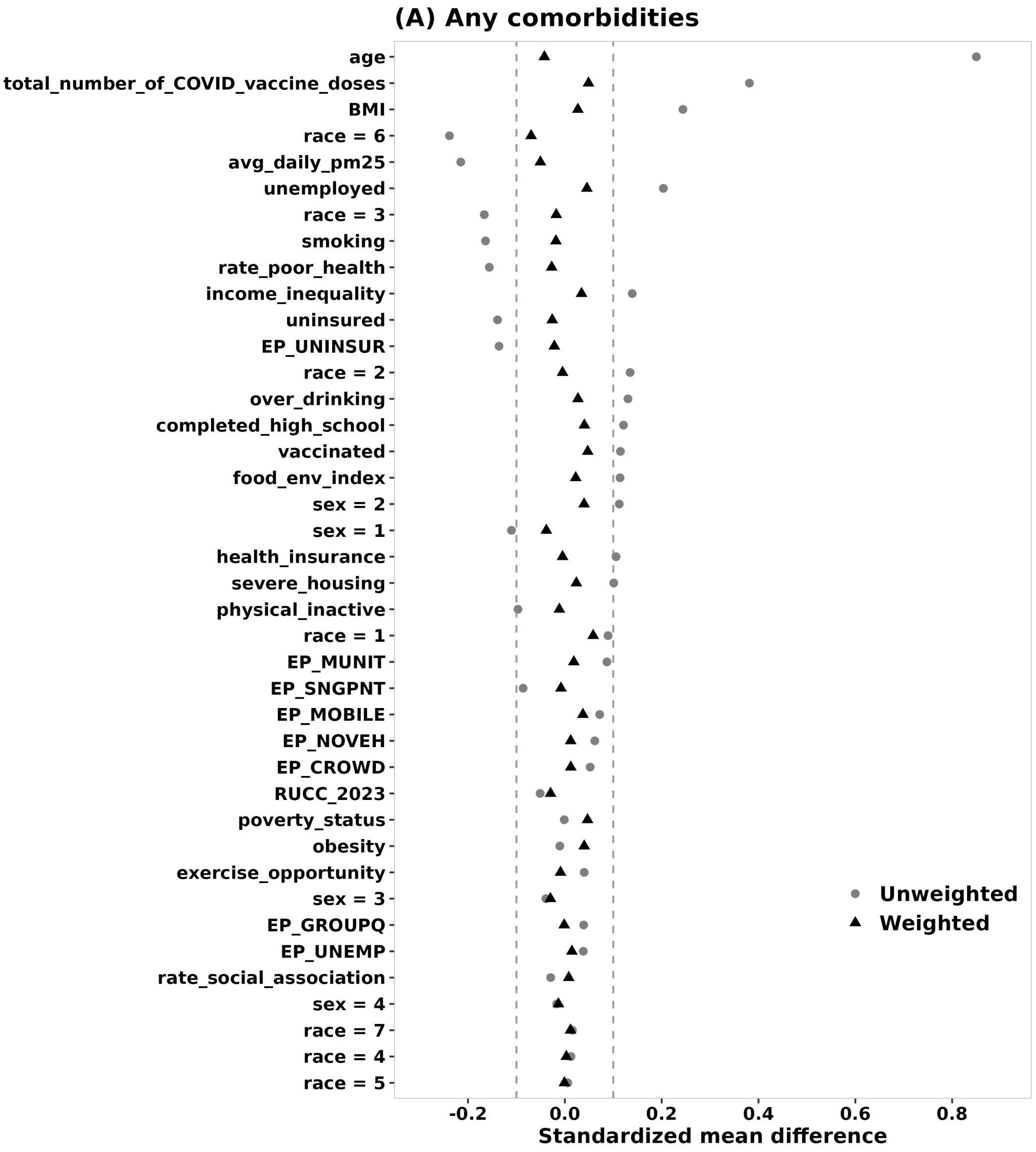


Figure S20: Covariate balance before and after inverse probability weighting under Cohort IV.

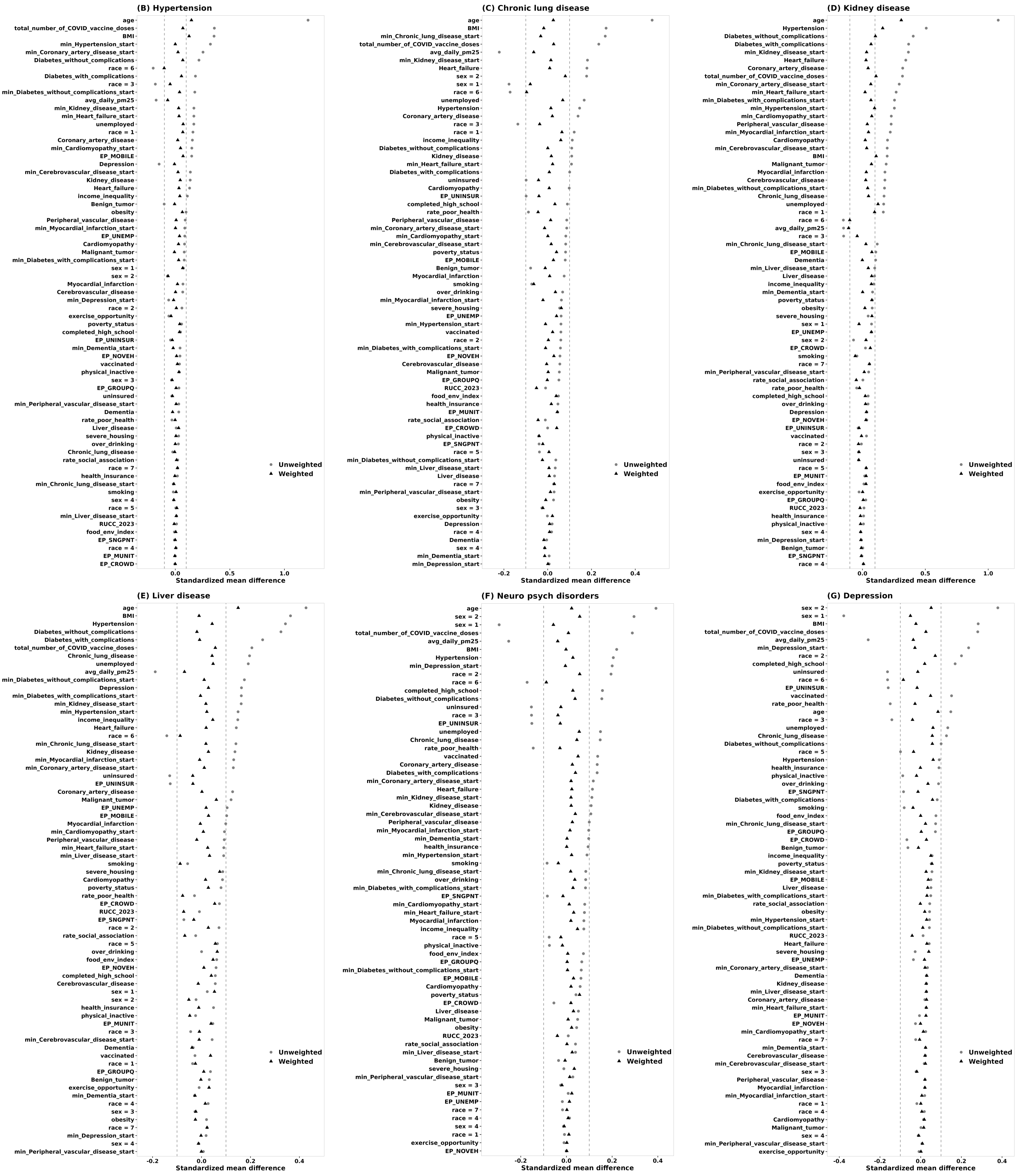

Figure S20: Covariate balance under Cohort IV (continued).

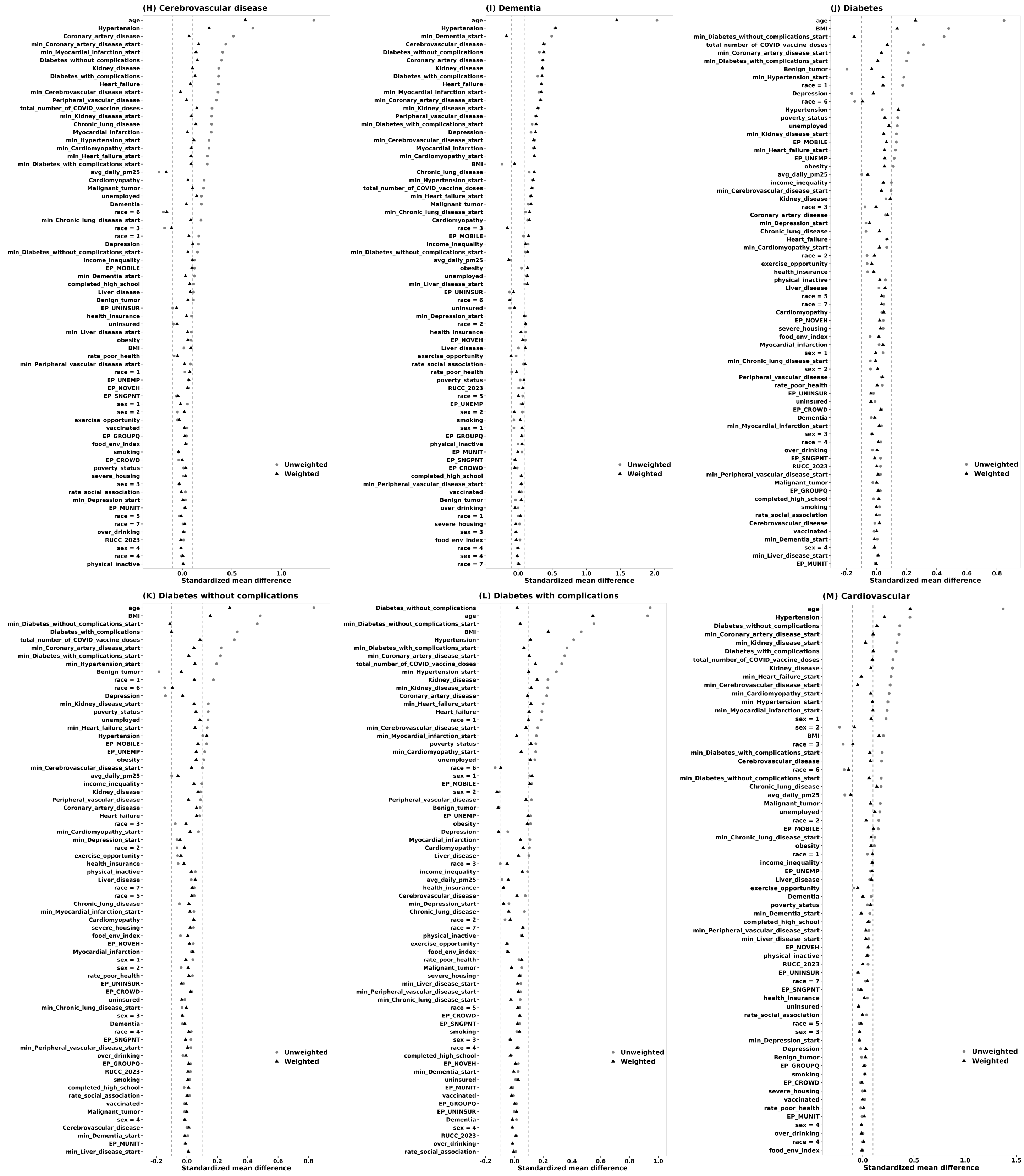


Figure S20: Covariate balance under Cohort IV (continued).

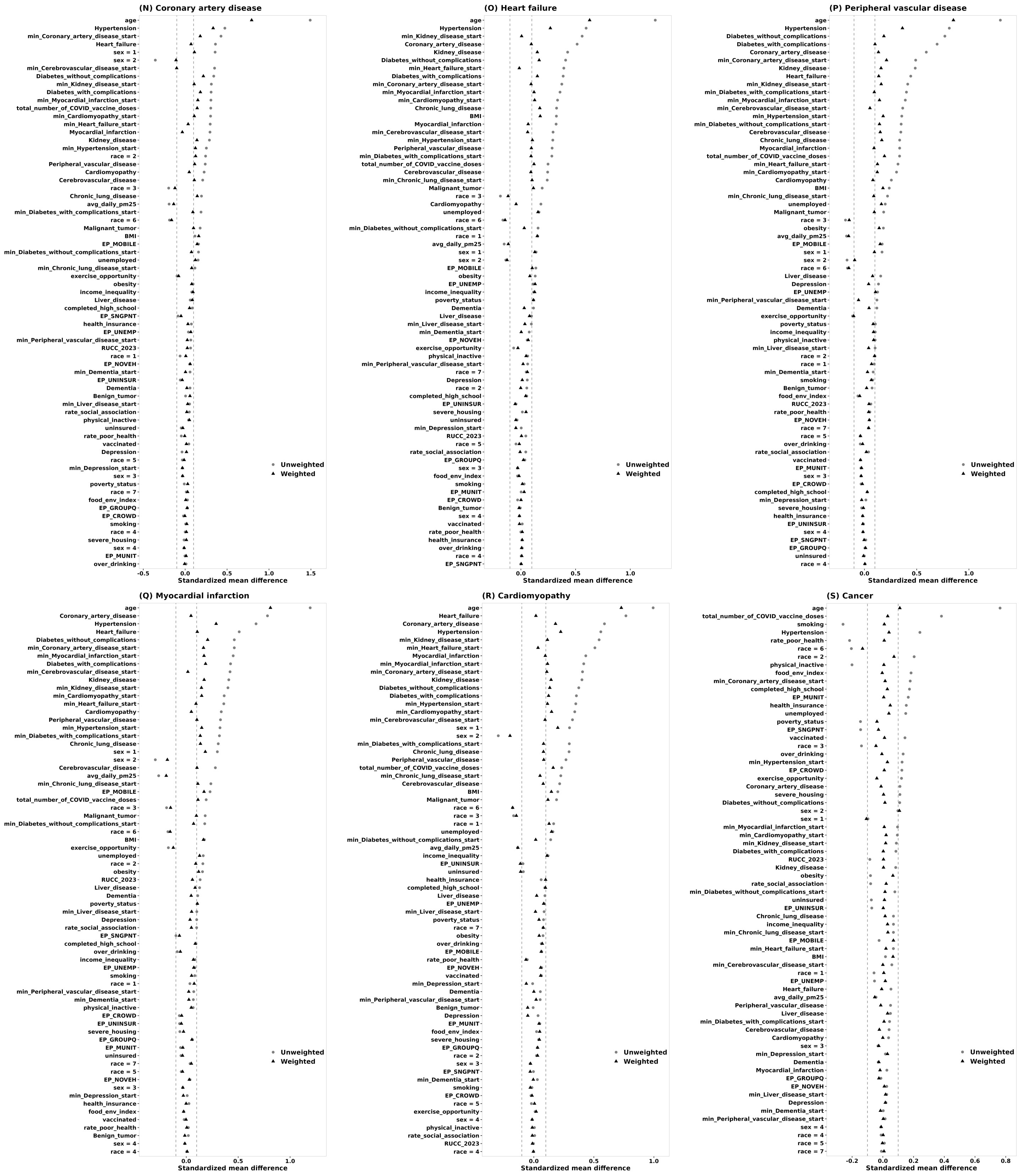


Figure S20: Covariate balance under Cohort IV (continued).

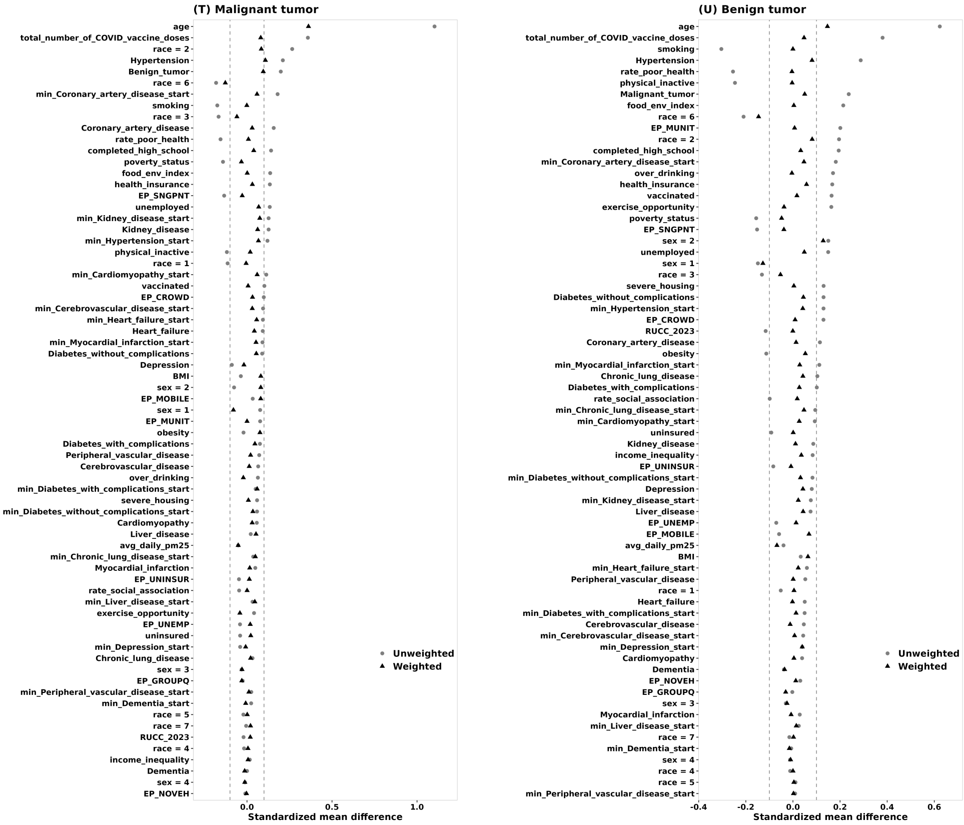


Figure S20: Covariate balance under Cohort IV (continued).

#### S6.2 Propensity overlap

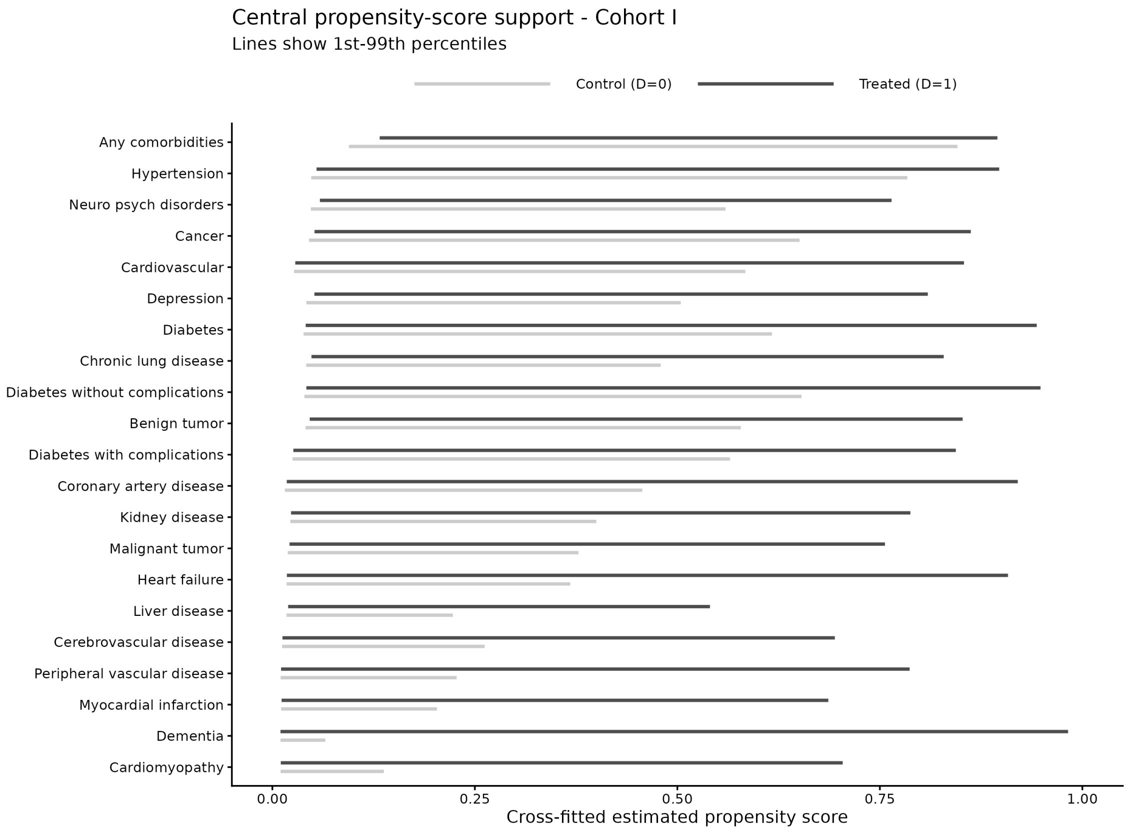


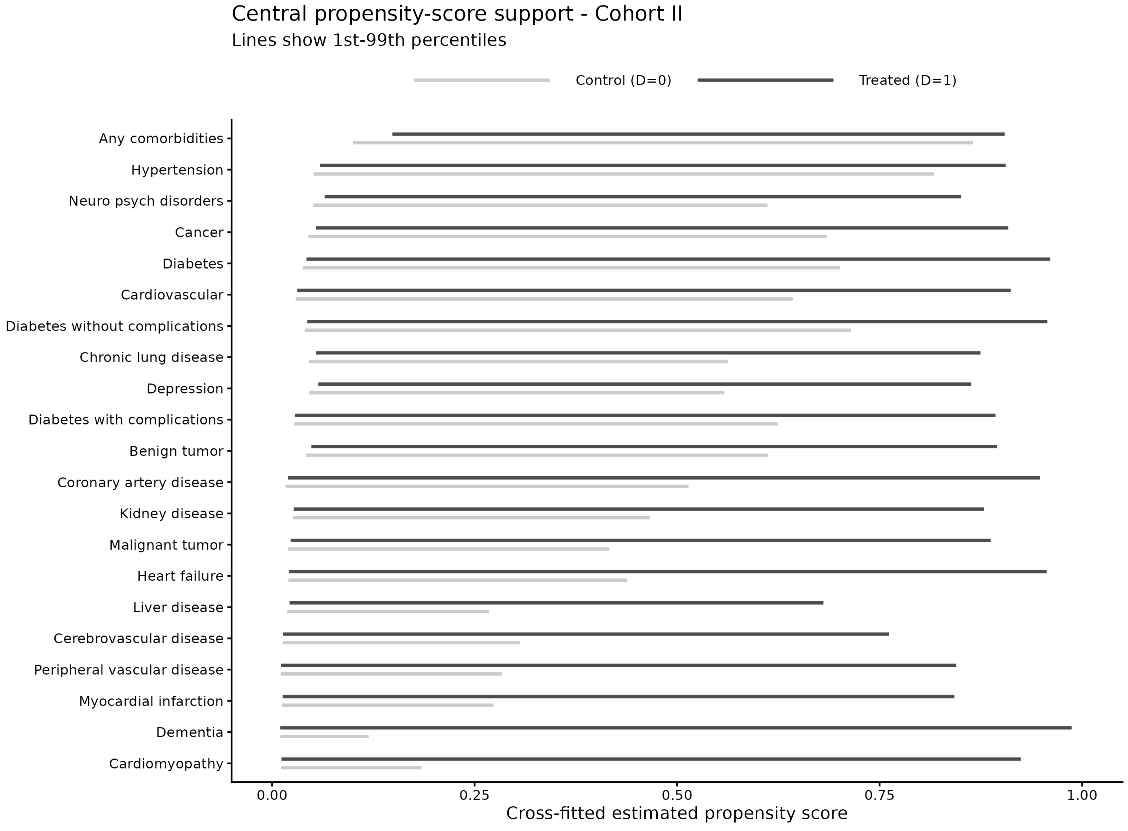


Figure S21: Propensity score overlap across cohorts I-II.

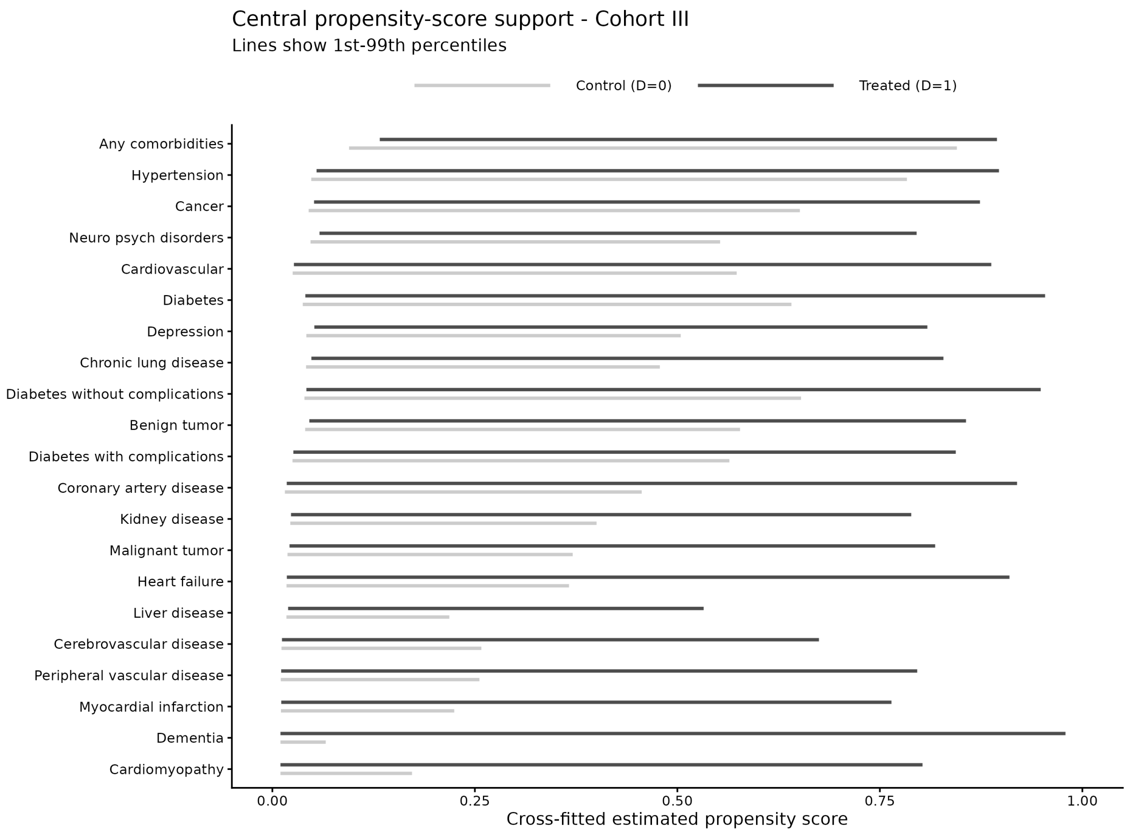


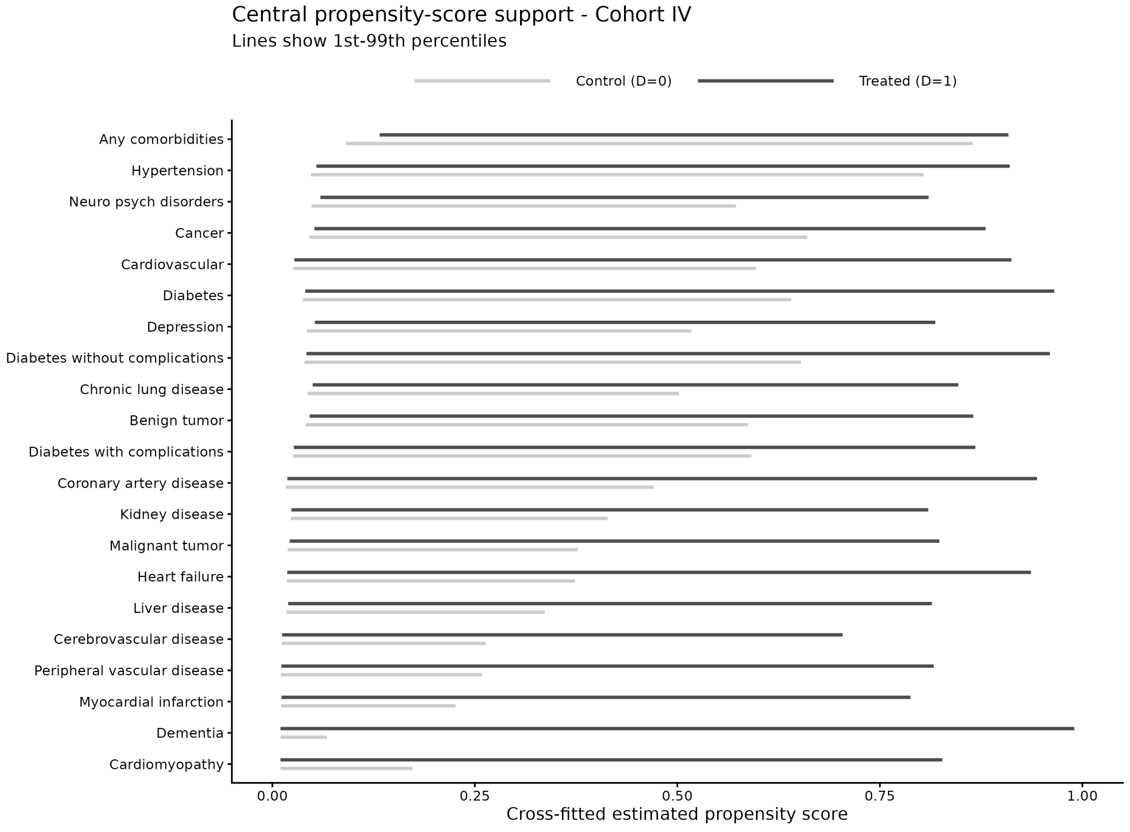


Figure S22: Propensity score overlap across cohorts III-IV.

### References

Berentschot, J.C., Bek, L.M., Drost, M., et al., 2025. Health outcomes up to 3 years and post-exertional malaise in patients after hospitalization for COVID-19: a multicentre prospective cohort study (CO-FLOW). The Lancet Regional Health – Europe 53, 101290. doi:10.1016/j.lanepe.2025.101290.

Chen, Z., Li, B., Chen, Y., Liu, J., Luo, F., Ogunyemi, K.O., Ge, Y., Ke, Y., Yang, Y., Chen, X., et al., 2026. Mapping spatial and social inequities of long covid across the united states: a retrospective cohort study. The Lancet Regional Health–Americas 56.

Hernandez-Romieu, A.C., Leung, S., Mbanya, A., Jackson, B.R., Cope, J.R., Bushman, D., Dixon, M., Brown, J., McLeod, T., Saydah, S., Datta, D., Koplan, K., Lobelo, F., 2021. Health care utilization and clinical characteristics of nonhospitalized adults in an integrated health care system 28–180 days after COVID-19 diagnosis—Georgia, May 2020–March 2021. MMWR Morbidity and Mortality Weekly Report 70, 644–650. doi:10.15585/mmwr.mm7017e3.

Igelström, E., Craig, P., Lewsey, J., Lynch, J., Pearce, A., Katikireddi, S.V., 2022. Causal inference and effect estimation using observational data. Journal of Epidemiology and Community Health 76, 960–966.

Imbens, G.W., 2024. Causal inference in the social sciences. Annual Review of Statistics and Its Application 11.

Institute of Medicine (US) Committee on Health Insurance Status and Its Consequences, 2009. Coverage matters, in: America's Uninsured Crisis: Consequences for Health and Health Care. National Academies Press, Washington, DC. URL: https://www.ncbi.nlm.nih.gov/books/NBK214963/.

Jacobs, E.T., Catalfamo, C.J., Colombo, P.M., et al., 2023. Pre-existing conditions associated with post-acute sequelae of COVID-19. Journal of Autoimmunity 135, 102991. doi:10.1016/j.jaut.2022.102991.

Reese, J.T., Blau, H., Casiraghi, E., Bergquist, T., Loomba, J.J., Callahan, T.J., Laraway, B., Antonescu, C., Coleman, B., Gargano, M., et al., 2023. Generalisable long covid subtypes: findings from the nih n3c and recover programmes. EBioMedicine 87.

Respiratory Therapy, 2025. Repeat covid infections behind most long covid cases—four-year study finds. Published online. URL: https://respiratory-therapy.com/disorders-diseases/infectious-diseases/coronavirus/repeat-covid-infections-most-long-covid-cases-four-year-study-finds/.

Samuels, T.A., Cohen, D., Brancati, F.L., Coresh, J., Kao, W.H.L., 2006. Delayed diagnosis of incident type 2 diabetes mellitus in the ARIC study. American Journal of Managed Care 12, 717–724.

Soriano, J.B., Murthy, S., Marshall, J.C., Relan, P., Diaz, J.V., 2022. A clinical case definition of post-covid-19 condition by a delphi consensus. The Lancet infectious diseases 22, e102–e107.

Subramanian, A., Nirantharakumar, K., Hughes, S., et al., 2022. Symptoms and risk factors for long COVID in non-hospitalized adults. Nature Medicine 28, 1706–1714. doi:10.1038/s41591-022-01909-w.

Tartof, S.Y., Malden, D.E., Liu, I.L.A., Sy, L.S., Lewin, B.J., Williams, J.T.B., Hambidge, S.J., Alpern, J.D., Daley, M.F., Nelson, J.C., McClure, D., Zerbo, O., Henninger, M.L., Fuller, C., Weintraub, E., Saydah, S., Qian, L., 2022. Health care utilization in the 6 months following SARS-CoV-2 infection. JAMA Network Open 5, e2225657. doi:10.1001/jamanetworkopen.2022.25657.

Whitfield, E., White, B., Denaxas, S., Lyratzopoulos, G., 2023. Diagnostic windows in non-neoplastic diseases: a systematic review. British Journal of General Practice .
